# Development and internal validation of a gradient-boosted trees model for prediction of delirium after surgery and anesthesia (the BioCog study)

**DOI:** 10.1101/2024.12.30.24319760

**Authors:** Florian Lammers-Lietz, Levent Akyuez, Diana Boraschi, Friedrich Borchers, Jeroen de Bresser, Sreyoshi Chatterjee, Marta M. Correia, Nikola M. de Lange, Thomas Bernd Dschietzig, Soumyabrata Ghosh, Insa Feinkohl, Izabela Ferreira da Silva, Marinus Fislage, Anna Fournier, Jürgen Gallinat, Daniel Hadzidiakos, Sven Hädel, Fatima Halzl-Yürek, Stefanie Heilmann-Heimbach, Maria Heinrich, Jeroen Hendrikse, Per Hoffmann, Jürgen Janke, Ilse M. J. Kant, Angelie Kraft, Roland Krause, Jochen Kruppa-Scheetz, Simone Kühn, Gunnar Lachmann, Markus Laubach, Christoph Lippert, David K. Menon, Rudolf Mörgeli, Anika Müller, Henk-Jan Mutsaerts, Markus Nöthen, Peter Nürnberg, Kwaku Ofosu, Malte Pietzsch, Sophie K. Piper, Tobias Pischon, Jacobus Preller, Konstanze Scheurer, Reinhard Schneider, Kathrin Scholtz, Peter H. Schreier, Arjen J. C. Slooter, Emmanuel A. Stamatakis, Clarissa von Haefen, Simone J. T. van Montfort, Edwin van Dellen, Hans-Dieter Volk, Simon Weber, Janine Wiebach, Anton Wiehe, Jeanne M. Winterer, Alissa Wolf, Norman Zacharias, Claudia Spies, Georg Winterer, the BioCog consortium

## Abstract

**IMPORTANCE:** Postoperative delirium (POD) is a multietiological condition and affects 20% of older surgical patients. It is associated with poor clinical outcome and increased mortality.

**OBJECTIVE:** We aimed to develop and validate a risk prediction algorithm for POD based on a multimodal biomarker database exploiting preoperative data (predisposing factors) and procedural factors as well as perioperative molecular changes associated with POD (precipitating factors).

**DESIGN:** BioCog is a prospective cohort study conducted from November 2014 to April 2017. Patients were followed up for seven postoperative days after surgery for POD. Gradient-boosted trees (GBT) with nested cross-validation was used for POD prediction.

**SETTING:** Patients aged ≥65 years were enrolled at the anesthesiologic departments of two tertiary care centers.

**EXPOSURE:** All patients underwent surgery with an expected duration of at least 60min. Clinical, neuropsychological, neuroimaging data and blood were collected and clinically well established as well as non-established biomarkers (e.g., gene expression profiling) were measured pre- and postoperatively.

**MAIN OUTCOME:** POD according to DSM 5 until the seventh postoperative day

**RESULTS:** 184 of 929 (20%) patients experienced POD. A GBT algorithm using both preoperative data, characteristics of the intervention and postoperative changes in laboratory parameters achieved the highest area under the curve (0.83, [0.79; 0.86]) with a Brier score of 0.12 (0.12; 0.13).

**CONCLUSIONS AND RELEVANCE:** Models combining predisposing factors with precipitating factors predict POD best. Non-routine laboratory data provide useful information for POD risk prediction, providing relevant results for future studies on the molecular factors of POD. In addition, possibly relevant molecular mechanisms contributing to the development of POD were identified, mostly indicating a dysregulated postoperative immune response. This study constitutes the basis for future hypothesis-driven analyses or implementation of prediction expert system for clinical practice.

## 1 Background

Delirium is an acute disturbance in attention, awareness, cognition, psychomotor behavior and emotional state because of another medical condition. The incidence of postoperative delirium (POD) ranges from 5-50% (1), but is most frequent in older patients (2, 3). POD incidence is assumed to rise in aging populations (4), challenging healthcare systems since it is associated with poor cognitive outcome, hospitalization, treatment costs, re-institutionalization, and mortality (3, 5).

Prehabilitation effectively mitigates postoperative neurocognitive disorders but is time consuming and prediction algorithms are necessary to carefully weigh POD risk against a delay of surgery (6).

Various previous studies have tried to build machine learning-based prediction tools for POD, usually based on retrospective analyses (7–11). The only two prospective studies achieved AUC values of 71% (12) and 74% (13). The prospective Biomarker Development for Postoperative Cognitive Impairment in the Elderly (BioCog) study was conducted with the main goal to improve POD-prediction. We were taking a systems medicine approach with focus on inflammatory alterations and the immune system, the cholinergic system and metabolic changes as well as indicators for early dementia based on an in-depth systematic review (1). Investigations included a wide range of perioperative clinical and neuropsychological parameters, neuroimaging, laboratory investigations and gene expression. Furthermore, the incorporation of precipitating factors may have additional value to predisposing factors.

The primary aim of this study was to develop and internally validate a POD risk index based on multimodal non-routine data intended for use by healthcare professionals to advise patients during medical decision making and allocating healthcare resources.

## 2 Methods

### 2.1 Study design

BioCog (www.biocog.eu, clinicaltrials.gov: NCT02265263, study protocol: (5)) is a prospective observational cohort study with the aim of identifying POD risk factors. The model was developed and internally validated in this cohort. All procedures were approved by the local ethics committees in Berlin, Germany (EA2/092/14) and Utrecht, Netherlands (14–469) and conducted in line with the declaration of Helsinki. All participants gave written informed consent prior to inclusion.

### 2.2 Participants

Male and female patients were enrolled in two tertiary care centers at the Charité– Universitätsmedizin Berlin, Germany, and the University Medical Center Utrecht, Netherlands. Consenting patients aged ≥65 years presenting for elective surgery with an expected duration >60min were included. Patients meeting one of the following criteria were excluded:

- positive screening for pre-existing major neurocognitive disorder defined as a Mini-Mental Status Examination (MMSE) score ≤23 points
- any condition interfering with neurocognitive assessment (severe sensory impairment, neuropsychiatric illness including alcohol and drug dependence, intracranial surgery)
- unavailability for follow-up assessment
- accommodation in an institution due to official or judicial order
- inability to give informed consent

### 2.3 Study procedures

The preoperative data were collected at least one day before surgery including medical history and clinical assessments, neuropsychological testing, blood collection and neuroimaging. Postoperative study visits took place twice daily until the seventh postoperative day.

### 2.4 Outcome

POD during the first seven days after surgery was the primary endpoint. Independently of the routine hospital procedures, POD screening was started in the recovery room and repeated twice per day at 8:00am and 7:00pm (±1h) up to seven days after surgery, by or under supervision of a study physician. POD was defined according to DSM-5 criteria and assessed by prospective screening with three validated tools which were recorded at each visit in accordance with current guidelines (2, 3), to mitigate the known tendency of physicians to underdiagnose POD. Patients were considered delirious if at least one of the following criteria was positive:

- ≥2 points on the Nursing Delirium Screening Scale (Nu-DESC),
- positive Confusion Assessment Method (CAM) score on a general ward,
- positive CAM for the Intensive Care Unit (CAM-ICU) score on an intensive care unit (ICU),
- chart review showing descriptions of delirium.

### 2.5 Clinical assessments

Before surgery, the study team recorded sociodemographic data and information on medication according to Carnahan’s anticholinergic drug scale, health-related quality of life (EQ5D), Mini-Nutritional Assessment (MNA) and Body Mass Index (BMI), tobacco and hazardous alcohol consumption (AUDIT). A functional and physical assessment battery including frailty and walking speed was conducted. Precipitating factors were recorded: duration of surgery and anesthesia, type of anesthetic procedure (regional and/or general anesthesia), type of surgery (intracranial, intrathoracic/-abdominal/-pelvic surgery or peripheral), postoperative pain, prescription of anticholinergic medication daily until the seventh postoperative day, length of hospital and ICU stay as well as complications and postoperative mortality until the 90^th^ postoperative day (eChapters 1.1-1.6).

### 2.6 Neuropsychological data

The preoperative cognitive assessment consisted of a comprehensive screen-based neuropsychological test battery (CANTAB, Cambridge Cognition Ltd., Cambridge, UK) and additional tests (Trail-Making-Test Parts A and B). MMSE score at the screening visit, CANTAB test scores and overall preoperative cognitive impairment (PreCI) were analyzed as risk factors.

PreCI is a dichotomous variable defined through comparison of cognitive test performance with a control group. We used multiple cognitive test parameters with moderate-to-good retest-reliability in the control group (14) and calculated z-scores of the baseline measurement in each test parameter assessed in the control group. The same z-transformation was then applied to the surgical cohort. Z-scores <-1·96 in at least two cognitive test parameters or an averaged z-score <-1·96 was used to define PreCI (eTable 2, eChapter 1.7).

### 2.7 Laboratory parameters

Preoperative serum and plasma samples were collected in supine position immediately before induction of anesthesia after eight hours of fasting and on the morning of the first postoperative day. Blood sampling was performed by trained clinic staff according to a standard operating procedure adapted from the German National Cohort Study (15). Samples were immediately sent to laboratories adjacent to the respective hospital site for analysis, or frozen at -80 °C and shipped to a central biobank at the Molecular Epidemiology Group, Max-Delbrück Center (MDC), Berlin for sample processing and storage. This group distributed samples for additional analyses to Atlas Biolabs GmbH as well as to several partners (Immundiagnostik AG in Bernsheim, Germany, Institute of Protein Biochemistry at Consiglio Nazionale delle Ricerche di Pisa, Immune Study Lab of Institute of Medical Immunology and BIH Center for Regenerative therapies at Charité-Universitätsmedizin Berlin). See eChapter 1.8 for a list of measured molecules. Whenever necessary, values were adjusted for laboratory.

### 2.8 Transcriptomics

Samples for transcriptomic analysis were collected in PAXgene tubes (Qiagen) at the same timepoints as other blood samples. Analyses were performed with Affymetrix Clariom S human microarray for RNA and Affymetrix® Flash Tag™ Biotin HSR (miRNA 4.1 Array Plates) for microRNA analyses (Thermo Fischer, Santa Clara, CA, USA) in a GeneTitan™ Multi-Channel Instrument by Atlas Biolabs GmbH (Berlin).

### 2.9 Neuroimaging

The magnetic resonance imaging (MRI) protocol included whole brain T1-weighted and T2-weighted high-resolution hippocampus imaging and diffusion tensor imaging (DTI). In addition, functional MRI and arterial spin labeling, but have not been considered for prediction due to low between-scanner agreement (inter-class correlation coefficient of 0·36-0·54 for functional connectivity in default mode, salience executive and dorsal attention networks and 0·17-0·39 for quantified cerebral blood flow). We calculated global and regional brain volumes including hippocampal subregions, cortical thickness and curvature from T1-weighted imaging, mean diffusivity, kurtosis and fractional anisotropy from DTI (eChapter 1.10).

### 2.10 Statistics

#### 2.10.1 Estimation of sample size

The rule of thumb of Harrell was used to plan an appropriate number of POD events for a stable prediction model, i.e., ≥10 events per independent variable in logistic regression (16), which was considered adequate for machine learning. Requiring 260 patients with POD for analysis of up to 26 independent predictor variables and expecting a 25% incidence of POD, number of required patients was N=1040. Assuming a drop-out rate of 15%, a total number of N=1200 patients was planned. The initial analysis plan stipulated a training/test split approach for internal validation due to its computational efficiency. Since the study finally achieved a lower cohort size, nested k-fold cross validation was used instead which works more efficiently on small samples.

#### 2.10.2 Analysis of single parameters

For descriptive purposes, associations of pre-/perioperative parameters with POD were analyzed using simple logistic regression. We report odds ratios (OR) with 95% confidence intervals (CI) for the depending variable POD (reference category: no POD). To improve the interpretability of single parameter analyses, standardizing transformations were applied to the raw variables (eChapter 1.11.1) or dichotomized according to clinically relevant cut-off values for presentation of interpretable ORs. Analyses were conducted in R (R Core Team, R Foundation for Statistical Computing, Vienna, Austria) and SPSS (IBM, Armonk, NY). No adjustments for multiple testing were made and therefore, results should be considered exploratory, and we abstain from reporting p-values.

#### 2.10.3 Machine learning

We applied machine learning (gradient boosted trees, GBT) to explore how the interplay of a larger set of predictors would benefit the prediction of POD risk in a bottom-up, data-driven fashion to allow unforeseen predictor-prediction relationships. Data available before surgery as well as data available on the first postoperative day by the latest were eligible for inclusion in machine learning, since these data were deemed useful for preoperative POD risk prediction as well as postoperative re-evaluation of further management.

Variables were assembled into blocks, i.e., preoperative data from the clinical assessment (“Clinical”), characteristics of the surgical intervention (“Precipitants” an “Pain”), preoperative neuroimaging data (“Imaging”), preoperative values) and perioperative difference in laboratory parameters measured in whole blood, plasma or serum (“Blood” and “Blood periop.”), preoperative RNA and µRNA abundance (“RNA” and “µRNA”), as well as perioperative difference in transcript abundance (“RNA periop.”). Different GBT models were built on combinations of various variable blocks. Combinations were selected sequentially, starting with simple models (i.e., using only variables from one block) and then adding further blocks based on the AUC, assumptions on feasibility and relevance for clinical routine. Models using RNA data were evaluated separately since transcript abundance was only available for a subgroup of patients.

The GBT algorithm takes a set of decision trees as weak classifiers and combines them to form a strong classifier. It does so by incrementally adding decision trees during training to steadily improve its previous performance. The sampling of input cases is focused on those cases that were hard to classify before training and individual tree predictions are weighted. During inference, the output is computed through sequential application of each tree. GBT provides a continuous output parameter bounded between 0 and 1, allowing the choice of a clinically relevant cut-off which can be flexibly adapted to address various clinical questions and is inherently able to handle missing data. Area under the receiver operating curve and the Brier score with 95% CI are provided. The Brier score measures the difference between predicted probabilities and actual outcomes, ranging between 0, for perfect prediction, and 1. Models were validated using nested cross-validation. This approach allows model hyperparameter optimization and model selection while avoiding model overfitting. While each of the training datasets is provided to a hyperparameter optimized procedure, the evaluation of hyperparameters is performed using another cross-validation procedure that splits up the each of the provided train dataset into another set of k-folds (see eChapter 1.11.2). Sex-specific analyses have been conducted for the best-performing model. The funding source was not involved in study design, data collection, analysis, interpretation, writing or submitting the manuscript.

## 3 Findings

We recruited 933 patients between November 2014 and April 2017. Table 1 characterizes the sample. The patient flow chart is given in figure 1. Additional details on excluded patients are given in eChapter 2.1. POD assessments were available for 929 patients. 184/929 (20%) patients developed POD.

**Figure 1:**
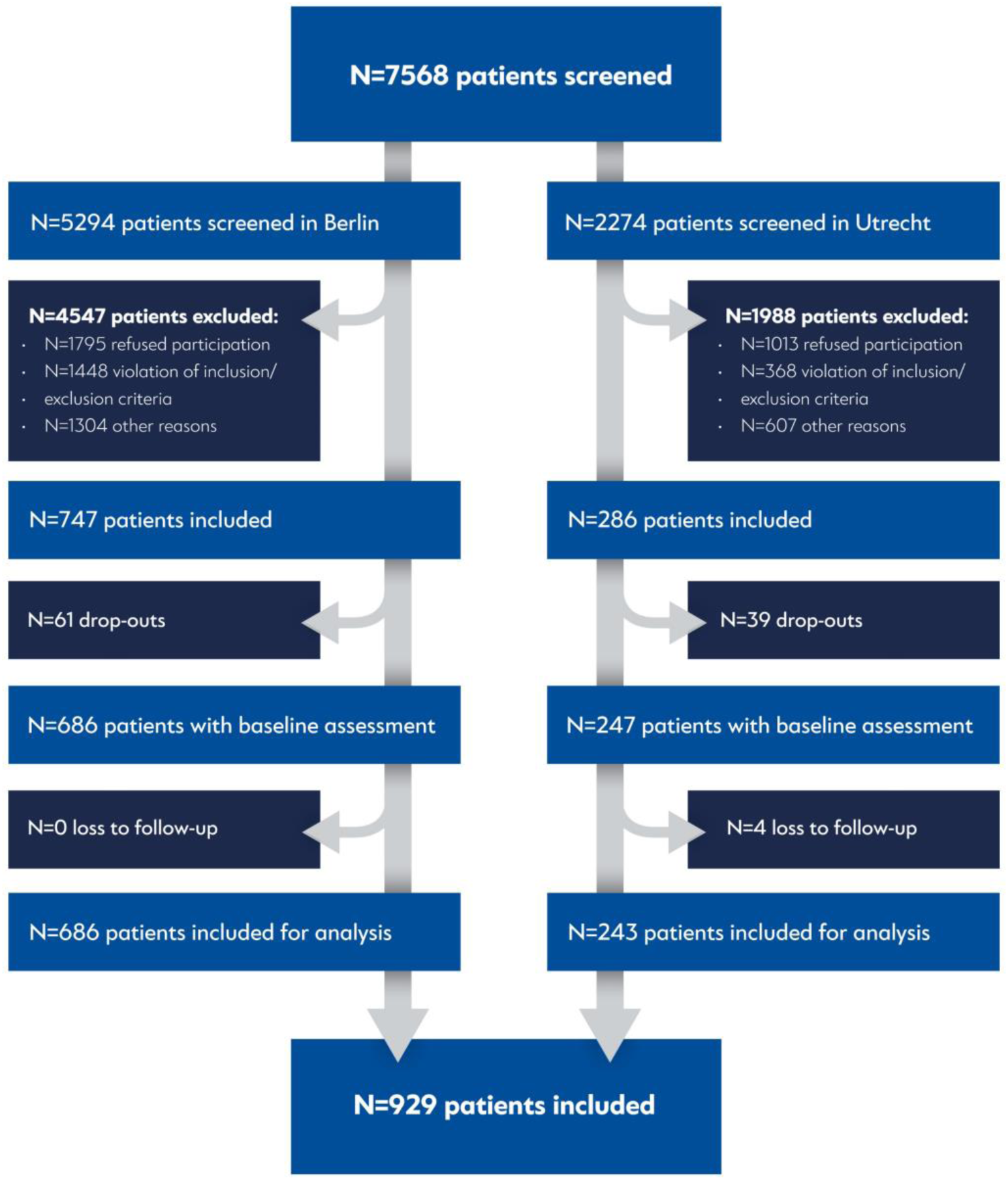
Patient flow chart.

**Table 1:**
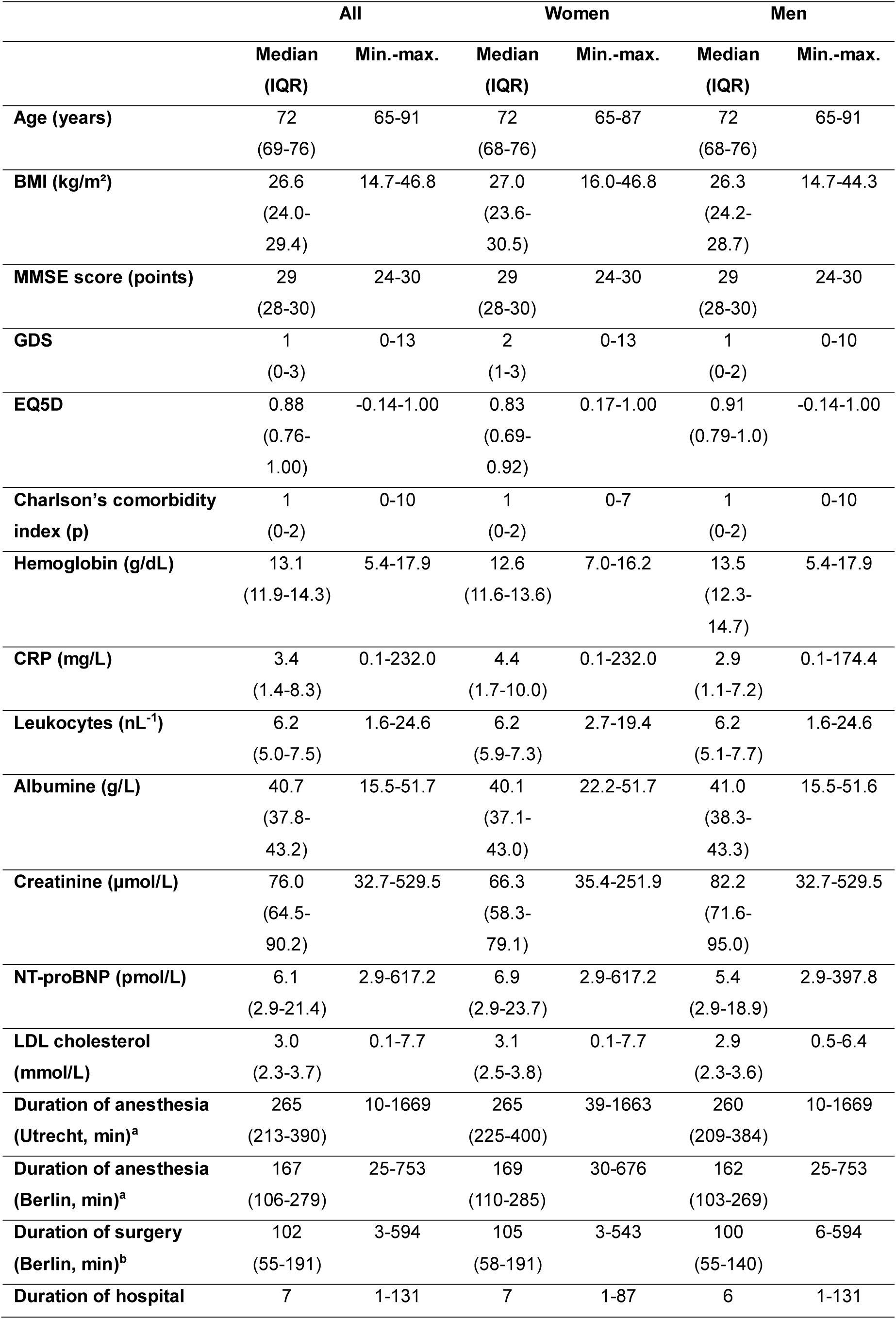
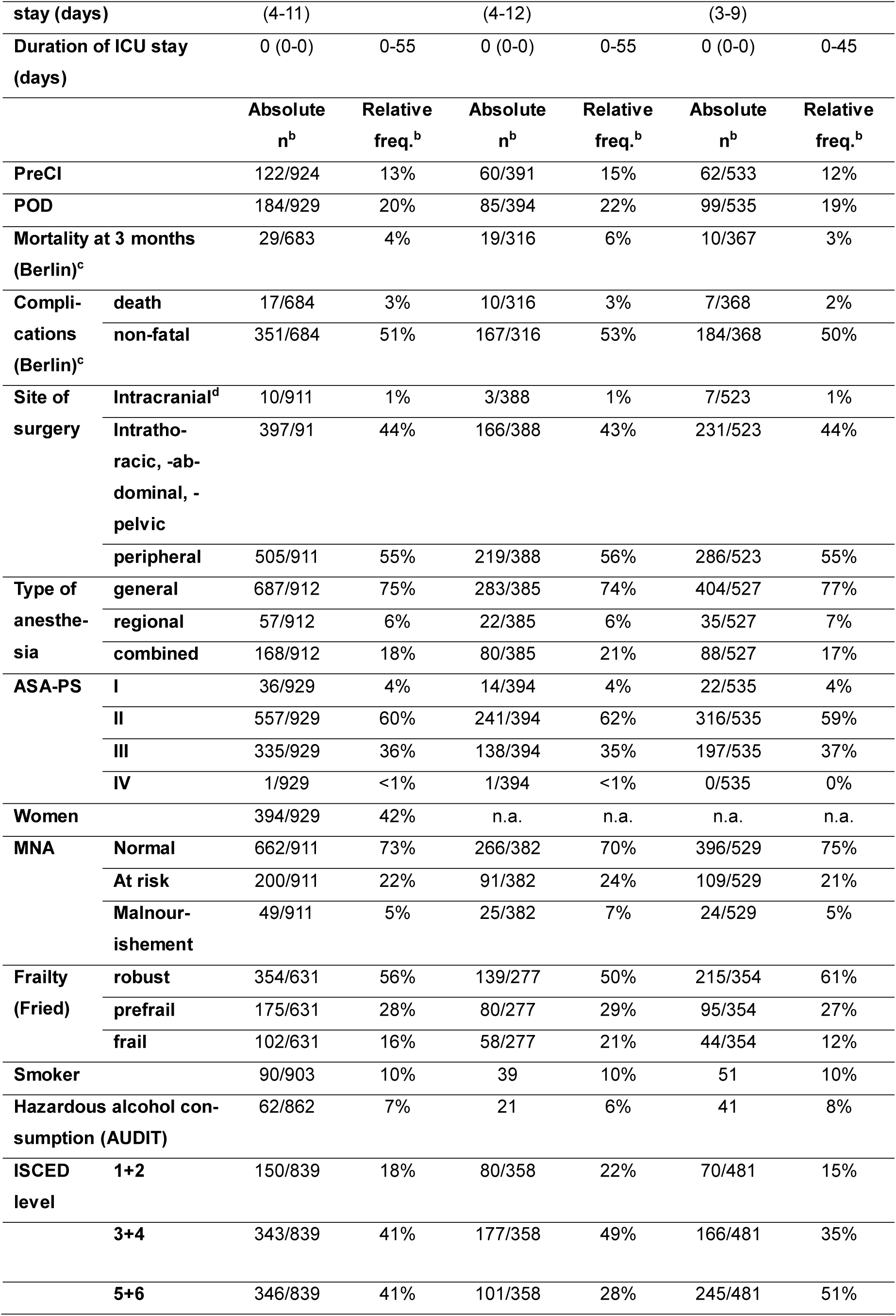
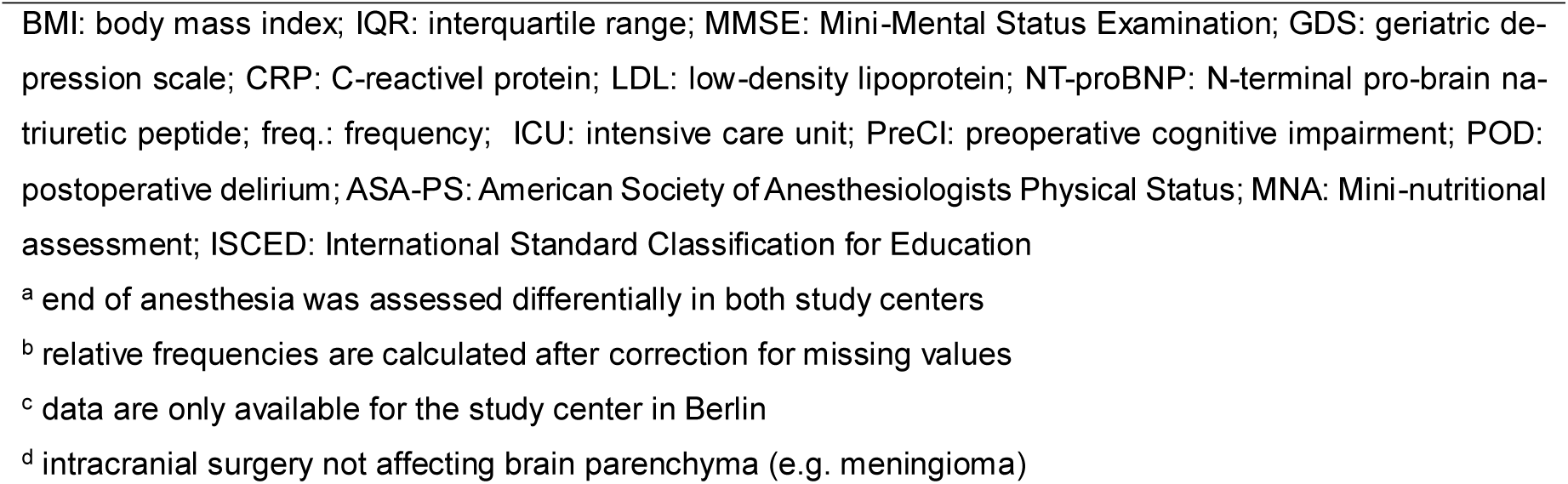
Sample description (N=929)

83/184 (45%) cases of POD were identified in the bedside screening only, 13/184 (7%) cases of POD were diagnosed from chart review only, and 88/184 (48%) cases were proven in both chart review and bedside screening. eFigure 4 and eTable 3 give an overview of daily POD incidence.

### 3.1 POD risk factors

Figure 2 displays unadjusted OR with 95% CIs for preoperative parameters with CIs excluding unity. Sample and effect sizes for all parameters are given in the online-only material.

**Figure 2:**
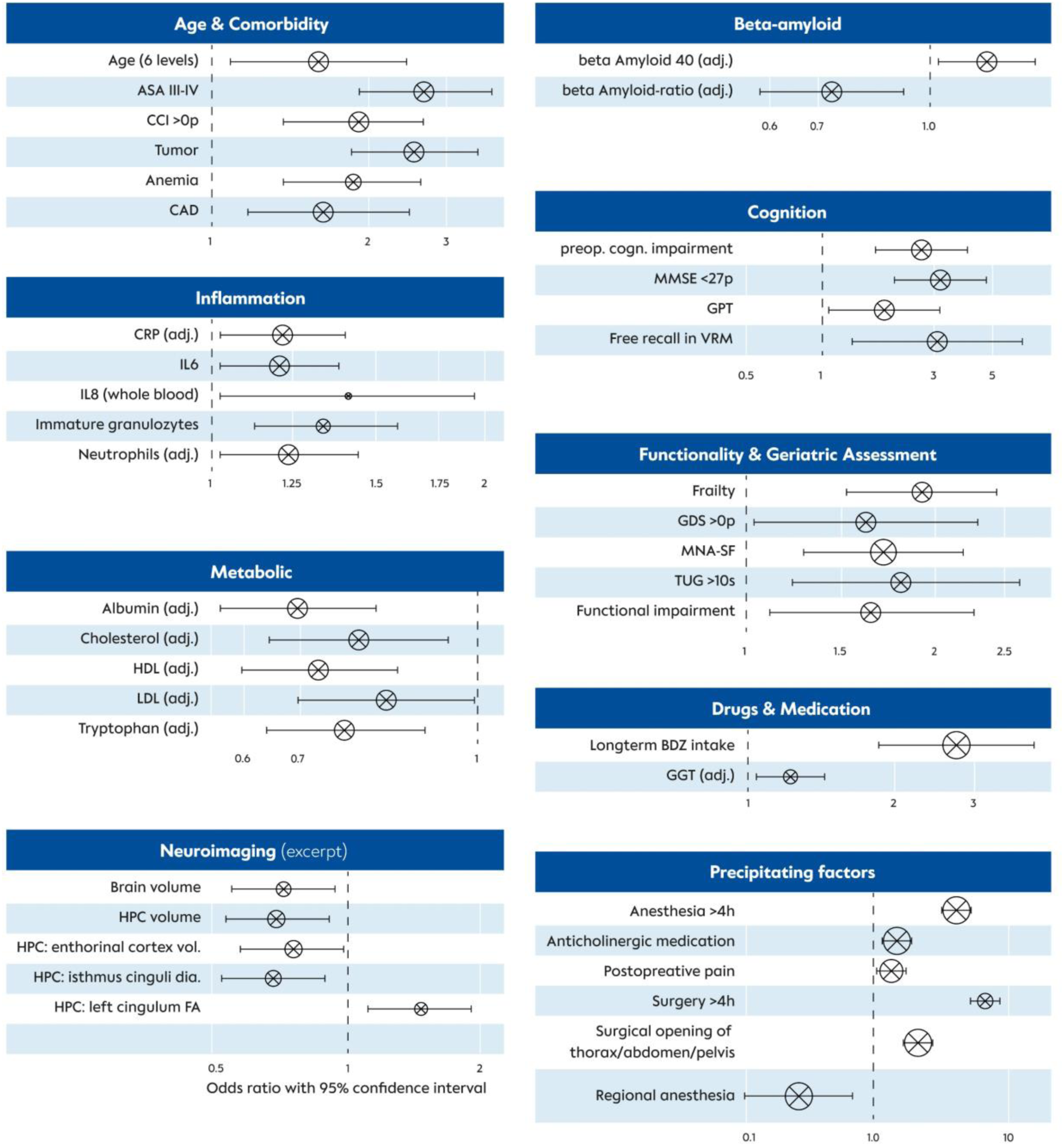
Summary of parameters that were significantly associated with POD. Odds ratios (OR) with 95% confidence interval (95% CI) are shown (only parameters are depicted with CI excluding unity). The diameter of the circle corresponds to the number of available datasets. See also supplementary material 2. The term tumor includes diagnoses of solid malignoma, leukemia and lymphoma. Abbreviations: adj.: adjusted for assessment in different study centers, p: points age & comorbidity: ASA: American Society of Anesthesiologists Physical Status, CAD: coronary artery disease CCI: Charlson comorbidity index inflammation: CRP: C-reactive protein, IL: interleukin cognition: GPT: Grooved Pegboard Test (completion time), MMSE: Mini-mental status examination, preop. cogn. impairment: preoperative cognitive impairment, VRM: Verbal Recognition Memory functionality & geriatric assessment: GDS: Geriatric depression scale, MNA-SF: Mini-nutritional assessment short form, TUG: Timed up-and-go test, frailty refers to Fried’s frailty phenotype metabolic: BMI: body mass index, HDL: high density lipoprotein, LDL: low density lipoprotein drugs & medication: GGT: γ-glutamyltransferase, BDZ: preoperative longterm prescription of benzodiazepines neuroimaging: CA: cornu ammonis, dia.: diameter (cortical thickness), FA: fractional anisotropy, HPC: hippocampus, vol.: volume

Age was directly associated with POD. Among age-related conditions, frailty had the strongest association with POD (OR 1·90 [1·49; 2·44] for category change in Fried’s phenotype), as well as slow walking speed (1·80 [1·22; 2·63] for TUG>10s), malnutrition (OR 1·67 [1·29; 2·16] for category change in MNA-SF), any functional impairment according to Barthel index or IADL assessment (OR 1·59 [1·12; 2·24]) and depressive symptoms (OR 1·57 (1·05; 2·41) for GDS>0).

An MMSE score <27points had a higher OR (3·10 [1·96; 4·85]) for POD than PreCI (OR 2·57 [1·69; 3·88], see also eTable 8 and eFigure 5)

Preoperatively higher levels of cholesterol (standardized, adjusted OR 0·79 [0·65; 0·95]) and associated lipoproteins (HDL and LDL) were protective against POD. A postoperative decrease in triglycerides, cholesterol and LDL were associated with higher POD incidence.

Four inflammatory parameters were positively associated with POD: IL6 (standardized OR 1·19 [1·03; 1·38]), whole blood IL8 (standardized OR 1·42 [1·02; 1·98]) (17), CRP (standardized, adjusted OR 1·20 [1·03; 1·41]), immature granulocyte fraction (standardized OR=1·34 [1·10; 1·63]) and neutrophil count (standardized, adjusted OR 1·22 [1·03; 1·46]). An increase of inflammatory parameters on the first postoperative assessment was associated with higher likelihood of POD (CRP: standardized, adjusted OR 1·59 [1·14; 2·21], IL6: standardized OR 1·76 [1·48; 2·09], and IL8: standardized OR 1·96 [1·18; 3·24]). Cellular immune response showed a more complex association with POD: Whereas a postoperative increase in leukocytes (standardized, adjusted OR 1·36 [1·12; 1·64]) and neutrophiles (standardized, adjusted OR 1·47 [1·2; 1·81]) was associated with POD, an increase in lymphocytes lowered the odds for POD (standardized, adjusted OR 0·66 [0·54; 0·81]). Higher levels tryptophan (standardized, laboratory-adjusted OR 0·74 [0·62; 0·89]) and albumin (standardized, adjusted OR 0·68 [0·56; 0·81]) also lowered the odds for POD. A higher plasma β-amyloid 42/40-ratio was found to be related to a lower POD likelihood (standardized, adjusted OR 0·74 [0·56; 0·93]), but this association seemed to be driven by increased POD risk in patients with higher levels of β-amyloid 40 (standardized, adjusted OR 1·20 [1·02; 1·41]).

Both higher preoperative γ-glutamyltransferase levels (standardized, adjusted OR 1·24 [1·06; 1·47]) and a postoperative decrease (standardized, adjusted OR 0·81 [0·68; 0·98]) were associated with POD. A postoperative increase in transaminases was associated with POD. Postoperatively decreasing levels of oxidative stress indicated by nitrotyrosine levels (standardized OR 0·72 [0·53; 0·98]) and nitrous oxide production indicated by homoarginine levels (standardized OR 0·48 [0·31; 0·73]) were associated with increased POD risk.

Longer duration of anesthesia (OR 4·42 [3·15; 6·27] for >4h) and surgery (OR 7·44 [4·84; 11·50] for >4h) as well as blood loss (standardized, adjusted OR for perioperative changes in Hb: 0·76 [0·63; 0·91], thrombocytes: 0·57 [0·46; 0·69], and albumin: 0·66 [0·54; 0·81]) were associated with POD. Compared to general anesthesia, surgery performed in regional anesthesia was associated with lower rates of POD (0·29 [0·09; 0·72]). Surgery with opening of thorax, abdomen or pelvis was associated with increased rates of POD compared to peripheral surgery (OR 3·00 [2·13; 4·25]). Pain (OR 2·16 [1·55; 3·01]) or intake of any anticholinergic medication (OR 2·35 [1·50; 3·84]) at least once during follow-up until the seventh postoperative day were both associated with POD (see also eTables 11-12, eFigures 6 and 7).

Various associations of structural MRI-derived parameters were observed (complete results: eFigure 8, eTable 15), we would like to emphasize a protective association of POD with global brain volume (standardized OR 0·71 [0·55; 0·92]) as well as hippocampus volume (standardized OR [0·67 (0·53; 0·86]).

### 3.2 Machine learning-prediction of POD

Figure 3 displays AUC for the GBT models built from the most relevant combinations of variable blocks and eTable 16 provides details on the model performance. Among the models using only preoperative data, the model using only clinical data performed best (AUC 0·76 [0·69; 0·81], Brier score: 0·14 [0·13; 0·16]). Adding preoperative blood or RNA data to the model did not improve the AUC.

**Figure 3:**
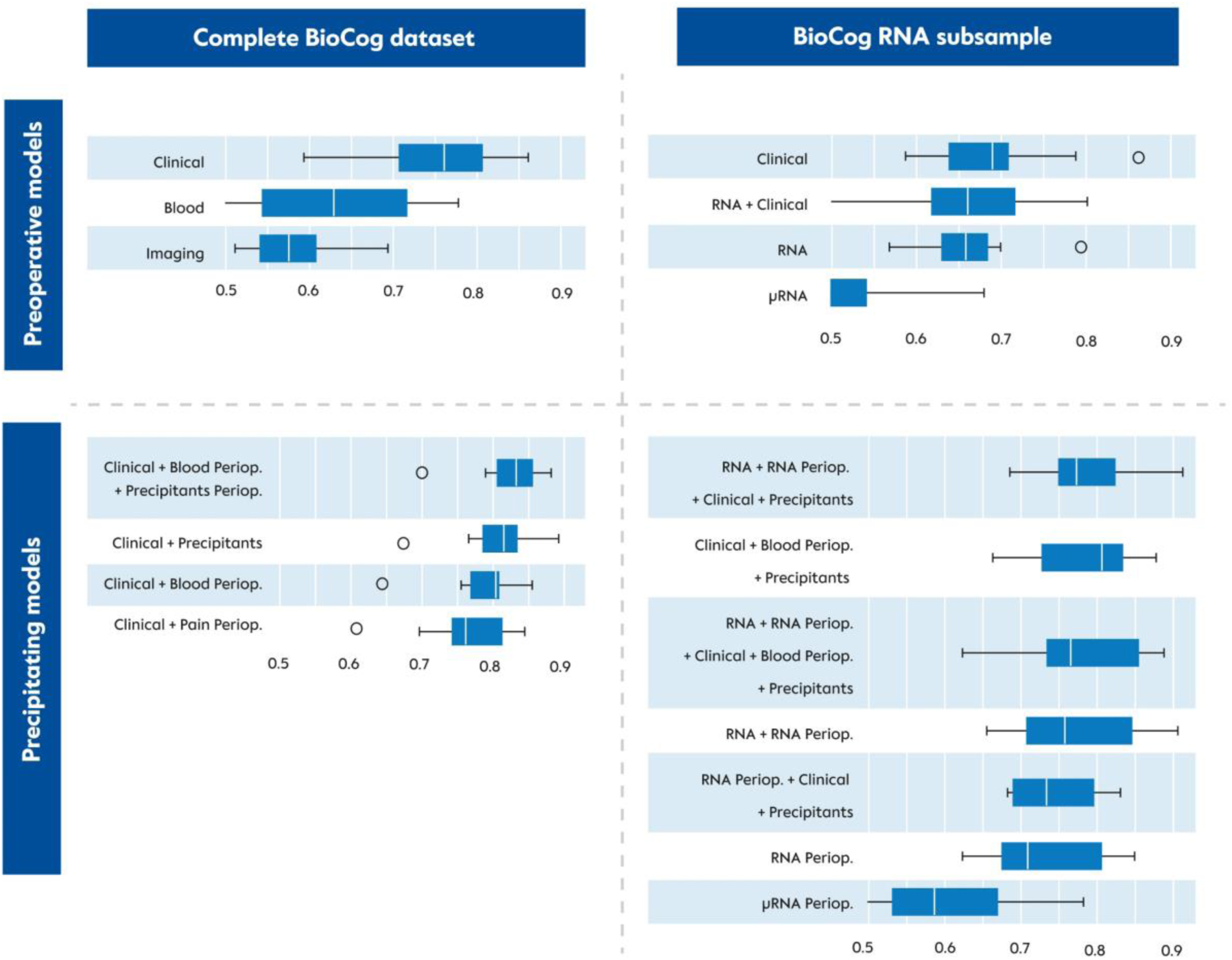
Boxplot displaying area under the curve (AUC) of the receiver-operating characteristic (ROC). A value of 1 indicates 100% sensitivity at 100% specificity, whereas a value of 0.5 indicates indiscriminability of the model for POD. Each model evaluates a different combination of available datasets, as indicated on the y-axis. Abbreviations: periop.: perioperative (referring to precipitating factors, e.g., pain or medication, and perioperative changes in molecule abundance), RNA: transcriptomic data features.

The model AUC was increased considerably by adding characteristics of the intervention (“Precipitants”) and perioperative changes in laboratory parameters (“Blood periop.”) to the clinical data, and the highest overall AUC was achieved by a model using these three blocks of data (AUC 0·83 [0·79; 0·86], Brier score 0·12 [0·12; 0·13]). The most important variables in this model are displayed in figure 4, and eFigure 10 displays sex-stratified Receiver-Operating Curves. This model also provided the highest performance in the subgroup of patients with RNA data (AUC 0·78 [0·73; 0·83], Brier score 0·15 [0·14; 0·16]), and adding transcript data to the model did not improve AUC. However, a model exploiting only pre- and and postoperative RNA data (“RNA+RNA periop.”) showed almost identical performance (AUC 0·77 [0·71; 0·78], Brier Score 0·15 [0·14; 0·16]).

**Figure 4:**
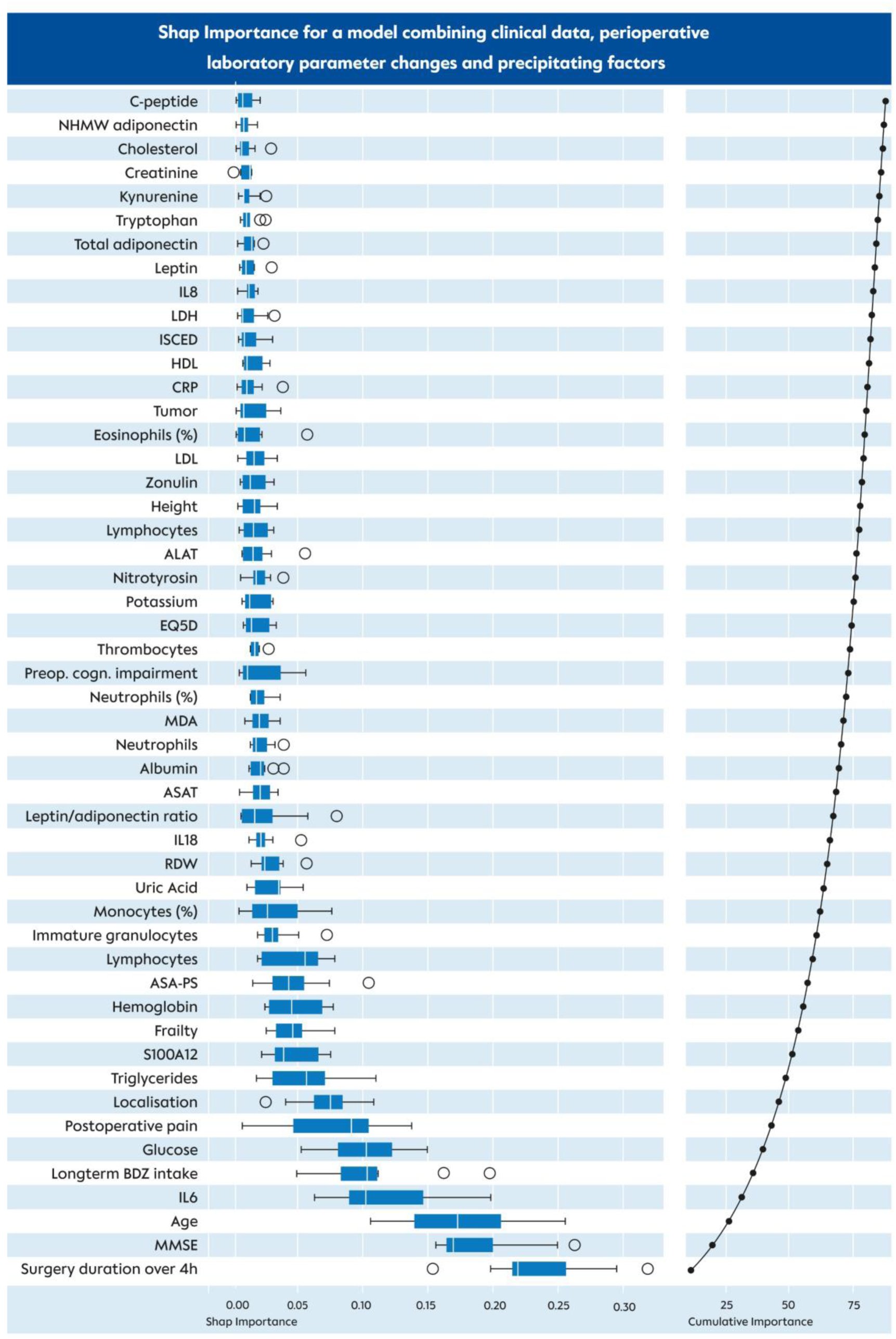
Feature importance of the model with the highest predictive performance. Tumor diagnosis includes solid malignancies, lymphoma and leukemia. Abbreviations: ALAT: alanin-aminotransferase, ASAT: aspartat-aminotransferase, BDZ: benzodiazepine, CRP: C-reactive protein, HDL: high-density lipoprotein, IL: interleukin, ISCED: International Standard Classification of Education, LDH: lactate dehydrogenase, LDL: low-density lipoprotein, MDA: malondialdehyde MMSE: Mini-mental status examination, NHWM: non-high molecular weight, postop.: postoperative, preop. cogn. impairment: preoperativ e cognitive impairment, RDW: red cell distribution width

eFigure 10 displays the relevant transcripts from the “RNA+RNA periop.” model: The perioperative changes in mRNA abundance were more often predictive of POD than preoperative abundance. Most important transcripts were *BTN3A1, LAP3, DSN1, HPGD* and *KIF4B*. Notably, both preoperative *JAK2* and circular *JAK2* mRNA were predictive of POD. In an exploratory Cox regression analysis, we found that a considerable number of transcripts (*HPGD, BTN3A1, LAP3, JAK2* and circular *JAK2*) were also associated with postoperative mortality (eTable 17, eFigure 11).

## 4 Discussion

We estimated POD prediction algorithms based on clinical, neuropsychological, blood-based and neuroimaging data. This is the first approach to POD prediction using data-driven analysis of a prospectively collected multimodal dataset. By aggregating clinical preoperative data, precipitating factors with preoperative laboratory values and postoperative changes, the model achieved good discriminability (AUC 83%) with good model fit.

Previous approaches used retrospectively collected data or merged heterogenous data from multiple studies (9, 11, 13). The only prospective study (SAGES) achieved an AUC of 71% using machine learning in preoperatively available clinical data (12). Our model solely relying on preoperative clinical data achieved similar performance (AUC 76%), and no improvement by adding preoperative non-routine data was achieved. Hence, thorough preoperative clinical evaluation to identify patients at risk can be considered a suitable approach in clinical routine. However, using algorithms as a diagnostic expert device can support quantifying POD risk and drive the establishment of POD risk assessment in routine clinical practice. Results suggest that information about intervention and postoperative course can improve the model to an AUC of 80%. Although models using precipitating data are intended for risk monitoring rather than prediction, relevant information is usually available before surgery, i.e., estimated duration of intervention and expected postoperative pain, and may be used for prediction as well.

Our analyses suggest that models exploiting precipitating factors and perioperative laboratory assessments can considerably improve POD risk monitoring, but neuroimaging and transcriptomic data do not. However, gene expression data may be of particular interest for further studies, since in the subgroup of patients with RNA data, a model exploiting only mRNA achieved a similar AUC (77%) compared to the best performing model (78%). A perioperative risk monitoring algorithm based on two gene expression analyses could relieve medical staff from extensive clinical assessments, be more cost-effective than using multiple independent laboratory assays and avoids data aggregation from different sources.

Many of the most predictive transcripts were mRNA of ubiquitously expressed genes involved in major molecular mechanisms such as cell proliferation (e.g., *DSN1*, *LAP3*, *KIF4B* and *JAK*). This may suggest that POD is a heterogeneous phenomenon originating via distinct molecular pathways. These central molecular nodes may be the common denominator among different POD subgroups, but nevertheless suitable for prediction. Certain transcripts suggest involvement of γδT-cells in neuroinflammation and neuroplasticity (*BTN3A1*)(18), metabolic dysregulation and autophagy (*LAP3*)(19), proliferation (*DSN1*, *KIF4B*)(20), interaction with the immune system (*JAK*)(21, 22), and senescence (*HPGD*)(23). These transcripts are also associated with 3-month mortality (eChapter 2.3). Above-mentioned molecules S100A12 (24), interleukins and zonulin (25) point to certain immune response pathways, which may be related to neurotransmitter dysbalance by tryptophan and kynurenine metabolism (26). Some of the identified molecular targets have already been discussed with respect to neurodegeneration, i.e., malondialdehyde, nitrotyrosine (27), metabolites of the kynurenine pathway (28), S100A12 (24) and zonulin (25).

The BioCog study is small in relation to the wide spectrum of parameters included in our database. To fully exploit the potential of machine learning, larger samples are necessary. The current sample excluded patients with MMSE score ≤23, but brain atrophy may be relevant biomarker in patients with preoperative cognitive impairment. Since external validation in an independent dataset is pending, we have used nested cross validation as an internal validation procedure, which is robust against overfitting. The focus of this manuscript is prediction, whereas a molecular causal model cannot be addressed here. E.g., single variable analyses have not been adjusted for confounders, warranting further analysis. The best model was chosen by ROC-AUC, which is a measure of discrimination in diagnostic testing. For prognostic questions, outcome probability estimation is preferable and more evaluations are needed (29).

POD screening was performed according to the evidence-based standard that measure POD at least twice a day and has a comprehensive geriatric assessment included to describe the clinical entity of this population. The clinical phenomenology was structured and annotated according to this standard (2).

BioCog has made advancements towards POD prediction and will facilitate comprehensive hypothesis-driven analyses including subgrouping of patients for better understanding of pathophysiological processes and conception of interventional studies. Our dataset can guide prevention strategies to reduce POD, e.g., via the JAK-pathway (22).

## Supporting information

eChapter

## 5 Acknowledgements

We thank the Koordinierungszentrum für Klinische Studien (KKS Berlin), and especially Alexander Krannich, and ALTA (Siena, Italy) who provided additional administrative and coordinating services throughout the study. Multimodal data management was conducted by Pharmaimage Biomarker Solutions GmbH. We thank our colleagues who participated as MD students and team of study nurses, especially Tuba Gülmez, Emmanuel Keller, Mario Lamping, Helene Michler, Juliane Dörfler, Zdravka Bosancic, Fieras Nosierat, Irene Mergele, Victoria Windmann and Anna Nottbrock. The authors further wish to thank the team of the student apprentices/interns of the Department of Anesthesiology at the Charité-Universitätsmedizin Berlin. Magnet resonance imaging has been supported by the Berlin Center for Advanced Neuroimaging core staff, especially Stefan Hetzer. Henning Krampe supported the study by recruiting and supervising students for neuropsychological testing.

## 6 Funding

The research leading to these results has received funding from the European Union Seventh Framework Program [FP7/2007-2013] under grant agreement n° 602461.

## 7 Data availability

Due to the protection of intellectual property, machine learning algorithms will not be made publicly available, but can be obtained from Dr. Winterer after signing a confidentiality agreement. Participant data may be made available upon request following publication to researchers who provide a methodologically sound proposal in accordance with applicable legal and regulatory restrictions. Proposals for data analysis must be directed to both. To gain access, requesting researchers will need to sign a data access agreement. Analyses will be limited to those approved in appropriate ethics and governance arrangements. All study documents which do not identify individuals (e.g. study protocol, informed consent form) will be freely available on request.

## 8 Statement on sex and gender equity in research

Data on the patients’ gender have not been collected, and we refer to “sex” throughout the manuscript. Although the best-performing model was found to have similar discriminability in women and men, analyses in the online-only material have neither been adjusted nor disaggregated for sex, although further in-depth analyses would require this step to yield valid results.

## 9 Protein products of mentioned genes

*BTN3A1* - butyrophilin

*LAP3* - leucine aminopeptidase 3

*DSN1* - DSN1 component of MIS12 kinetochore complex

*HPGD* - 15-hydroxyprostaglandin dehydrogenase

*KIF4B* - kinesin family member 4B

## 10 Author contributions

Florian Lammers-Lietz: Writing – Original Draft, Investigation, Validation, Visualization; Levent Akyuez; Methodology, Investigation, Resources; Diana Boraschi: Methodology, Formal Analysis, Writing – Review & Editing; Friedrich Borchers: Conceptualization, Investigation, Formal Analysis, Validation, Data Curation, Writing – Review and Editing; Jeroen de Bresser: Methodology, Investigation, Formal Analysis; Sreyoshi Chatterjee: Investigation, Formal Analysis; Marta M. Correia: Formal Analysis; Nikola M. de Lange: Methodology, Investigation, Formal Analysis; Thomas Bernd Dschietzig: Conceptualization, Methodology, Investigation, Formal Analysis, Supervision, Funding Acquisition; Soumyabrata Ghosh: Methodology, Investigation, Formal Analysis; Insa Feinkohl: Conceptualization, Methodology, Formal Analysis, Data Curation; Izabela Ferreira da Silva: Investigation, Formal Analysis; Marinus Fislage: Investigation, Writing – Original Draft, Formal Analysis; Anna Fournier: Methodology, Investigation, Formal Analysis; Jürgen Gallinat: Methodology, Investigation, Formal Analysis, Supervision; Daniel Hadzidiakos: Conceptualization, Methodology, Investigation, Data Curation, Validation; Sven Hädel: Software, Data Curation; Fatima Halzl-Yürek: Investigation; Stefanie Heilmann-Heimbach: Methodology, Investigation, Formal Analysis; Maria Heinrich: Investigation, Data Curation, Validation; Jeroen Hendrikse: Methodology, Investigation, Formal Analysis; Per Hoffmann: Methodology, Investigation, Formal Analysis; Jürgen Janke: Data Curation; Ilse M. J. Kant: Investigation, Methodology, Validation; Angelie Kraft: Formal Analysis; Roland Krause: Methodology, Investigation, Formal Analysis, Supervision; Jochen Kruppa-Scheetz: Methodology, Formal Analysis, Data Curation; Simone Kühn: Conceptualization, Methodology, Investigation, Formal Analysis, Supervision, Funding Acquisition; Gunnar Lachmann: Investigation, Methodology; Markus Laubach: Investigation; Christoph Lippert: Methodology, Investigation, Formal Analysis; David K. Menon: Conceptualization, Resources, Supervision, Writing – Review and Editing; Rudolf Mörgeli: Methodology, Investigation; Anika Müller: Methodology, Investigation; Henk-Jan Mutsaerts: Methodology, Investigation, Formal Analysis; Markus Nöthen: Methodology, Investigation, Formal Analysis; Peter Nürnberg: Conceptualization, Methodology, Supervision, Project Administration, Funding Acquisition; Kwaku Ofosu: Investigation; Malte Pietzsch: Conceptualization, Funding Acquisition, Methodology; Sophie K. Piper: Methodology, Formal Analysis; Tobias Pischon: Conceptualization, Methodology, Resources, Data Curation, Writing – Original Draft, Supervision, Project Administration, Funding Acquisition; Jacobus Preller: Supervision, Conceptualization, Methodology, Investigation, Formal Analysis, Writing – Review and Editing, Project Administration; Konstanze Scheurer: Project Administration; Reinhard Schneider: Methodology, Investigation, Formal Analysis; Kathrin Scholtz: Project Administration; Peter H. Schreier: Writing – Review and Editing; Arjen J. C. Slooter: Conceptualization, Funding Acquisition, Methodology, Writing – Review and Editing, Administration; Emmanuel A. Stamatakis: Supervision, Conceptualization, Funding Acquisition, Investigation, Methodology, Writing – Review and Editing, Project Administration; Clarissa von Haefen: Data Curation, Project Administration; Simone J. T. van Montfort: Investigation, Validation; Edwin van Dellen: Methodology, Investigation; Hans-Dieter Volk: Resources, Writing – Review and Editing; Simon Weber: Conceptualization, Funding Acquisition, Methodology; Janine Wiebach: Methodology, Formal Analysis, Software, Data Curation; Anton Wiehe: Methodology, Formal Analysis, Visualization, Software; Jeanne M. Winterer: Investigation; Alissa Wolf: Methodology, Formal Analysis, Project Administration; Norman Zacharias: Methodology, Formal Analysis, Investigation; Claudia Spies: Conceptualization, Methodology, Validation, Software, Resources, Data Curation, Writing – Original Draft, Supervision, Project Administration, Funding Acquisition; Georg Winterer: Conceptualization, Methodology, Software, Resources, Data Curation, Writing – Original Draft, Supervision, Project Administration, Funding Acquisition; Florian Lammers-Lietz and Georg Winterer have verified the underlying data.

## 11 Assistance in scientific writing

Neither artificial intelligence tools nor a professional medical writer has assisted in manuscript preparation.

## 12 Conflict of interest

Georg Winterer is currently licensing a Class IIa medical device (web-based software tool for risk prediction of POD and POCD in clinical practice). Dr. Winterer is CEO of PharmaImage Biomarker Solutions GmbH Berlin (Germany) and President of its subsidiary Pharmaimage Biomarkers Incl. (Cambridge, MA, USA).

Dr. Spies, Dr. Winterer, Dr. Boraschi, Dr. Dschietzig, Dr. Kühn, Dr. Nürnberg, Dr. Pischon, Dr. Pietzsch, Dr. Slooter, Dr. Stamatakis, Dr. Weber, report grants from the European Commission during the conduct of the study. Dr. Winterer reports grants from the Deutsche Forschungsgemeinschaft (DFG)/German Research Society and from the German Ministry of Health. Dr. Spies reports grants from DFG/German Research Society, Einstein Foundation Berlin, Deutsches Zentrum für Luft-und Raumfahrt e.V. (DLR)/German Aerospace Center, Projektträger im DLR/Projec Management Agency, Gemeinsamer Bundesausschuss (GBA)/Federal Joint Committee, inneruniversity grants, Stifterverband/Non-Profit Society Promoting Science and Education, European Society of Anesthesiology and Intensive Care, BMWI – Federal Ministry of Economic Affairs and Climate Action, Dr. F. Köhler Chemie GmbH, Sintetica GmbH, Max-Planck-Gesellschaft zur Förderung der Wissenschaft e.V., Metronic, BMBF – Federal Ministry of Education and Research, Robert Koch Institute and payments by Georg Thieme Verlag, board activity for Prothor, Takeda Pharmaceutical Company Ltd., Lynx Health Science GmbH, AWMF (Association of the Scientific Medical Societies in Germany), DFG, Deutsche Akademie der Naturforscher Leopoldina e.V. (German National Academy of Sciences Leopoldina), Berliner Medizinische Gesellschaft, European Society of Intensive Care Medicine (ESICM), European Society of Anaesthesiology and Intensive Care (ESAIC), Deutsche Gesellschaft für Anästhesiologie und Intensivmedizin (DGAI)/German Society of Anaesthesiology and Intensive Care Medicine, German Interdisciplinary Association for Intensive Care and Emergency Medicine (DIVI) as well as patents 15753 627.7, PCT/EP 2015/067731, 3 174 588, 10 2014 215 211.9, 10 2018 114 364.8, 10 2018 110 275.5, 50 2015 010 534.8, 50 2015 010 347.7, 10 2014 215 212.7.

Gunnar Lachmann and Maria Heinrich report grants from the BIH Charité Clinician Scientist Program during conduct of the study. Dr. Dschietzig reports personal fees from Immundiagnostik AG during the conduct of the study. Dr. Lammers-Lietz and Anton Wiehe report personal fees from Pharmaimage Biomarker Solutions GmbH during the conduct of the study. Dr. Lachmann reports personal fees from Sobi, the University of Zurich and Thieme outside the submitted work. Dr. Wolf receives fees from the Kompetenz-Centrum Qualitätssicherung. Dr. Stamatakis reports funding from Stephen Erskine Fellowship from Queens’ College of the University of Cambridge, UK outside the BioCog study. Dr. Bresser reports funding from Alzheimer Nederland outside of the study. Dr. Gallinat received funding from the German Research Foundation (DFG), Federal Ministry of Education and Research (BMBF) and received payment for five lectures and presentations with about 1.500 € per presentation sponsored by Lundbeck, Janssen-Cilag and Boehringer. Dr. Heilmann-Heimbach receives personal fees from Life&Brain GmbH.

None of the other authors have a conflict of interest to declare.

## Notes

### Author Declarations

Ethics committees of the Charite-Universitaetsmedizin Berlin, Germany, (EA2/092/14) and University Medical Center Utrecht (UMC), Utrecht University, Netherlands, (14-469) gave ethical approval of this work.

