## Supplementary material for "Development and internal validation of a gradient-boosted trees model for prediction of delirium after surgery and anesthesia (the BioCog study)": eChapter

### Table of supplementary content

eChapter 1: Methods ... 4
eChapter 1.1: Data collection and clinical assessments ... 4
eChapter 1.2: Sociodemographic data ... 4
eChapter 1.3: Comorbidities ... 4
eChapter 1.4: Functionality and geriatric examination ... 4
eChapter 1.5: Nutrition and lifestyle factors ... 5
eChapter 1.6: Perioperative parameters ... 5
eChapter 1.7: Neuropsychological testing ... 6
eChapter 1.8: Laboratory values ... 7
eChapter 1.9: Transcriptomics ... 8
eChapter 1.10: Neuroimaging ... 8
eChapter 1.10.1: Image acquisition ... 8
eChapter 1.10.2: Neuroimaging data processing ... 9
eChapter 1.10.3: Structured database ... 10
eChapter 1.11: Statistical analyses ... 10
eChapter 1.11.1: Single variable analyses ... 10
eChapter 1.11.2: Machine learning approach ... 11
eChapter 2: Supplementary Results ... 16
eChapter 2.1: Description of excluded patients ... 16
eChapter 2.1.1: Utrecht ... 16
eChapter 2.1.2: Berlin ... 16
eChapter 2.2: Overview on missing data ... 17
eChapter 2.3: Associations between transcript abundance and mortality ... 52
eChapter 3: References ... 54

### eChapter 1: Methods

#### eChapter 1.1: Data collection and clinical assessments

Demographic and clinical data were collected by a structured patient interview and clinical examination, questionnaires, and consultation of the patient's case file. Whenever possible, additional medical reports have been obtained from the patients. All data have been collected by qualified study physicians or trained research nurses and study assistants under supervision of a study physician.

#### eChapter 1.2: Sociodemographic data

Sociodemographic data on age, sex and education were collected. Education was defined by the WHO 1997 International Standard Classification of Education (ISCED 1997). ISCED levels were aggregated in three categories referring to level 1-2, level 3-4 (primary and secondary education) and levels ≥5 (tertiary education). Sex could only be reported as "male" or "female", neglecting other entities. The study protocol did not consider an explicit differentiation between sex and gender, and the data presented here are likely to correspond to biological sex. There was no particular advice on how data on the patient's sex should be recorded – i.e. self-report as well as extraction from the patient file were equally legitimate sources of information according to the study protocol.

#### eChapter 1.3: Comorbidities

Clinical data comprise the Charlson Comorbidity Index, American Society of Anesthesiologists Physical Status (ASA PS) and distinct preoperative medical conditions. The ASA PS scale was dichotomized into ASA PS I-II and III-IV for analyses. Preoperative diagnoses of arterial hypertension, coronary artery disease, diabetes mellitus (with either dietetic/oral treatment or receiving insulin therapy), history of stroke or transient ischemic attacks and preoperative tumor, leukemia or lymphoma were recorded and analyzed separately.

Depressive symptoms were assessed separately using the Geriatric Depression Scale (GDS) score. Ipsative imputation was used to adjust for missing answers to single items from the GDS. For the Charlson Comorbidity Index, we merged leukemia, lymphoma and solid tumors into one single variable including preoperative suspicion of a solid tumor not confirmed by pathological appraisal (benign or malignant).

#### eChapter 1.4: Functionality and geriatric examination

Functional impairments were assessed using the Barthel index and Instrumental Activities of Daily Living.  Patients were assessed for frailty (Fried's phenotype and Study of Osteoporotic Fractures [SOF]), falling incidences in the previous year and low gait speed in Timed-Up-and-Go test. Health-related quality of life was measured in the EQ5D-5L questionnaire.

For activities of daily living, any below-maximum score (Barthel index<100, IADL<8) was defined as functional impairment. Performance of more than 10 seconds in the Timed Up-and-go test (TUG) indicated gait slowing.

Frailty was assessed using two different scoring systems: Fried's frailty phenotype is defined slowness, weakness, weight loss, exhaustion and mobility(1). Slowness was defined by completion time >10s in the TUG. Weakness was assessed by low maximal hand grip strength adjusted for sex and BMI. Unintentional weight loss of ≥5% or ≥3 kg in the previous year was determined by patient's self-report. Self-reported exhaustion was assessed in the geriatric depression scale or the hospital anxiety and depression scale. Self-reported immobility was defined by the "inability to walk without difficulty" item from the EQ5D-5L and the Barthel index. Patients scoring ≥3, 1-2 or none of these items were categorized as frail, pre-frail and non-frail, respectively. SOF items were scored as weight loss ≥5% in the past year, inability to complete five consecutive chair rises, and answering “none or a little of the time” when asked for having a lot of energy. Patients scoring ≥2, 1 or none of these items were categorized as frail, pre-frail and non-frail, respectively(2).

HRQoL was assessed with the EQ5D-5L, because this questionnaire is a generic HRQoL instrument which can be applied on patients across indications and is therefore suitable for the heterogeneous cohort of BioCog patients. The EQ5D-5L is validated for many languages and a well-accepted instrument in the scientific health-economic community. The EQ5D-5L consists of 5 questions with 5 possible answer levels to describe the patient’s health state and a visual analogue scale (VAS), where a patient is to rate his/her overall well-being. Its completion takes between 2-3 minutes. Country specific value sets have been used to allocate validated index values to each health state(3-5).

#### eChapter 1.5: Nutrition and lifestyle factors

Data on the nutritional status were collected using the Body Mass Index and Short Form of the Mini Nutritional Assessment (MNA-SF). Alcohol consumption and drinking behavior was assessed with the Alcohol Use Disorder Identification Test. Patients were inquired for current smoking and lifetime tobacco consumption.

In the MNA-SF, 12-14 points indicate a normal nutritional status, whereas scores 0-7 and 8-11 points suggest malnutrition or the risk for malnutrition, respectively. Sex-specific scores of 5 of 40 points for women and 8 of 40 points for men were considered as cut-offs for alcohol dependency in the Alcohol Use Disorder Identification Test.

#### eChapter 1.6: Perioperative parameters

Type and duration of surgery and anesthesia, length of ICU and hospital stay were recorded. Postoperative pain and prescription of anticholinergic drugs were assessed for the delirium screening period. Further, time-adjusted analysis of postoperative pain and anticholinergic medication were run.

Pain scores were assessed at each visit for up to seven days using the Non-Visual Numeric Rating Scale (NVNRS), Behavioral Pain Scale (BPS) and the Critical-Care Pain Observation Tool (CPOT). Pain was defined as at least one positive score for at least one visit (NRS≥5/10, BPS≥6/12, CPOT≥3/8).

Duration of surgery was drawn from the electronic patient file for only one of the two study centers.

Since the data did not allow to determine the temporal sequence of ICU admission, ICU stay as well as hospital stay data were not analyzed as risk factors for POD.

In Berlin, end of anesthesia was defined as discharge of the patient from operating room, even if the patient was still intubated. In Utrecht, end of anesthesia was defined as extubation, even if the patient was extubated several days later on the ICU and the patient had received analgosedation.

The exact date of death was obtained for all patients who did not return for the follow-up assessment and could not be proven to be alive at 90 days after surgery from the medical records or due to personal contact. For this purpose, we obtained the exact date of deceasing from the patient files or the resident’s registration office. 90-days mortality was only obtained for patients recruited in Berlin.

#### eChapter 1.7: Neuropsychological testing

Neuropsychological testing was performed by trained study assistants in accordance with a standard operating procedure which was consented on with two neuropsychologists. Two independent assessors checked the data on plausibility by also considering free-text entries of research team members. When data for a participant was incomplete, missing values were imputed. If the data were missing due to impaired concentration or poor understanding of test instructions, missing data time point (N=42, 5%). When values were missing at random, e.g. due to technical difficulties or environmental disturbances, random forest imputation was applied to replace missing values for single cognitive test parameters (N=168, 18%). Data were not imputed when neuropsychological testing was missing completely (N=5, 1%). The missForest package for R Statistical Software was used for imputations(6).

The whole assessment comprised multiple cognitive domains:

SRT: The participant is shown a square on a computer screen and asked to respond to this stimulus by selecting a button as fast as possible.

PAL first trial memory score: Boxes were displayed on the screen and opened one at a time, in a randomized order. One or more of them will contain a pattern. The patterns shown in the boxes are then displayed in the middle of the screen, once at a time, and the subject must touch the box where the pattern was originally located. Each stage had ten attempts (trials) in total (the first presentation of all the shapes, then up to nine repeat presentations). If the subject made an error, the patterns were re-presented to remind the subject of their locations. When the subject got all the locations correct, they proceeded to the next stage. If the subject could not complete a stage correctly, the test terminated. First trial memory score was the measure was the number of patterns correctly located after the first trial, summed across the stages completed (range 0-26 in the clinical mode, with 26 meaning all the patterns were correctly located for all stages first time).

VRM delayed recognition: The participant was shown a list of 12 words once and asked to immediately recall freely as many of the presented words as possible. Twenty minutes after the word list presentation the participant had to correctly identify the initially presented words from a 24 words list containing 12 false distractors.

GPT for the dominant hand: The participant was asked to insert 25 pegs with a key alongside into wholes in a board as quickly as possible Key slots were rotated randomly, demanding visual-motor coordination skills and manual dexterity. Test parameter of interest was the task completion time using the dominant hand. Completion times of more than 300s were removed during plausibility checks in accordance with the testing manual.

TMT-B: The trail making task required a subject to connect a sequence of 25 consecutive targets on a sheet of paper. There were two parts to the test: in the first, the targets were all numbers (1, 2, 3, etc.) to connect sequentially; in the second part, numbers and letters (1, A, 2, B, etc.) had to be connected in alternating order. If the subject made an error, the test administrator corrected them before the subject continued the task. The completion time taken to complete the second part of the test, in which the subject alternated between numbers and letters, was used to examine executive functions.

To generate a dichotomous variable indicating preoperative cognitive impairment (PreCI) in the surgical cohort from the neurocognitive test battery, we recruited a non-surgical cohort of control participants to collect cognitive reference data: 114 non-surgical control participants with identical inclusion/exclusion criteria were recruited from outpatient clinics, primary care, elderly homes and via calls at public talks and invited to perform consecutive neuropsychological testing at baseline as well as after one week and three months. The control sample included 56 women (49%), the median age was 72 years (range 65-91 years) and the median MMSE was 29 points (range: 24-30 points). 33 (30%) participants received primary level and 44 (40%) received tertiary level education(7).

For the definition of PreCI, we used multiple cognitive test parameters moderate-to-good retest-reliability in the control group (intraclass coefficient between baseline and 3 months ≥0.75 based on a mean of multiple measurements, absolute-agreement, 2-way mixed-effects models)(7):

- mean correct latency from the Simple Reaction Time (SRT, reaction time),
- number of correctly remembered items in the free recall on the Verbal Recognition Memory test (VRM, free recall),
- number of correctly recognized items after delay on the VRM (delayed recognition memory),
- span length in the Spatial Span task (SSP, working memory),
- first trial memory score from the Paired Associate Learning test (PAL, visual memory),
- completion time of the Trail-Making-Test-B (TMT-B, executive functions), and
- completion time for the Grooved Pegboard test (GPT, fine motor skills).

Prior to calculation, SRT, GPT and TMT-B were log-transformed and sign-reversed to achieve an approximate normal distribution and a correspondence of higher scores with better cognitive performance. We calculated z-scores of the baseline measurement in each test parameter assessed in the control group. The same z-transformation was then applied to the surgical cohort. Z-scores <-1·96 in at least two cognitive test parameters or a compound z-score <-1·96 averaged over all z-scores was used to define preoperative cognitive impairment. eTable 3 gives an overview on the overlap of below threshold performance in different cognitive test parameters.

#### eChapter 1.8: Laboratory values

Blood parameters commonly determined in routine clinical care (albumin, γ-glutamyltransferase, uric acid, creatinine, lactate dehydrogenase, potassium, hemoglobin, glucose, glycated hemoglobin HbA1c, immature granulocytes, lymphocytes, mean corpuscular volume, mean platelet volume, NT-proBNP, neutrophils, eosinophils, eosinophils, reticulated platelets, thrombocytes, triglycerides, total cholesterol, low-density lipoprotein LDL, high-density lipoprotein HDL) were analysed by the laboratories adjacent to the study centers.

Aβ42, Aβ40, c-reactive protein (CRP), calprotectin, h-arginine, symmetric (SDMA) and asymmetric dimethylarginine (ADMA), troponin, malondialdehyde (MDA), nitrotyrosin, tryptophan, kynurenine, kynurenine acid, S100A12, interleukins [IL2, IL6, serum IL8, whole blood IL8, IL-10, IL18], oxidized LDL, intact proinsulin, c-peptide, leptin, soluble leptin receptor, total adiponectin and high and non-high molecular weight adiponectin were analyzed by parthners in the consortium (Immundiagnostik AG in Bernsheim, Germany, Institute of Protein Biochemistry, Consiglio Nazionale delle Ricerche (CNR) di Pisa in Italy, BIH Center for Regenerative therapies (BCRT), Charité – Universitätsmedizin Berlin and Berlin Institute of Health, Berlin, Germany).

Values below the detection limit of troponin, MDA, CRP and NT-proBNP have been replaced with a fixed value.

#### eChapter 1.9: Transcriptomics

For transcriptomics, blood was collected before anesthesia (day 0), next day (day +1) and after 3 months (day +90) in PAXgene blood RNA tubes (Qiagen). Total RNA, including RNA longer than approximately 18 nucleotides, was isolated by means of the PAXgene blood miRNA isolation kit (Qiagen) according to the manufacturer's instructions. Total RNA (RIN > 6·0) was processed and hybridized to microarrays according to Affymetrix' specifications. RNA amplification and microarray preparation were done by ATLAS Biolabs GmbH. 100ng of mRNA per fraction was amplified and loaded onto Affymetrix Clariom S human microarray plate for 96 samples (Thermo Fischer, Santa Clara, CA, USA) in accordance with manufacturer's recommendations. Hybridization, washing, staining and imaging took place in the GeneTitan™ Multi-Channel (MC) Instrument to provide the automated array processing. Both spike control oligos and hybridization control stages of the procedure were performed according to the manufacturer's instructions and under quality control. For microRNA analysis, 400ng of total RNA were labeled with biotin using Affymetrix® Flash Tag™ Biotin HSR. The arrays (miRNA 4.1 Array Plates) were hybridized, washed and scanned with the GeneTitan® MC Instrument. Both spike control oligos and hybridization control stages of the procedure were performed according to the manufacturer's instructions and under quality control. Raw data were normalized with the robust multi-array average method implemented in the Affymetrix Expression Console software. Further, all data were quality checked with R-package arrayQualityMetrics to assess the reproducibility, identify apparent outlier arrays and noise(8).

DNA was also prepared from buffy coats, but for sample size reasons, genotyping results are not included in this paper. For reasons of sample size, it is intended to pool genotype data with genome data from other research groups at a later time point.

#### eChapter 1.10: Neuroimaging

The imaging protocol comprised T1- and T2-weighted structural imaging sequences, high-resolution anatomical imaging of the hippocampus, arterial spin labeling, diffusion tensor imaging and resting-state combined fMRI/EEG.

##### eChapter 1.10.1: Image acquisition

In Berlin, data were collected at the Berlin Center for Advanced Neuroimaging using a 3T Magnetom Trio MR scanner (Siemens, München, Germany) with a 32-channel head coil. In Utrecht, data were collected with two Achieva 3T MRI scanner (Phillips, Amsterdam, Netherlands) equipped with a 8- and 32-channel head coil.

T1-weighted 3D structural brain scans were acquired using two similar MPRAGE sequences (magnetisation prepared rapid gradient echo, voxel size: 1x1 x1mm$^2$; Berlin: FOV=256x256x192 mm$^2$, TR=2500ms, TE=4·77ms, 7° flip angle; Utrecht: FOV=256x232x192mm$^2$, TR=7·9ms, TE=4·5ms, 8° flip angle).

High resolution imaging of the hippocampus was only acquired in Berlin with a TSE sequence designed for Siemens scanners (turbo spin echo with 0·4x0·4mm$^2$ voxel size in 24 slices at 2mm thickness FOV=150x150mm$^2$, TR=8020s, TE=48ms, echo train length=15).

Diffusion tensor images were acquired in 30 directions using EPI sequences (echo-planar imaging; Berlin: 2·5x2·5mm$^2$ voxel size in 50 slices at 2·5mm thickness, FOV=240x240mm$^2$, TR=6500ms, TE=100ms, $b_1$=0s/mm$^2$, $b_2$=1000s/mm$^2$, $b_3$=2500s/mm$^2$, 90° flip angle; Utrecht: 2·5x2·5mm$^2$ voxel size in 60 slices at 2·5mm thickness, FOV: 240v240mm$^2$, TR=7483ms, TE=100ms, $b_1$=0s/mm$^2$, $b_2$=1000s/mm$^2$, $b_3$=3000s/mm$^2$, 90° flip angle).

#### eChapter 1.10.2: Neuroimaging data processing

Imaging analyses were conducted by the Clinical Neuroscience Research Group, Dept. of Anesthesiology & Intensive Care Medicine, Charité – Universitätsmedizin Berlin (simultaneous fMRI/EEG, Nucleus Basalis Meynert), Pharmaimage Biomarker Solutions GmbH (volumetric analyses incl. High-Res Hippocampus), Utrecht (lesion analyses), Cambridge (diffusion tensor imaging).

##### eChapter 1.10.2.1: Cortical and subcortical gray matter

We used the FreeSurfer image analysis suite (version 6.0) to perform cortical segmentation. We quantified the variability of both subcortical volume estimates as well as cortical thickness in each of the parcels defined by the Desikan-Killiany (DK) surface-based cortical labeling protocol(9).

##### eChapter 1.10.2.2: Hippocampus

Hippocampal subfield volumes (cornu ammonis (CA)1, CA2/CA3 (sum of CA2 and CA3), dentate gyrus, subiculum, entorhinal cortex and parahippocampal gyrus) were quantified using the open-access Automated Segmentation of Hippocampal Subfields (ASHS) tool, which has been shown to achieve excellent agreement with manual segmentation and with intraclass correlations comparable to the overlap between human raters in manual segmentation(10, 11). The algorithm provides estimates by means of method multi-atlas segmentation, similarity-weighted voting, and a novel learning-based bias correction technique.

##### eChapter 1.10.2.3: Global brain volume and basal forebrain cholinergic system

SPM12 (Wellcome Centre for Human Neuroimaging, UCL, London, UK) in a MATLAB (Nattick, MA, USA) environment has been used to segment T1w images into partitions of gray and white matter as well as cerebrospinal fluid. Brain volume was calculated from the sum of voxels in the thresholded brain masks generated from summation of grey and white matter voxel masks.

The segmented grey and white matter images were further fed into DARTEL(12) to generate a BioCog-specific template. The procedure was repeated for a probabilistic atlas of the basal forebrain cholinergic system (BFCS)(13). The resulting DARTEL flow fields were used to label the basal forebrain of each patient and calculate the volume of the whole BFCS as well as the Nucleus basalis magnocellularis of Meynert corresponding to regions Ch4 and Ch4p (NBM). The method has been previously described(14).

##### eChapter 1.10.2.4: Diffusion Tensor Imaging

Diffusion weighted images were corrected for artifacts including denoised via MPPCA, Gibbs ringing removal eddy current and head motion correction, and bias-field correction(15-19). Fractional anisotropy (FA) and mean diffusivity (MD) maps were calculated with weighted least-squares tensor fitting(20).

To parcellate whit matter, we used TractSeg which segments tracts using a fully convolutional neural network (FCNN) utilizing fields of fiber orientation distribution factors (fODF) peaks. TractSeg takes as input the three principal fiber directions per voxel, adding up to nine input channels -three per principal direction. The principal directions were extracted using the multi-shell multi-tissue constrained spherical deconvolution (CSD) and peak extraction available in MRtrix(19, 20). A 2D encoder-decoder FCNN then produces one tract probability image for each orientation (coronal, axial, sagittal) and for each tract. The 2D encoder-decoder FCNN architecture was inspired by the U-Net encoder-decoder architecture previously proposed(21). The tract probability images from the three orientations are then concatenated in the channel dimension resulting in a 3D image. The output was binarised (thresholded 0·5 and binarised) to create discrete distinctions between the particular fiber tract regions or something else. The approach enables for multi-label segmentation with several tracts sharing one voxel. This is used as input for a second FCNN which runs three times. The three outputs per tract from the second FCNN are merged using the mean to generate the final segmentation. The final segmentation is a 72-channel image, wherein each channel contains the voxel probabilities for one tract. Reference binary segmentation for 72 major white matter tracts for each subject were generated semi-automatically.

Those reference segmentations are used as labels for training and validating our network(22). ROI fractional anisotropy (FA), mean diffusivity (MD) and kurtosis (MK) maps were calculated and mean and standard deviation (std) were computed within binary masks generated from the detected tracts. ROI FA/MD weighted mean/std take into account this probabilistic information. That is, voxels that have a lower probability to belong to the specific fiber tract are weighted accordingly to contribute less to the mean/std.

##### eChapter 1.10.3: Structured database

All clinical data were entered in the electronic clinical case report form (eCRF) SecuTrial® (interActive Systems, Berlin, Germany). All clinical data as well as the collected neuropsychological test data, lab values and neuroimaging data underwent extensive quality and plausibility checks and then transferred to our open source data management system (XNAT 1.7.4 https://www.xnat.org/) which was implemented, hosted and structured, including automatic data processing pipelines by Pharmaimage Biomarker Solutions GmbH. Management of -omics data was conducted at the Bioinformatics core, Luxembourg Centre for Systems Biomedicine (LCSB), University of Luxembourg.

#### eChapter 1.11: Statistical analyses

##### eChapter 1.11.1: Single variable analyses

Single variable analyses were conducted at the Dept. of Department of Anesthesiology and Operative Intensive Care Medicine together with Institute of Biometry and Clinical Epidemiology (both Charité – Universitätsmedizin Berlin).

Supplementary result tables contain the number of available datasets for each variable, the number of missing values in patients with POD or POCD as well as in patients without the respective postoperative impairment. Tables state mean values with standard deviations in the compared groups for continuous variables. For categorical variables, the absolute number of cases as well as the relative frequency of a predictor characteristic in the group with POD or POCD and the group without postoperative cognitive impairment are shown.

Continuously scaled clinical variables were categorized according to clinically relevant cut-off values for presentation of interpretable ORs. In the case of duration of anesthesia, dichotomization had the additional purpose to level out different recording practices in the two study centers.

To assess perioperative changes in blood-based parameters, we calculated the difference between postoperative and preoperative parameter levels. For presentation purposes, laboratory variables were standardized prior to simple logistic regression. Whenever a blood-based parameter has been measured in more than one laboratory or with different kits, the parameter has been adjusted for laboratory site by regression and saved as standardized variables. No other transformations have been applied.

In the analysis of neuroimaging results, neither adjustments were made for global brain volume nor for the MRI scanner, since analysis of “travelling brains” suggested acceptable between-scanner agreement for the measures presented in this manuscript.

##### eChapter 1.11.1.1: Supplementary analysis of postoperative pain and anticholinergic medication

For postoperative pain and anticholinergic medication, two types of analyses were conducted. In a global approach, we analyzed the general association of any prescription of anticholinergic medication during the postoperative period or the occurrence of any therapy-demanding postoperative pain exacerbation with POD during the screening period. To account for the assumed causal association of pain or medication with delirium, we conducted a time-adjusted analysis. We thus calculated the incidence of postoperative pain and anticholinergic prescriptions from the day of surgery until postoperative day 1, 2, 3, etc. Analogously, we calculated delirium incidence after the day of surgery, postoperative day 1, 2, 3, etc. We then analyzed associations of postoperative pain and anticholinergic medication between day of surgery and postoperative day X with delirium incidence from postoperative day X+1 and discharge or postoperative day 7, e.g. postoperative pain until postoperative day 2 was considered to be associated with delirium on postoperative day 3 or later.

##### eChapter 1.11.1.2: Treatment of continuously scaled variables for single variable analysis

Clinical variables and scores were transformed into dichotomous or ordinal variables based on clinicians' recommendation. For GDS and CCI, patients with a score of at least 1 were compared to patients with a score of 0, which may be interpreted as "having at least one symptom of depression" and "having at least one comorbidity limiting expectancy". For the BMI, the categories underweight (<18·5kg/m²), ideal weight (18·5-24·9kg/m²), overweight (25-29·9kg/m²) and obesity were used (≥30kg/m²). To consider a non-linear association of BMI and POD/POCD risk, we also compared obesity with normal weight to overweight (excluding all patients with underweight) and underweight with normal weight to overweight (excluding all patients with obesity). Low MMSE was defined as a score of 24-26 points. MNA and AUDIT have been categorized according to recommended cut-off values. To level out differences in the recording practices between the two study centers, anesthesia duration was dichotomized at 4h to ameliorate the effects of outliers. Instead of duration of ICU stay, ICU admission independent of duration has been analyzed. Neuroimaging variables significantly associated with POD were normalized to a standard deviation of 1. Laboratory variables were either normalized or analyzed as standardized residuals after adjustment for laboratory procedures. Some continuous variables had been dichotomized during the validation process prior to database entry (Barthel, IADL, postoperative anticholinergic medication score according to Carnahan)

#### eChapter 1.11.2: Machine learning approach

Machine learning analyses were conducted by Pharmaimage Biomarker Solutions GmbH (Berlin) with support from Adalab UG (Hamburg) and the Hasso-Plattner Institute (Potsdam). The multiple-predictor methods were programmed in Python, using the GBT implementation of the XGBoost library (https://xgboost.readthedocs.io/en/stable/#).

##### eChapter 1.11.2.1: Performance evaluation using nested k-fold cross-validation

After training the parameters of a machine learning model, the performance needs to be evaluated on a separate dataset than the one that the model was trained on. This way one can test whether or not the model performance generalizes and does not simply memorize the training dataset. This is done by separating the dataset into a train-validation split. To account for the sampling bias that occurs when randomly assigning samples of the dataset into both splits, we generated k different train-validation splits. The evaluation scores on these k splits are then averaged. This procedure is called *k-fold cross-validation*.

Next to the learned parameters of the model (e.g. linear weights in a linear regression), ML models oftentimes require further pre-settings called hyperparameters. When working with GBT we need to specify beforehand how many decision trees are combined. Hyperparameters like this one should also generalize to unseen data. Hence, we apply k-fold cross-validation also on this outer layer of evaluation.

In summary, this procedure is called *nested cross-validation* as we split our pre-processed data into *k_outer* development-testing splits and each development split into *k_inner* train-validation splits. Nested cross-validation enables us to optimize model parameters in an inner loop and hyperparameters in an outer loop. This process yields k_outer final models for which we compute mean and confidence intervals of the evaluation metrics. We use *k_outer=1*0, *k_inner*=5, tune the model per *k_inner* split for 10,000 training iterations, and stratify with regard to POD and non-POD cases.

**eChapter 1.11.2.2: Precision, recall, specificity, & sensitivity**

When evaluating a model by means of precision and recall or specificity and sensitivity, one generally has to choose a trade-off between the instances of each of those metric pairs. Precision is defined as $\frac{Truepositives}{Truepositives+Falsepositives}$ which captures how many of those cases that a model deems POD-positive are actually POD-positive. Recall is defined as $\frac{Truepositives}{Truepositives+Falsenegatives}$ and semantically shows how many of those who are really POD-positive are correctly recognized as such by the model. Hence the Recall is identical to the sensitivity metric. Finally, the definition of specificity is $\frac{Truenegatives}{Truenegatives+Falsepositives}$ and it shows how many of the cases that are classified as POD-negative are actually negative. The precision metric is not defined over the number of negative cases. This makes it less susceptible to an underrepresentation of the positive target class, as is the case in our POD dataset. As a consequence, precision is focused on capturing the classifier’s capabilities to model the positive class and retrieve information on it.

As the output of a machine learning model is usually a continuous random variable, we defined a threshold that determines which output value is considered positive or negative in a binary fashion. The above-mentioned metrics, hence, depend on how we choose to set this threshold. We automatically selected the threshold for which the sum of precision and recall is highest. It is possible to adjust it in order to determine a specified value on a target metric. However, this comes with the trade-off of changing all other metrics implicitly and needs to be selected based on expert considerations.

In evaluating our model for POD prediction, we focus on two main metrics: Receiver-operating characteristic-area under the curve (ROC AUC) measures the model's ability to differentiate between POD and non-POD cases, providing an overview of performance across various thresholds. In addition, precision-recall curve (PR) is essential for our imbalanced dataset. It assesses the trade-off between precision (true positives out of positive predictions) and recall (correct identification of actual POD cases). The area under this curve (average precision, AP) summarizes the model's performance. The Brier score B was calculated to assess model calibration(23).

**eChapter 1.11.2.3: Data processing**

The dataset comprises a detailed enumeration of cases and features across various data types, summarized in eTable 1. For an in-depth analysis of each parameter, refer to the single variable analyses in eTables 4-15.

In the assessment of the 'Timed Up & Go' test, a novel approach was adopted to account for missing data. A dedicated 'missing value feature' was introduced, acknowledging that non-completion of the test might indicate a patient's inability to perform the task due to health constraints.

In analyzing RNA features, the focus was solely on gene expression profiles. Control measurements were excluded to refine the dataset for more targeted analysis.

Pain levels at two critical points were considered: on the day of the operation and the first day following the operation. This approach aids in understanding the acute pain trajectory in the perioperative period.

The analysis of neuroimaging data was streamlined by reducing the initial set of 1083 features to two pivotal variables: Total Brain Volume and Nucleus Basalis Meynert (NBM) Volume. These parameters are widely recognized as indicators of neurodegeneration in (incipient) dementia. Further inclusion of imaging variables did not yield significant improvements in model performance, as evidenced in our experimental results.

The study included only perioperative features that were consistently measured across both datasets. Patient selection was restricted to those with complete data at both timepoints (pre- and post-operation). The perioperative feature analysis is based on the difference in measurements between these two critical timepoints.

**eTABLE 1: NUMBER OF CASES AND FEATURES PER DATATYPE**

| Data | Cases (N) | Features (N) |
|---|---|---|
| **Clinical** | 929 | 31 |
| **Blood** | 813 | 69 |
| **Blood Periop.** | 813 | 67 |
| **Imaging** | 478 | 2 |
| **Precipitant** | 929 | 3 |
| **RNA** | 650 | 20893 |
| **RNA Periop.** | 377 | 20893 |
| **Micro RNA** | 719 | 36353 |
| **Micro RNA Periop.** | 448 | 36353 |
| **Pain Periop.** | 883 | 2 |

##### eChapter 1.11.2.4: Training setup

For constructing our predictive models, we utilized the XGBoost library (version 2.0.2) to train gradient boosted trees (XGBClassifier). A nested k-fold validation method was employed to fine-tune and assess the hyperparameters and models, thereby minimizing the risk of overfitting. The data was partitioned into two segments: 10% reserved for testing and 90% for development. Within the development segment, 20% was further allocated to validation. This setup ensured rigorous validation through an outer loop (10-fold) and an inner loop (5-fold).

Separate analyses were conducted across different data sources: clinical/neuropsychological, blood-based, neuroimaging, and transcriptomic. Additionally, models combining these data sources were also evaluated. In each domain, we explored models both with and without the inclusion of precipitating factors, such as the duration of anesthesia, site of surgery, uncontrolled postoperative pain, and postoperative anticholinergic medication. Blood-based data collected on the first postoperative day were also incorporated. Each model was initially developed and assessed within its specific domain before proceeding to multi-domain aggregations. Given the extensive nature of RNA data (over 20,000 features) and its limited availability (smaller patient subset), a specialized approach was adopted. Models pertaining to RNA data were exclusively evaluated using the patient subset for whom RNA data was available. For all other data types and models excluding RNA data, the full cohort of 929 patients was utilized. In cases of missing values, imputation techniques, as previously described, were applied to ensure data completeness and integrity.

**eChapter 1.11.2.5: Hyperparameter tuning process**

GBT models are inherently dependent on several key hyperparameters, including learning rate, maximum tree depth, and the number of estimators. To optimize these parameters, we defined specific value ranges based on plausible expectations, establishing a search space anticipated to contain the optimal settings for our objectives. The tuning process was automated, treating hyperparameter optimization as a search problem. This approach leverages an optimization algorithm to systematically identify the most effective hyperparameter configuration. The configuration is based on a predefined quality metric—ROC AUC (Receiver Operating Characteristic Area Under the Curve) for Post-Operative Delirium (POD) classification on a validation dataset.

Evaluation and Optimization Framework: In our methodology, we meticulously evaluated 50 distinct hyperparameter configurations for each outer fold within the nested k-fold cross-validation framework. This evaluation was facilitated by the Optuna framework (version 2.9.1), renowned for its implementation of efficient Bayesian optimization techniques. For each outer fold, Optuna generates a set of hyperparameters, which is then assessed using the validation sets of the corresponding inner folds. This assessment primarily focuses on the average Area Under the Curve (AUC) of the Receiver Operating Characteristic (ROC) derived from these inner folds. The hyperparameter set demonstrating the highest performance on the inner fold validations—as gauged by AUC—is selected as the optimal configuration. This chosen set is subsequently employed to train the model on the entire dataset corresponding to the respective outer fold.

This rigorous approach ensures the reliability and robustness of our model by optimizing it based on comprehensive cross-validation, a critical aspect in the context of high-stakes clinical decision-making. It is important to note that the final hyperparameters varied across folds, aligning with the aim of finding the most suitable configuration for each dataset. This approach not only enhances the model's accuracy but also ensures a robust comparison across different trials.

Predefined Ranges for Tunable Hyperparameters: The hyperparameter tuning involved the following ranges:

- Maximum depth: integer range: 3-10
- Learning rate: floating range 0·005-0·1 on a logarithmic scale
- Number of estimators: integer range 5-100
- Subsample: floating range from 0·8-1·0
- Column sample by tree: floating range 0·6-1·0
- Γ: floating range 0·0-5·0

These ranges were carefully selected to encompass a wide spectrum of potential configurations, thereby ensuring comprehensive exploration of the hyperparameter space.

**eChapter 1.11.2.6: Performance evaluation using nested k-fold cross-validation**

We employed nested k-fold cross-validation for a robust evaluation of our GBT model's performance. This method is pivotal in confirming the model's generalizability, ensuring its effectiveness extends beyond the training dataset. The dataset was divided into 'k' splits to reduce sampling bias, following standard k-fold cross-validation procedures, and underwent 'k_outer' splits for development-testing, each further segmented into 'k_inner' train-validation splits. This nested format allows for simultaneous tuning of model parameters (inner loop) and hyperparameters (outer loop). We set 'k_outer' at 10 and 'k_inner' at 5, with each outer split undergoing 50 training iterations. Stratification was based on POD case presence. The nested approach yielded 'k_outer' models, for which we calculated mean scores and confidence intervals for key metrics, ensuring an in-depth assessment of performance across data subsets.

This methodological approach underpins the model's reliability and adaptability to different clinical settings.

### eChapter 2: Supplementary Results

#### eChapter 2.1: Description of excluded patients

##### eChapter 2.1.1: Utrecht

Of 1013 patients who refused to participate, 304 (30%) were not interested in research participation, 381 (38%) found that the research procedures were to stressfull and 328 (32%) mentioned that the tests took up too much time. Of 368 patients who violated inclusion/exclusion criteria, 169 (46%) did not meet inclusion criteria, 13 (4%) were excluded due to an MMSE score <24 points, 135 (37%) were not eligible for MRI and 51 (14%) participated in another study. Of 607 patients who were excluded for other reasons, 63 (10%) could not attend the assessments due to inavailability of transportation facilities, and no specific reason was recorded in 554 (90%) of these patients.

##### eChapter 2.1.2: Berlin

No individual reasons were recorded for 1795 patients who refused to participate. Of 1448 patients who did not fulfill inclusion/exclusion criteria, 355 (25%) were expected to have surgery less than 60min duration, 14 (1%) were scheduled for surgery in local anesthesia, 19 (1%) were younger than 65 years, 292 (21%) were not eligible for MRI (226 [16%] had contraindications and 66 (5%) were not eligible for transportation to the MRI faculty), 51 (4%) had a diagnosis of dementia (21, 2%) or an MMSE score <24 points (30, 2%), 205 (14%) patients had conditions interfering with cognitive testing (66 [5%] had impaired vision or hearing, 119 [8%] did not speak German, 20 [1%] had severe neuropsychiatric illness), 160 (11%) were not able to give informed consent (18 [1%] had a legal attendant, 123 [8.5%]had severe impairment of speech, 19 [1%] due to other reasons), 76 (5%) participated in another trial, 276 (19%) were expected not to attend follow-up assessments since they were either outpatients (34, 2%) or due to significant general health detriment (242, 17%). Of 1304 patients who were excluded for other reasons, 27 (2%) were excluded because time until surgery was not sufficient for a complete assessment, since surgery was canceled (29, 2%) or scheduled for another hospital (5, <1%). In 63 (5%) cases, there was no MRI capacity available. 325 (25%) patients could not be contacted and 855 patients were not included for organisational or unspecified reasons (66%).

### eChapter 2.2: Overview on missing data

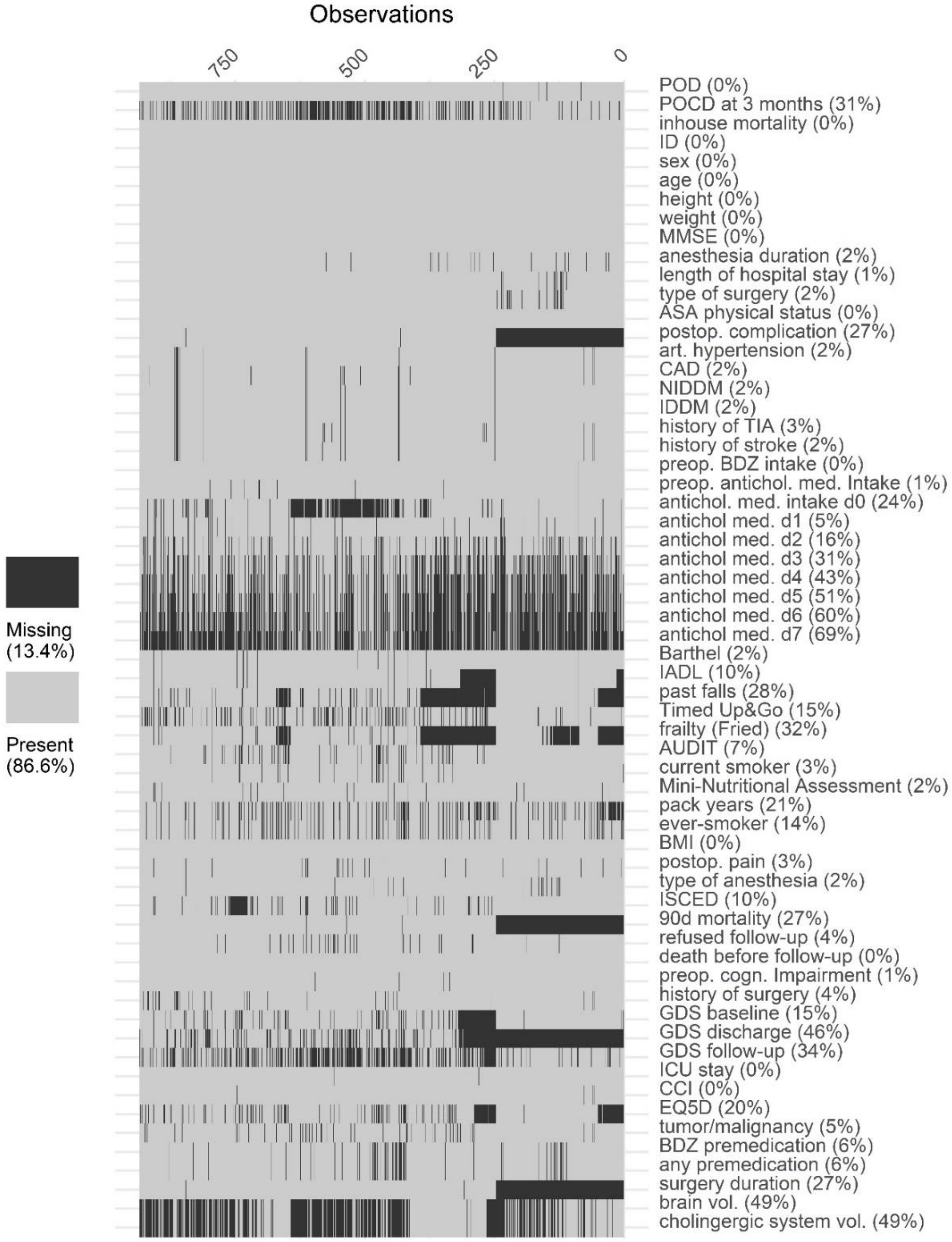


**EFIGURE 1: SUMMARY OF MISSING CLINICAL DATA.**

Abbreviations: POD: postoperative delirium, POCD: postoperative cognitive dysfunction, ASA: American Society of Anesthesiologists, CAD: Coronary artery disease, (N)IDDM: (non-) insulin dependent diabetes mellitus, TIA: transient ischemic attack, preop.: preoperative, BDZ: benzodiazepine, antichol. med.: anticholinergic medication, d: day, IADL: instrumental activities of daily living, AUDIT: alcohol use disorder identification test

Abbreviations: POD: postoperative delirium, POCD: postoperative cognitive dysfunction, ASA: American Society of Anesthesiologists, CAD: Coronary artery disease, (N)IDDM: (non-) insulin dependent diabetes mellitus, TIA: transient ischemic attack, preop.: preoperative, BDZ: benzodiazepine, antichol. med.: anticholinergic medication, d: day, IADL: instrumental activities of daily living, AUDIT: alcohol use disorder identification test

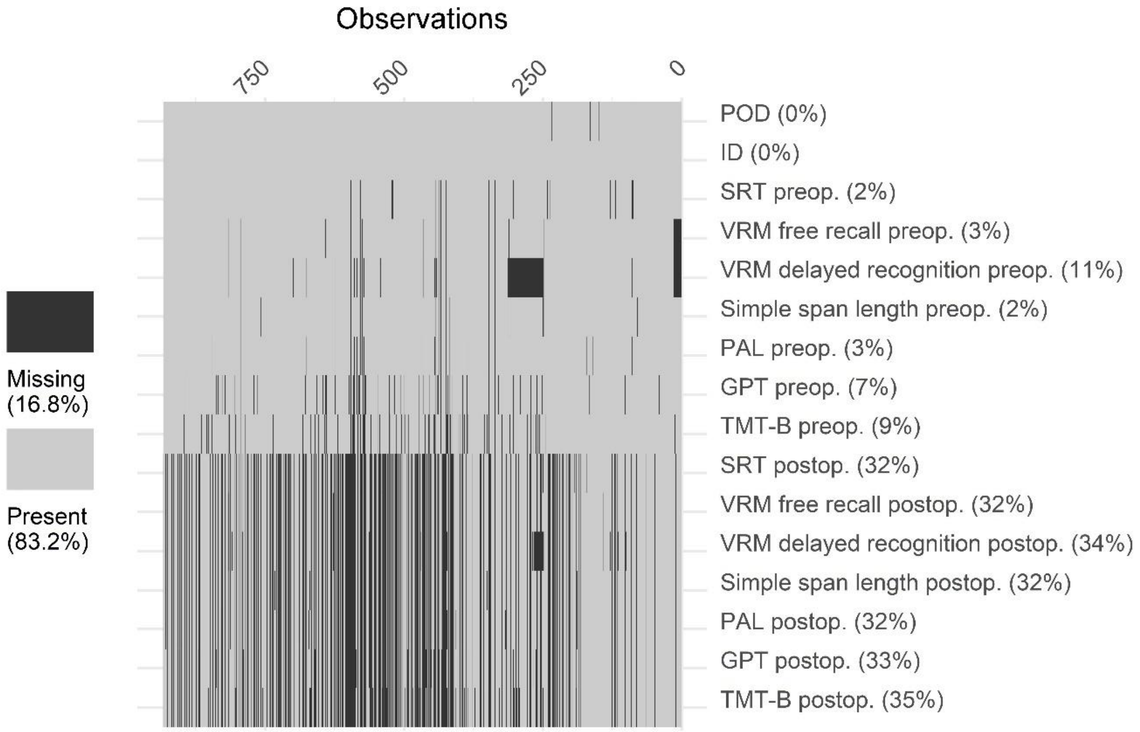


**eFigure 2: Missing data in the neuropsychological testing.**

Abbreviations: POD: postoperative delirium; SRT: Simple reaction time: VRM: Verbal recognition memory; PAL: Paired associate learning; GPT: Grooved pegboard test; TMT-B: Trail-making test: part B; preop: preoperative; postop: postoperative.

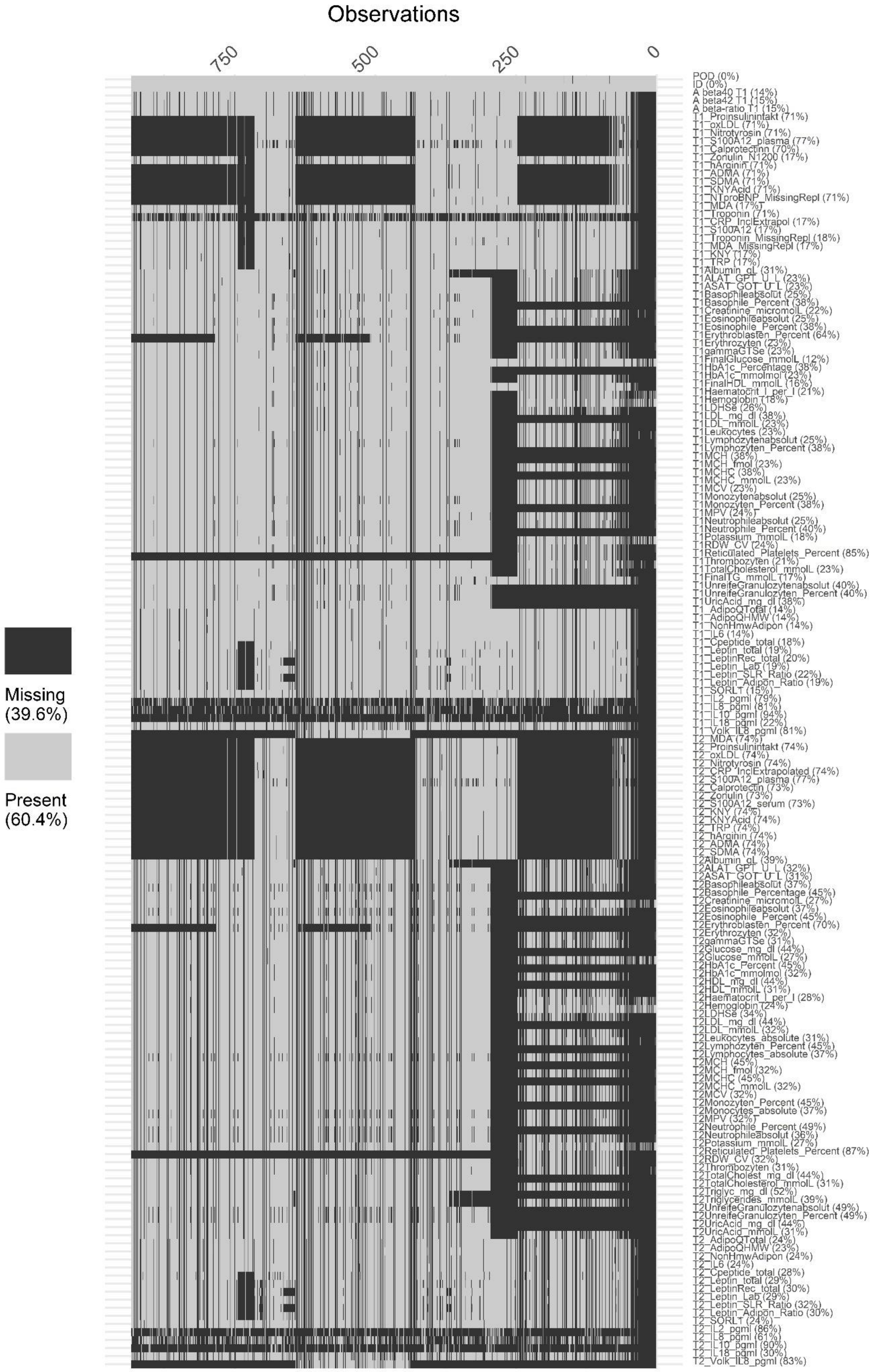


**eFigure 3: Overview on missing blood-based parameters.**

**eTable 2: Overlap of patients performing below the z<-1·96 threshold between different cognitive test parameters.**

The number of patients below the threshold is given on the diagonal as an absolute number (N). The number of patients scoring lower than z<-1·96 in two different cognitive test parameters is also given as an absolute number (N), with the associated $\chi^2$-test statistic (including degrees of freedom) and p-value. PAL: Paired Associate Learning test, VRM: Verbal Recognition Memory Test, FR: free recall, Rec: recognition memory, SSP: Simple Span length, GPT: Grooved Pegboard Test, SRT: Simple Reaction Time, TMT-B: Trail-Making-Test part B, PreCI: preoperative cognitive impairment.

| | PAL | VRM-FR | VRM-Rec | SSP | GPT | SRT | TMT-B | compound | PreCI |
|---|---|---|---|---|---|---|---|---|---|
| **PAL** | N=106 | N=14 $\chi^2$=37·8 (1) p<0·001 | N=17 $\chi^2$=28·6 (1) p<0·001 | N=29 $\chi^2$=16·3 (1) p<0·001 | N=25 $\chi^2$=30·4 (1) p<0·001 | N=12 $\chi^2$=18·6 () p<0·001 | N=28 $\chi^2$=59·2 (1) p<0·001 | N=45 $\chi^2$=155·8 (1) p<0·001 | N=65 $\chi^2$=241·9 (1) p<0·001 |
| **VRM FR** | | N=30 | N=12 $\chi^2$=76·3 (1) p<0·001 | N=9 $\chi^2$=6·1 (1) p=0·013 | N=4 $\chi^2$=0·7 (1) p=0·41 | N=4 $\chi^2$=7·8 (1) p=0·005 | N=13 $\chi^2$=55·6 (1) p<0·001 | N=18 $\chi^2$=94·4 (1) p<0·001 | N=24 $\chi^2$=120·7 (1) p<0·001 |
| **VRM-Rec** | | | N=48 | N=14 $\chi^2$=9·0 (1) p=0·003 | N=8 $\chi^2$=3·5 (1) p=0·061 | N=7 $\chi^2$=16·2 (1) p<0·001 | N=18 $\chi^2$=63·4 (1) p<0·001 | N=26 $\chi^2$=120·7 (1) p<0·001 | N=33 $\chi^2$=136·3 (1) p<0·001 |
| **SSP** | | | | N=133 | N=27 $\chi^2$=23·6 (1) p<0·001 | N=12 $\chi^2$=11·7 (1) p=0·001 | N=20 $\chi^2$=11·8 (1) p=0·001 | N=39 $\chi^2$=73·7 (1) p<0·001 | N=57 $\chi^2$=119·2 (1) p<0·001 |
| **GPT** | | | | | N=84 | N=10 $\chi^2$=16·7 (1) p<0·001 | N=29 $\chi^2$=93·8 (1) p<0·001 | N=42 $\chi^2$=181·3 (1) p<0·001 | N=53 $\chi^2$=200·7 (1) p<0·001 |
| **SRT** | | | | | | N=84 | N=7 $\chi^2$=7·8 (1) p=0·005 | N=17 $\chi^2$=66·4 (1) p<0·001 | N=22 $\chi^2$=78·3 (1) p<0·001 |
| **TMT-B** | | | | | | | N=71 | N=50 $\chi^2$=340·3 (1) p<0·001 | N=59 $\chi^2$=327·9 (1) p<0·001 |
| **compound** | | | | | | | | N=86 | N=86 $\chi^2$= 623·4 (1) p<0·001 |
| **PreCI** | | | | | | | | | N=122 |

eTable 3: Number (relative fraction) of delirious patients identified by bedside screening and chart review.

| Postoperative day | Positive in bedside screening (N [%]) | Positive in chart review (N [%]) | Positive in chart review and at bed-side (N [%]) |
|---|---|---|---|
| **0** | 109 (100%) | n. a. | n. a. |
| **1** | 28 (42%) | 24 (36%) | 15 (22%) |
| **2** | 21 (44%) | 10 (21%) | 17 (35%) |
| **3** | 15 (30%) | 15 (30%) | 20 (40%) |
| **4** | 10 (25%) | 7 (18%) | 23 (58%) |
| **5** | 8 (25%) | 8 (25%) | 16 (50%) |
| **6** | 9 (30%) | 10 (33%) | 11 (37%) |
| **7** | 8 (14%) | 35 (63%) | 13 (23%) |

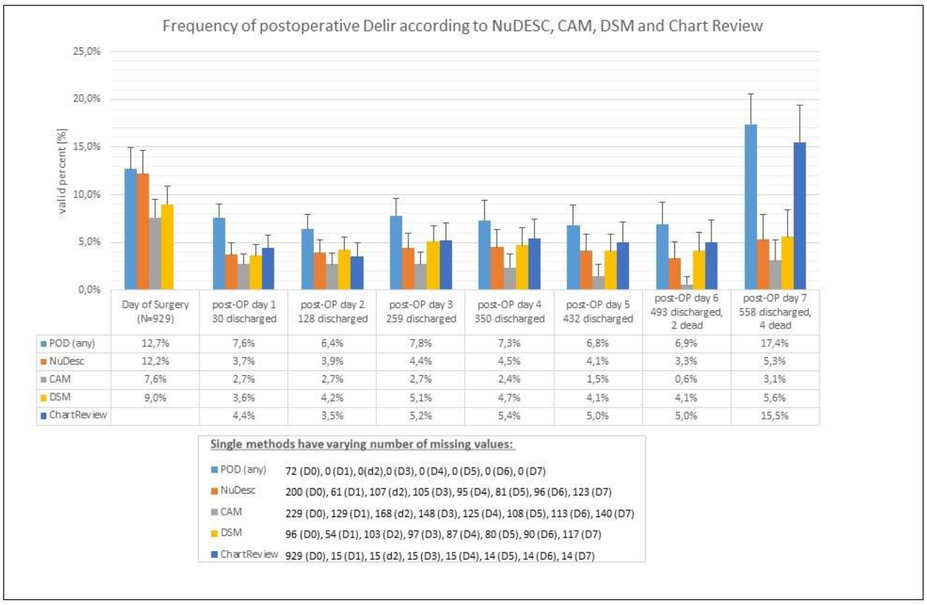


| | Day of Surgery (N=929) | post-OP day 1 30 discharged | post-OP day 2 128 discharged | post-OP day 3 259 discharged | post-OP day 4 350 discharged | post-OP day 5 432 discharged | post-OP day 6 493 discharged, 2 dead | post-OP day 7 558 discharged, 4 dead |
|---|---|---|---|---|---|---|---|---|
| POD (any) | 12,7% | 7,6% | 6,4% | 7,8% | 7,3% | 6,8% | 6,9% | 17,4% |
| NuDesc | 12,2% | 3,7% | 3,9% | 4,4% | 4,5% | 4,1% | 3,3% | 5,3% |
| CAM | 7,6% | 2,7% | 2,7% | 2,7% | 2,4% | 1,5% | 0,6% | 3,1% |
| DSM | 9,0% | 3,6% | 4,2% | 5,1% | 4,7% | 4,1% | 4,1% | 5,6% |
| ChartReview | | 4,4% | 3,5% | 5,2% | 5,4% | 5,0% | 5,0% | 15,5% |

**Single methods have varying number of missing values:**

- POD (any) 72 (D0), 0 (D1), 0(d2),0 (D3), 0 (D4), 0 (D5), 0 (D6), 0 (D7)
- NuDesc 200 (D0), 61 (D1), 107 (d2), 105 (D3), 95 (D4), 81 (D5), 96 (D6), 123 (D7)
- CAM 229 (D0), 129 (D1), 168 (d2), 148 (D3), 125 (D4), 108 (D5), 113 (D6), 140 (D7)
- DSM 96 (D0), 54 (D1), 103 (D2), 97 (D3), 87 (D4), 80 (D5), 90 (D6), 117 (D7)
- ChartReview 929 (D0), 15 (D1), 15 (d2), 15 (D3), 15 (D4), 14 (D5), 14 (D6), 14 (D7)

**eFigure 4: Frequency of POD according to NuDesc, CAM, DSM and chart review.**

The missing values for the day of surgery (d0) each postoperative day (d1-d7) are indicated for the complete screening period.

**ETABLE 4: SOCIODEMOGRAPHIC PARAMETERS IN 745 PATIENTS WITHOUT AND 184 PATIENTS WITH POD.**

Abbreviations: ISCED: International Standard Classification of Education. Data are given number of included patients (N), missing data for patients without/with POD (nNPOD/nPOD), mean ± standard deviation or the absolute (relative) number of patients with a predictive characteristic among patients without/with POD. Odds ratios (OR) with 95% confidence intervals (CI).

| | | N | Missing $n_{NPOD}/n_{POD}$ | No POD | POD | OR (95% CI) |
|---|---|---|---|---|---|---|
| **Age (y)** | | 929 | 0/0 | 72±5 | 73·8±5 | *1·07 (1·04; 1·10)* |
| **Sex (reference: male)** | | 929 | 0/0 | 309 (41%) | 85 (46%) | 1·21 (0·87; 1·68) |
| **ISCED (reference: level 5-6)** | **1-2** | 839 | 71/19 | 121 (18%) | 29 (18%) | 1·07 (0·84; 1·35) |
| | **3-4** | | | 279 (41%) | 62 (39%) | |

##### eTABLE 5: COMORBIDITIES IN 745 PATIENTS WITHOUT AND 184 PATIENTS WITH POD.

Abbreviations CCI: Charlson Comorbidity Index, ASA-PS: American Society of Anesthesiologists Physical Status, TIA: transient ischemic attack, NIDDM: non-insulin dependent diabetes mellitus, IDDM: insulin dependent diabetes mellitus, GDS: geriatric depression scale. Data are given number of included patients (N), missing data for patients without/with POD (nNPOD/nPOD), mean ± standard deviation or the absolute (relative) number of patients with a predictive characteristic among patients without/with POD. Odds ratios (OR) with 95% confidence intervals (CI). * The reference groups includes patients without the respective condition.

| | N | Missing $n_{NPOD}/n_{POD}$ | No POD | POD | OR (95% CI) |
|---|---|---|---|---|---|
| **CCI ≥1p (ref.: CCI=0)** | 925 | 4/0 | 443 (60%) | 137 (74%) | *1·96 (1·37; 2·84)* |
| **ASA-PS III-IV (ref.: ASA-PS I-II)** | 929 | 0/0 | 236 (32%) | 100 (54%) | *2·57 (1·85; 3·57)* |
| **Arterial Hypertension*** | 915 | 12/2 | 462 (63%) | 123 (68%) | 1·22 (0·87; 1·73) |
| **Coronary artery disease*** | 906 | 18/5 | 132 (18%) | 49 (27%) | *1·70 (1·70; 2·47)* |
| **Anemia*** | 764 | 142/23 | 197 (33%) | 77 (49%) | *1·89 (1·33; 2·69)* |
| **Diabetes mellitus*** | 915 | 11/3 | 155 (21%) | 48 (27%) | 1·35 (0·93; 1·96) |
| **NIDDM*** | 915 | 11/3 | 93 (13%) | 30 (17%) | 1·37 (0·88; 2·14) |
| **IDDM*** | 915 | 11/3 | 62 (8%) | 18 (10%) | 1·20 (0·69; 2·08) |
| **History of TIA*** | 904 | 20/5 | 25 (3%) | 8 (4%) | 1·31 (0·54; 2·83) |
| **History of stroke*** | 910 | 15/4 | 41 (6%) | 13 (7%) | 1·31 (0·66; 2·43) |
| **Preoperative tumor, lymphoma or leukemia*** | 879 | 36/14 | 206 (30%) | 85 (50%) | *2·44 (1·74; 3·44)* |
| **History of at least one past surgery*** | 896 | 28/5 | 668 (97%) | 167 (93%) | 1·02 (0·55; 2·05) |
| **GDS ≥1p (ref.: GDS=0)** | 792 | 117/24 | 441 (70%) | 126 (79%) | *1·57 (1·05; 2·41)* |

eTABLE 6: PREOPERATIVE MEDICATION IN 745 PATIENTS WITHOUT AND 184 PATIENTS WITH POD.

Data are given number of included patients (N), missing data for patients without/with POD (nNPOD/nPOD), mean ± standard deviation or the absolute (relative) number of patients with a predictive characteristic among patients without/with POD. Odds ratios (OR) with 95% confidence intervals (CI). Patients without respective medication were defined as the reference group.

| | N | Missing $n_{NPOD}/n_{POD}$ | No POD | POD | OR (95% CI) |
|---|---|---|---|---|---|
| **Longterm benzodiazepine medication** | 928 | 1/0 | 113 (15%) | 60 (34%) | *2·70 (1·87; 3·89)* |
| **Preoperative longterm anticholinergic medication** | 919 | 10/0 | 185 (25%) | 54 (29%) | 1·23 (0·86; 1·76) |
| **Premedication** | 874 | 37/18 | 86 (12%) | 25 (15%) | 1·28 (0·78; 2·05) |
| **Benzodiazepine premedication** | 874 | 37/18 | 83 (12%) | 25 (15%) | 1·34 (0·82; 2·16) |

**eTable 7: Functionality and preoperative cognitive status in 745 patients without and 184 patients with POD.**

Abbreviations: IADL: Instrumental activities of daily living, MMSE: Mini-Mental Status Examination, GPT: Grooved Pegboard Test, VRM: Verbal Recognition Memory, SOF: Study of Osteoporotic Fracture, SRT: Simple Reaction Time (mean latency), SSP: simple span length, TMT-B: Trail-Making-Test part B, PAL: Paired Associate Learning (memory score), TUG: Timed Up-and-go-test. Data are given number of included patients (N), missing data for patients without/with POD (nNPOD/nPOD), mean ± standard deviation or the absolute (relative) number of patients with a predictive characteristic among patients without/with POD. Odds ratios (OR) with 95% confidence intervals (CI).

| | | N | Missing $n_{NPOD}/n_{POD}$ | No POD | POD | OR (95% CI) |
|---|---|---|---|---|---|---|
| Functional Impairment (Barthel or IADL) (ref.: Barthel score=100 and IADL score=8) | | 849 | 74/6 | 111 (28%) | 67 (38%) | *1·59 (1·12; 2·24)* |
| Barthel score<100 (ref.: Barthel score=100) | | 914 | 14/1 | 160 (22%) | 59 (32%) | *1·69 (1·18; 2·42)* |
| IADL<8 (ref.: IADL=8) | | 834 | 85/10 | 69 (10%) | 35 (20%) | *2·14 (1·37; 3·35)* |
| Any past falling incident (ref.: no history of falls) | | 668 | 223/38 | 118 (23%) | 32 (22%) | 0·96 (0·61; 1·48) |
| TUG>10s (ref. TUG ≤10s) | | 788 | 102/39 | 156 (24%) | 53 (37%) | *1·80 (1·22; 2·63)* |
| Preoperative EQ5D | | 746 | 151/32 | 0.84±0.18 | 0.81±0.21 | 0·418 (0·166; 1·048) |
| Frailty (Fried, reference: robust) | Prefrail | 920 | 7/2 | 366 (50%) | 100 (55%) | *1·90 (1·49; 2·44)* |
| | Frail | | | 84 (11%) | 42 (23%) | |
| Frailty (SOF, reference: robust) | Prefrail | 631 | 256/42 | 128 (26%) | 47 (33%) | *1·83 (1·44; 2·33)* |
| | Frail | | | 63 (12%) | 39 (27%) | |
| MMSE <27p | | 929 | 0/0 | 56 (8%) | 37 (20%) | *3·10 (1·96; 4·85)* |
| Preoperative cognitive impairment (ref.: no PreCI) | | 923 | 4/1 | 79 (11%) | 43 (23%) | *2·57 (1·69; 3·88)* |
| GPT (ref·: z-score ≥-1·96) | | 868 | 38/23 | 54 (8%) | 22 (14%) | *1·91 (1·13; 3·25)* |
| VRM free recall (ref·: z-score ≥-1·96) | | 899 | 23/7 | 15 (2%) | 11 (6%) | *3·12 (1·41; 6·92)* |
| VRM recognition (ref·: z-score ≥-1·96) | | 824 | 87/18 | 22 (3%) | 11 (7%) | 2·05 (0·97; 4·32) |
| SRT (ref·: z-score ≥-1·96) | | 906 | 18/5 | 22 (3%) | 10 (6%) | 1·90 (0·88; 4·08) |
| SSP (ref·: z-score ≥-1·96) | | 911 | 13/5 | 102 (14%) | 26 (15%) | 1·05 (0·66; 1·67) |
| TMT-B (ref·: z-score ≥-1·96) | | 841 | 55/33 | 27 (4%) | 8 (5%) | 1·37 (0·61; 3·09) |
| PAL (ref·: z-score ≥-1·96) | | 905 | 16/8 | 74 (10%) | 25 (14%) | 1·47 (0·90; 2·38) |

**eTable 8: Exploratory analysis of continuously scaled neurocognitive test data of 745 patients without and 184 patients with POD.**

Abbreviations: GPT: Grooved Pegboard Test, VRM: Verbal Recognition Memory, SRT: Simple Reaction Time (mean latency), SSP: simple span length, TMT-B: Trail-Making-Test part B, PAL: Paired Associate Learning (memory score). Data are given number of included patients (N), missing data for patients without/with POD (nNPOD/nPOD), mean ± standard deviation or the absolute (relative) number of patients with a predictive characteristic among patients without/with POD. Odds ratios (OR) with 95% confidence intervals (CI).

| | N | Missing $n_{NPOD}/n_{POD}$ | No POD | POD | OR (95% CI) |
|---|---|---|---|---|---|
| **GPT (s)** | 868 | 38/23 | 96·7±28·5 | 109·4±39·8 | *1·011 (1·006; 1·016)* |
| **VRM free recall (no.)** | 899 | 23/7 | 6·0±1·9 | 5·3±2·0 | *0·82 (0·75; 0·90)* |
| **VRM recognition (no.)** | 824 | 87/18 | 21·7±2·0 | 21·3±2·2 | 0·93 (0·86; 1·01) |
| **SRT (ms)** | 906 | 18/5 | 323±101 | 352±137 | *1·002 (1·001; 1·003)* |
| **SSP (no.)** | 911 | 13/5 | 4·8±1·1 | 4·6±1·0 | *0·82 (0·70; 0·96)* |
| **TMT-B (s)** | 841 | 55/33 | 113±47 | 127±51 | *1·006 (1·002; 1·009)* |
| **PAL (no.)** | 905 | 16/8 | 13·5±4·6 | 12·4±4·8 | *0·95 (0·92; 0·98)* |

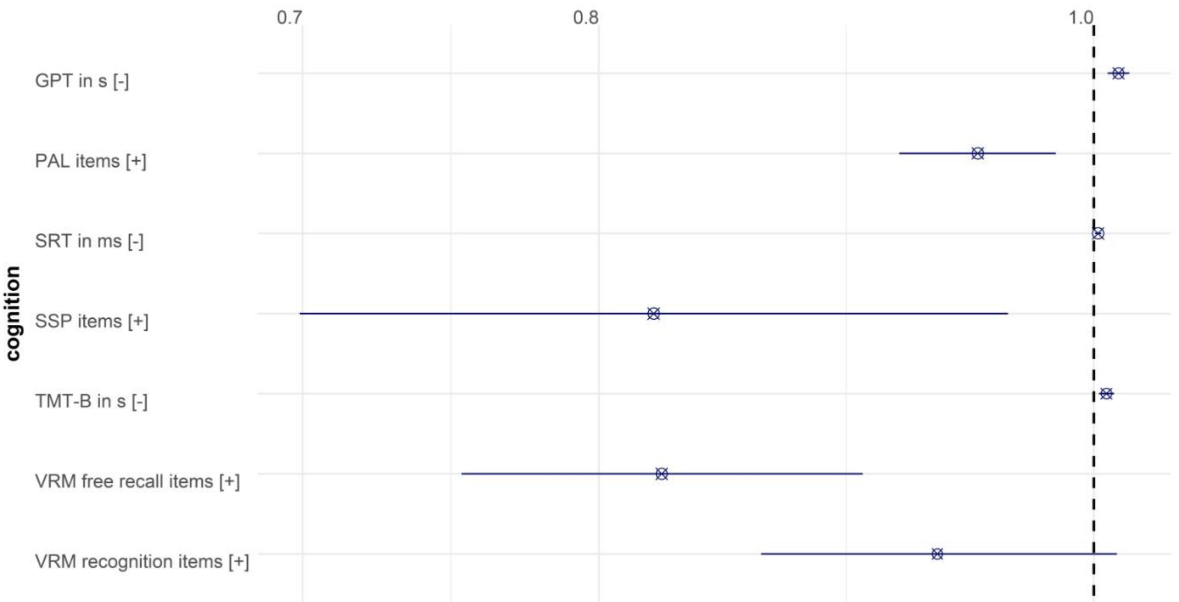


**eFigure 5: Odds ratios with 95% confidence intervals for association of neuropsychological tests with POD.**

For interpretation of the effect directions, [-] indicates that a higher test score is associated with poor cognitive performance, whereas [+] indicates that a high score indicates better performance. Abbreviations: GPT: Grooved Pegboard Test, ms: milliseconds VRM: Verbal Recognition Memory, s: seconds, SRT: Simple Reaction Time (mean latency), SSP: simple span length, TMT-B: Trail-Making-Test part B, PAL: Paired Associate Learning (memory score).

eTable 9: Nutrition and drug consumption in 745 patients without and 184 patients with POD.

Abbreviations: Alcohol Use Disorder Identification Test, BMI: Body Mass Index, MNA-SF: Mini-Nutritional Assessment Short Form. Data are given number of included patients (N), missing data for patients without/with POD (nNPOD/nPOD), mean ± standard deviation or the absolute (relative) number of patients with a predictive characteristic among patients without/with POD. Odds ratios (OR) with 95% confidence intervals (CI).

| | | N | Missing $n_{NPOD}/n_{POD}$ | No POD | POD | OR (95% CI) |
|---|---|---|---|---|---|---|
| **AUDIT (ref.: <5p for women, <8p for men)** | | 862 | 51/16 | 44 (7%) | 14 (8%) | 1·28 (0·66; 2·33) |
| **Current smoking (ref: nonsmoker)** | | 903 | 17/9 | 72 (10%) | 18 (10%) | 1·04 (0·59; 1·76) |
| **Eversmoker (ref.: neversmoker)** | | 798 | 100/31 | 389 (60%) | 91 (59%) | 0·97 (0·67; 1·38) |
| **Pack years** | | 734 | 154/51 | 10.30±16.6 | 11.5±18.70 | 1·005 (0·994; 1·016) |
| **BMI** | **<18·5kg/m²** | 933 | 0/0 | 10 (1%) | 3 (2%) | 0·97 (0·79; 1·20) |
| | **18·5-24·99kg/m²** | | | 233 (31%) | 63 (34%) | |
| | **25-29·99kg/m²** | | | 338 (45%) | 74 (40%) | |
| | **≥30kg/m²** | | | 164 (22%) | 44 (24%) | |
| **Obesity (BMI≥30kg/m², ref: BMI 18·5-29·99kg/m²)** | | 916 | 0/0 | 164 (22%) | 44 (24%) | 1·12 (0·76; 1·64) |
| **Underweight (BMI <18·5kg/m², ref: BMI 18·5-29·99kg/m²)** | | 721 | 0/0 | 10 (2%) | 3 (2%) | 1·25 (0·34; 4·61) |
| **MNA-SF (ref.: 12-14p)** | **0-7p** | 911 | 16/2 | 33 (5%) | 16 (9%) | *1·67 (1·29; 2·16)* |
| | **8-11p** | | | 145 (20%) | 55 (33%) | |

##### eTable 10: Precipitating factors in 745 patients without and 184 patients with POD

In total, type of surgery was analyzed in 911 subjects, and reduced sample sizes are a consequence of comparing two types of surgery whilst excluding the third one, respectively. Data are given number of included patients (N), missing data for patients without/with POD (nNPOD/nPOD), mean ± standard deviation or the absolute (relative) number of patients with a predictive characteristic among patients without/with POD. Odds ratios (OR) with 95% confidence intervals (CI). Abbreviations: ICU: intensive care unit.

| | | N | Missing nNPOD/nPOD | No POD | POD | OR (95% CI) |
|---|---|---|---|---|---|---|
| **Duration of anesthesia >4h (ref.: ≤4h)** | | 912 | 17/0 | 228 (31%) | 123 (67%) | *4·42 (3·15; 6·27)* |
| **Duration of surgery >4h (ref.: ≤4h)** | | 684 | 202/43 | 56 (10%) | 65 (46%) | *7·44 (4·84; 11·50)* |
| **Regional anesthesia (ref.: general or combined general/regional anesthesia)** | | 912 | 13/4 | 176 (7%) | 4 (2%) | *0·29 (0·09; 0·72)* |
| **Surgery** | **Intracranial (ref.: peripheral)** | 514 | 294/121 | 8 (2%) | 2 (3%) | 1·82 (0·37; 8·75) |
| | **Intrathoracic, -abdominal, -pelvic (ref.: peripheral)** | 901 | 21/7 | 281 (39%) | 116 (66%) | *3·00 (2·13; 4·25)* |
| **Any anticholinergic medication on postoperative day 0-7 (ref.: no medication)** | | 903 | 17/9 | 537 (74%) | 152 (87%) | *2·35 (1·50; 3·84)* |
| **Postoperative pain (ref.: no pain)** | | 904 | 20/5 | 242 (33%) | 93 (52%) | *2·16 (1·55; 3·01)* |

**eTABLE 11: TIME-ADJUSTED ANALYSIS OF THE ASSOCIATION BETWEEN POSTOPERATIVE ANTICHOLINERGIC MEDICATION AND DELIRIUM.**

This analysis accounts for the temporal relationship between the application of anticholinergic medication and the occurrence of delirium. Thus, univariate analyses shown here describe the association of anticholinergic medication given on the day of surgery until postoperative day X with delirium occurrence on postoperative day X+1 until postoperative day 7/discharge. Data are given number of included patients (N), missing data for patients without/with POD (nNPOD/nPOD), mean ± standard deviation or the absolute (relative) number of patients with a predictive characteristic among patients without/with POD. Odds ratios (OR) with 95% confidence intervals (CI). Patients without anticholinergic medication were defined as the reference group.

| | N | Missing $n_{NPOD}/n_{POD}$ | No POD after anticholinergic medication | POD after anticholinergic medication | OR (95% CI) |
|---|---|---|---|---|---|
| **Anticholinergic medication on day of surgery** | 699 | 165/56 | 408 (65%) | 54 (78%) | *1·96 (1·11; 3·67)* |
| **Anticholinergic medication until postoperative day 1** | 820 | 14/9 | 498 (68%) | 80 (87%) | *3·08 (1·71; 6·05)* |
| **Anticholinergic medication until postoperative day 2** | 675 | 15/8 | 428 (73%) | 81 (94%) | *6·09 (2·68; 17·56)* |
| **Anticholinergic medication until postoperative day 3** | 561 | 14/5 | 364 (76%) | 76 (95%) | *6·11 (2·47; 20·34)* |
| **Anticholinergic medication until postoperative day 4** | 497 | 14/5 | 337 (80%) | 72 (96%) | *6·05 (2·19; 25·13)* |
| **Anticholinergic medication until postoperative day 5** | 422 | 13/5 | 295 (83%) | 67 (99%) | *13·4 (2·87; 238·90)* |
| **Anticholinergic medication until postoperative day 6** | 348 | 10/3 | 250 (87%) | 62 (100%) | n.a. |

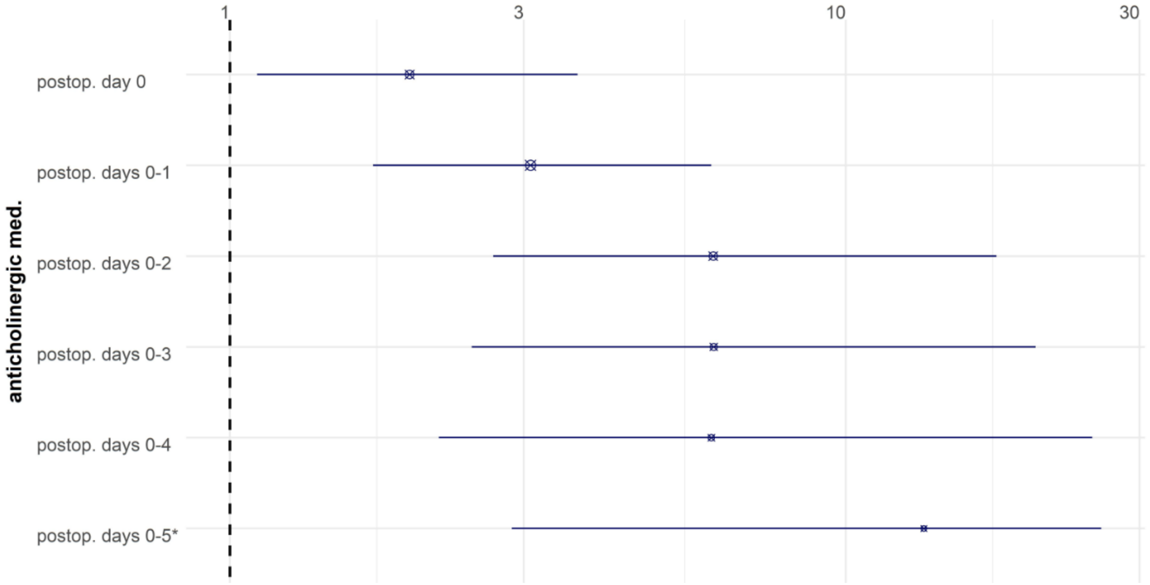


**eFigure 6 Time-adjusted analyses of postoperative anticholinergic prescriptions and delirium.**

The associations of anticholinergic prescriptions until postoperative day 1 and delirium on day 2 or later as well as prescriptions until day 2 and delirium on day 3 or later, and so forth, have been analyzed. The figure displays odds ratios with 95% confidence intervals. The day of surgery is counted as postoperative day 0.

* the upper limit of the confidence interval has been truncated at 26

Abbreviations: med.: medication; postop.: postoperative

###### eTable 12: Time-adjusted analysis of the association between postoperative pain and delirium

The analyses accounts for the temporal relationship between postoperative pain and the occurrence of delirium. Thus, univariate analyses shown describe the association of postoperative pain on the day of surgery until postoperative day X with delirium occurrence on postoperative day X+1 until postoperative day 7/discharge. Data are given number of included patients (N), missing data for patients without/with POD (nNPOD/nPOD), mean ± standard deviation or the absolute (relative) number of patients with a predictive characteristic among patients without/with POD. Odds ratios (OR) with 95% confidence intervals (CI). For calculation of ORs, patients without pain are treated as the reference group.

| | N | Missing nNPOD/nPOD | No POD after pain | POD after pain | OR (95% CI) |
|---|---|---|---|---|---|
| **Postoperative pain on the day of surgery** | 705 | 160/55 | 223 (35%) | 24 (34%) | 0·96 (0·57; 1·61) |
| **Postoperative pain until postoperative day 1** | 805 | 30/8 | 283 (40%) | 50 (54%) | *1·76 (1·14; 2·73)* |
| **Postoperative pain until postoperative day 2** | 638 | 22/8 | 268 (46%) | 48 (56%) | 1·48 (0·94; 2·34) |
| **Postoperative pain until postoperative day 3** | 556 | 19/5 | 235 (49%) | 49 (61%) | *1·62 (1·00; 2·65)* |
| **Postoperative pain until postoperative day 4** | 493 | 18/5 | 227 (54%) | 51 (68%) | *1·79 (1·07; 3·06)* |
| **Postoperative pain until postoperative day 5** | 420 | 15/5 | 337 (96%) | 66 (97%) | 1·47 (0·40; 9·46) |
| **Postoperative pain until postoperative day 6** | 347 | 11/3 | 274 (96%) | 60 (67%) | 1·20 (0·31; 7·91) |

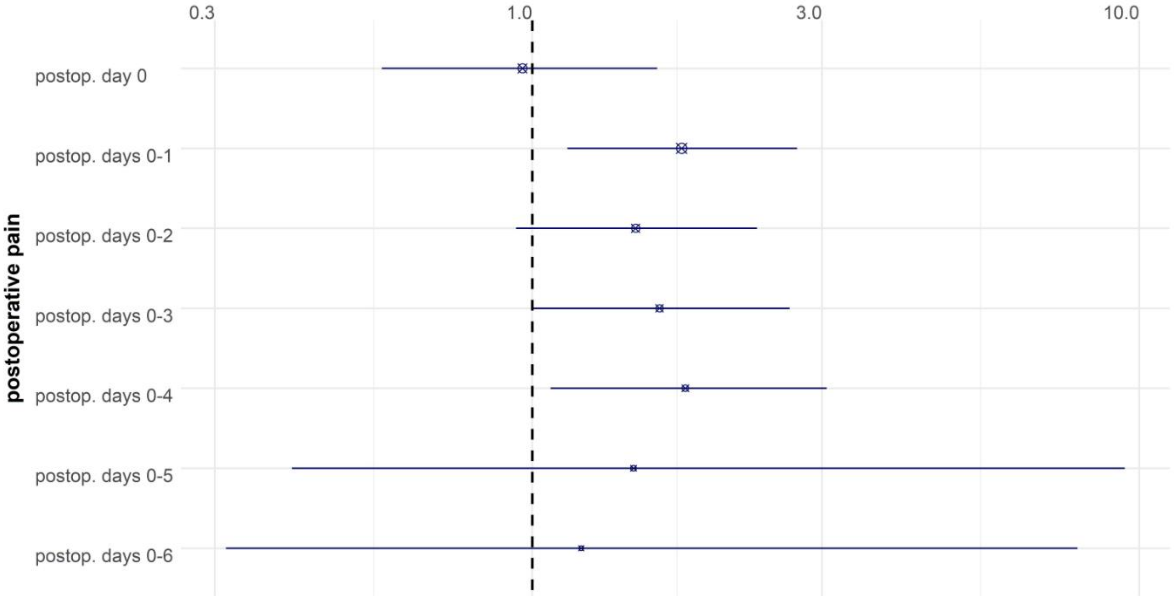


**eFIGURE 7: TIME-ADJUSTED ANALYSES OF POSTOPERATIVE THERAPY-DEMANDING PAIN AND DELIRIUM.**

The associations of pain until postoperative day 1 and delirium on day 2 or later as well as between pain until day 2 and delirium on day 3 or later, and so forth, have been analyzed. The figure displays odds ratios with 95% confidence intervals. The day of surgery is counted as postoperative day 0.

Abbreviations: postop.: postoperative

**eTable 13: Baseline values for blood-based markers in 745 patients without and 184 patients with delirium.**

* Odds ratios for standardized variables. ** Odds ratios for laboratory adjusted standardized variables. Abbreviations: CRP: C-reactive protein, IL: interleukin, NT-proBNP: N-terminal pro-brain natriuretic peptide, ADMA/SDMA: asymmetric/symmetric dimethylarginine, MCV: mean corpuscular volume, RDW: red cell distribution width, (N)HMW: (non) high molecular weight, LDL/HDL: low/high-density lipoprotein, SORL1: sortilin-related receptor-1, ALT: alanine aminotransferase, AST: aspartate aminotransferase, γ-GT: γ-glutamyltransferase, LDH: lactate dehydrogenase. Data are given number of included patients (N), missing data for patients without/with POD (nNPOD/nPOD), mean ± standard deviation or the absolute (relative) number of patients with a predictive characteristic among patients without/with POD. Odds ratios (OR) with 95% confidence intervals (CI).

| | N | Missing $n_{NPOD}/n_{POD}$ | No POD | POD | OR (95% CI) |
|---|---|---|---|---|---|
| **CRP (mg/L)** | 775 | 125/29 | 7·9±16·9 | 12·1±18·4 | _1·20 (1·03; 1·41)_** |
| **IL2 (pg/mL)** | 200 | 576/153 | 1089±1432 | 1139±1471 | 1·03 (0·72; 1·48)* |
| **IL6 (pg/mL)** | 801 | 102/26 | 5·9±24·7 | 12·7±42·6 | _1·19 (1·03; 1·38)_* |
| **IL8 (pg/mL)** | 178 | 610/141 | 574±647 | 703±768 | 1·19 (0·87; 1·63)* |
| **IL8, whole blood (pg/mL)** | 178 | 628/123 | 450±519 | 673±777 | _1·42 (1·02; 1·98)_* |
| **IL10 (pg/mL)** | 55 | 702/172 | 272±195 | 309±167 | 1·21 (0·66; 2·19)* |
| **IL18 (pg/mL)** | 730 | 160/39 | 42·6±34·5 | 42·8±26·1 | 1·01 (0·84; 1·20)* |
| **Calprotectin (ng/mL)** | 275 | 511/143 | 1435±1400 | 1968±1928 | 1·29 (0·99; 1·71)* |
| **Troponin T (pg/mL)** | 767 | 133/29 | 16·3±8·9 | 18·1±10·4 | 1·15 (0·98; 1·35)** |
| **NT-proBNP (pmol/L)** | 271 | 515/143 | 21·9±57·0 | 19·7±33·2 | 0·96 (0·66; 1·39)* |
| **S100A12 (ng/mL)** | 772 | 128/29 | 156±199 | 193±153 | 1·15 (0·98; 1·35)** |
| **Homoarginin (µmol/L)** | 271 | 514/144 | 1·5±0·6 | 1·6±0·6 | 1·03 (0·74; 1·43)* |
| **ADMA (µmol/L)** | 271 | 514/144 | 0·70±0·10 | 0·70±0·10 | 1·21 (0·87; 1·68)* |
| **SDMA (µmol/L)** | 271 | 514/144 | 0·8±0·2 | 0·9±0·3 | 1·29 (0·97; 1·71)* |
| **Hemoglobin (g/dL)** | 764 | 142/23 | 13·1±1·8 | 12·4±2·0 | _0·67 (0·56; 0·79)_** |
| **MCV (fL)** | 714 | 174/41 | 90·4±4·6 | 90·8±5·1 | 1·08 (0·90; 1·29)** |
| **Thrombocytes (1/nL)** | 735 | 161/33 | 227±70 | 236±80 | 1·13 (0·94; 1·33)** |
| **Reticulated platelets (%)** | 136 | 628/165 | 3·2±1·6 | 2·8±1·2 | 0·75 (0·44; 1·30)* |
| **Leukocytes (1/nL)** | 718 | 172/39 | 6·5±2·2 | 6·9±3·0 | 1·19 (1·00; 1·40)** |
| **Lymphocytes (1/nL)** | 696 | 189/44 | 1·7±0·6 | 1·5±0·7 | 0·84 (0·69; 1·02)** |
| **Neutrophils (1/nL)** | 695 | 189/45 | 3·9±1·7 | 4·3±2·4 | _1·22 (1·03; 1·46)_** |
| **Eosinophils (1/nL)** | 696 | 189/44 | 0·2±0·1 | 0·2±0·1 | 1·02 (0·85; 1·22)** |
| **Basophils (1/nL)** | 696 | 189/44 | 0±0 | 0±0 | 0·98 (0·82; 1·19)** |
| **Immature granulocytes (%)** | 561 | 305/63 | 0·4±0·3 | 0·5±0·6 | _1·34 (1·10; 1·63)_* |
| **HMW adiponectin (ng/mL)** | 802 | 101/26 | 1·9±1·3 | 2·0±1·5 | 1·06 (0·89; 1·25)* |
| **NHMW adiponectin (ng/mL)** | 800 | 103/26 | 12·6±7·2 | 13·0±8·0 | 1·06 (0·89; 1·25)* |
| **Total adiponectin (ng/mL)** | 800 | 103/26 | 14·6±8·1 | 15·1±9·0 | 1·06 (0·89; 1·26)* |
| **C peptide (pmol/mL)** | 764 | 135/30 | 1·1±0·7 | 1·2±0·7 | 0·96 (0·79; 1·14)** |
| **Intact proinsulin (pmol/L)** | 274 | 512/143 | 7·9±8·9 | 9·5±9·2 | 1·16 (0·88; 1·53)* |
| **Glucose (mmol/L)** | 766 | 90/16 | 5·9±1·7 | 6·2±1·9 | 1·16 (0·98; 1·35)** |
| **HbA1c (mmol/mol)** | 713 | 175/41 | 36·7±8·8 | 36·5±9·6 | 1·01 (0·84; 1·21)** |
| **Leptin (ng/mL)** | 753 | 140/36 | 24·0±27·1 | 22·4±28·9 | 1·05 (0·88; 1·25)** |

| | N | Missing $n_{NPOD}/n_{POD}$ | No POD | POD | OR (95% CI) |
|---|---|---|---|---|---|
| **Soluble leptin rec. (ng/mL)** | 743 | 156/32 | 20·8±9·4 | 21·9±12·8 | 1·08 (0·91; 1·28)** |
| **Leptin/adiponectin ratio** | 750 | 143/36 | 2·1±2·7 | 2·1±2·9 | 1·07 (0·89; 1·26)** |
| **Leptin/soluble leptin receptor ratio** | 741 | 166/38 | 1·7±3·0 | 1·8±3·8 | 1·09 (0·92; 1·28)** |
| **LDL (mmol/L)** | 715 | 174/40 | 3·1±1·0 | 2·9±1·0 | *0·82 (0·68; 0·99)*** |
| **Oxidized LDL (ng/mL)** | 272 | 514/143 | 210±394 | 178±311 | 0·91 (0·62; 1·33)* |
| **HDL (mmol/L)** | 783 | 115/31 | 1·4±4·0 | 1·2±0·4 | *0·72 (0·59; 0·87)*** |
| **Triglycerides (mmol/L)** | 776 | 120/33 | 1·7±1·5 | 1·8±0·9 | 1·07 (0·90; 1·25)** |
| **Total cholesterol (mmol/L)** | 716 | 173/40 | 4·9±1·2 | 4·6±1·2 | *0·79 (0·65; 0·95)*** |
| **Malondialdehyde (µmol/L)** | 775 | 125/29 | 1·3±0·6 | 1·4±0·9 | 1·17 (0·99; 1·37)** |
| **Nitrotyrosine (nM)** | 273 | 513/143 | 461±458 | 455±328 | 0·99 (0·70; 1·39)* |
| **Tryptophan (µmol/L)** | 773 | 126/30 | 48·3±12·7 | 44·4±13·5 | *0·74 (0·62; 0·89)*** |
| **Kynurenin (µmol/L)** | 772 | 127/30 | 2·8±0·9 | 2·9±1·0 | 1·14 (0·96; 1·34)** |
| **Kynurenin-acid (nM)** | 273 | 512/144 | 40·8±39·5 | 57·7±95·5 | 1·23 (0·96; 1·58)* |
| **ALT (U/L)** | 717 | 172/40 | 27·2±32·7 | 32·9±59·5 | 1·12 (0·96; 1·33)** |
| **AST (U/L)** | 748 | 172/39 | 29·0±32·6 | 24·4±63·2 | 1·12 (0·95; 1·34)** |
| **γ-GT (U/L)** | 716 | 173/40 | 50·1±103·3 | 86·7±182·5 | *1·24 (1·06; 1·47)*** |
| **Albumin (g/L)** | 644 | 238/47 | 40·5±4·2 | 38·6±5·4 | *0·68 (0·56; 0·81)*** |
| **Creatinine (µm/L)** | 723 | 166/40 | 80·3±28·0 | 86·0±56·1 | 1·15 (0·98; 1·35)** |
| **Potassium (mmol/L)** | 760 | 146/23 | 4·2±0·4 | 4·2±0·5 | 1·02 (0·86; 1·22)** |
| **LDH (U/L)** | 689 | 199/41 | 231·6±97·6 | 238·2±98·2 | 1·05 (0·87; 1·24)** |
| **Uric acid (mmol/L)** | 717 | 172/40 | 0·345±0·089 | 0·344±0·105 | 1·02 (0·84; 1·22)** |
| **β-Amyloid 40 (pg/mL)** | 796 | 107/26 | 285·6±84·6 | 300·5±92·7 | *1·20 (1·02; 1·41)*** |
| **β-Amyloid 42 (pg/mL)** | 789 | 113/27 | 43·7±45·5 | 41·6±24·0 | 0·86 (0·68; 1·09)** |
| **β-Amyloid Aβ42/Aβ40-ratio** | 789 | 113/27 | 0·148±0·101 | 0·138±0·063 | *0·74 (0·56; 0·93)*** |

eTABLE 14: DIFFERENCES BETWEEN BASELINE AND POSTOPERATIVE LEVELS OF BLOOD-BASED MARKERS IN 745 PATIENTS WITHOUT AND 184 PATIENTS WITH DELIRIUM

* Odds ratios for standardized variables.

** Odds ratios for laboratory adjusted standardized variables. Abbreviations: CRP: C-reactive protein, IL: interleukin, NT-proBNP: N-terminal pro-brain natriuretic peptide, ADMA/SDMA: asymmetric/symmetric dimethylarginine, MCV: mean corpuscular volume, RDW: red cell distribution width, (N)HMW: (non-) high molecular weight, LDL/HDL: low/high-density lipoprotein, SORL1: sortilin-related receptor-1, ALT: alanine aminotransferase, AST: aspartate aminotransferase, γ-GT: γ-glutamyltransferase, LDH: lactate dehydrogenase. Data are given number of included patients (N), missing data for patients without/with POD (nNPOD/nPOD), mean ± standard deviation or the absolute (relative) number of patients with a predictive characteristic among patients without/with POD. Odds ratios (OR) with 95% confidence intervals (CI).

| | N | Missing $n_{NPOD}/n_{POD}$ | No POD | POD | OR (95% CI) |
|---|---|---|---|---|---|
| **CRP (mg/L)** | 242 | 538/149 | 31·73 (46·62) | 55·75 (45·60) | 1·59 (1·14; 2·21)** |
| **IL2 (pg/mL)** | 77 | 682/170 | -373·59 (2048·23) | -157·29 (987·38) | 1·17 (0·54; 2·51)* |
| **IL6 (pg/mL)** | 706 | 184/39 | 66·10 (94·53) | 139·11 (134·00) | 1·76 (1·48; 2·09)* |
| **IL8 (pg/mL)** | 127 | 653/149 | 630·29 (1470·21) | 2116·17 (3601·40) | 1·96 (1·18; 3·24)* |
| **IL10 (pg/mL)** | 24 | 729/176 | -48·75 (302·31) | 458·38 (941·41) | 5·56 (0·47; 66·26)* |
| **IL18 (pg/mL)** | 611 | 261/57 | 3·43 (20·56) | 13·53 (31·84) | 0·77 (0·49; 1·22)* |
| **Calprotectin (ng/mL)** | 248 | 534/147 | 1023·80 (1859·21) | 1532·62 (2693·39) | 1·25 (0·92; 1·7)* |
| **Troponin T (pg/mL)** | 243 | 537/149 | 6·84 (14·87) | -0·64 (14·39) | 0·58 (0·39; 0·86)* |
| **S100A12 (ng/mL)** | 186 | 587/156 | 0·05 (3·06) | 1·16 (4·07) | 1·5 (0·95; 2·39)* |
| **Homoarginin (µmol/L)** | 240 | 540/149 | 0·08 (0·45) | -0·22 (0·54) | 0·48 (0·31; 0·73)* |
| **ADMA (µmol/L)** | 240 | 540/149 | -0·04 (0·13) | -0·08 (0·15) | 0·69 (0·48; 1·00)* |
| **SDMA (µmol/L)** | 240 | 540/149 | 0·12 (0·21) | 0·11 (0·35) | 0·95 (0·66; 1·37)* |
| **Hemoglobin (g/dL)** | 675 | 212/42 | 0·06 (0·94) | -0·22 (1·16) | 0·76 (0·63; 0·91)** |
| **MCV (fL)** | 615 | 256/58 | 0·02 (0·92) | -0·08 (1·26) | 0·90 (0·74; 1·10)** |
| **Thrombocytes (1/nL)** | 625 | 251/53 | 0·13 (0·87) | -0·48 (1·28) | 0·57 (0·46; 0·69)** |
| **Reticulated platelets (%)** | 109 | 653/167 | 0·46 (2·00) | 1·13 (1·61) | 1·41 (0·84; 2·37)* |
| **Leukocytes (1/nL)** | 620 | 252/57 | -0·07 (0·90) | 0·25 (1·29) | 1·36 (1·12; 1·64)** |
| **Lymphocytes (1/nL)** | 567 | 290/72 | 0·08 (0·99) | -0·34 (0·95) | 0·66 (0·54; 0·81)** |
| **Neutrophils (1/nL)** | 567 | 289/73 | -0·08 (0·95) | 0·32 (1·12) | 1·47 (1·2; 1·81)** |
| **Eosinophils (1/nL)** | 567 | 290/72 | 0·04 (0·95) | -0·15 (1·16) | 0·83 (0·68; 1·02)** |
| **Basophils (1/nL)** | 567 | 290/72 | 0·01 (1·00) | -0·02 (1·01) | 0·98 (0·79; 1·2)** |
| **HMW adiponectin (ng/mL)** | 686 | 201/42 | 0·06 (0·95) | -0·24 (1·13) | 0·7 (0·59; 0·85)** |
| **NHMW adiponectin (ng/mL)** | 679 | 207/43 | 0·04 (0·95) | -0·15 (1·18) | 0·79 (0·66; 0·93)** |
| **Total adiponectin (ng/mL)** | 680 | 206/43 | 0·47 (0·94) | -0·18 (1·19) | 0·76 (0·64; 0·90)** |
| **C peptide (pmol/mL)** | 656 | 227/46 | 0·05 (1·00) | -0·17 (0·97) | 0·83 (0·67; 1·03)** |

| | | | | | |
|---|---|---|---|---|---|
| **Intact proinsulin (pmol/L)** | 240 | 540/149 | 8·88 (14·10) | 11·92 (13·50) | 1·21 (0·88; 1·68)* |
| **Glucose (mmol/L)** | 667 | 225/37 | -0·04 (0·99) | 0·13 (1·03) | 1·18 (0·98; 1·41)** |
| **HbA1c (mmol/mol)** | 615 | 256/58 | -0·01 (1·03) | 0·02 (0·86) | 1·03 (0·84; 1·26)** |
| **Leptin (ng/mL)** | 650 | 230/49 | 0·02 (1·05) | -0·06 (0·78) | 0·9 (0·73; 1·11)** |
| **Soluble leptin rec. (ng/mL)** | 673 | 213/43 | -0·03 (0·91) | 0·09 (1·28) | 0·87 (0·71; 1·08)** |
| **Leptin/adiponectin ratio** | 646 | 233/50 | 0·003 (1·003) | -0·01 (0·98) | 0·96 (0·79; 1·17)** |
| **Leptin/soluble leptin receptor ratio** | 619 | 258/52 | 0·04 (1·04) | -0·14 (0·82) | 0·80 (0·64; 1·01)* |
| **LDL (mmol/L)** | 621 | 252/56 | 0·12 (0·91) | -0·45 (1·17) | 0·58 (0·47; 0·71)** |
| **Oxidized LDL (ng/mL)** | 242 | 539/148 | -37·37 (225·84) | -64·38 (201·51) | 0·50 (0·66; 1·23)* |
| **HDL (mmol/L)** | 623 | 250/56 | 0·04 (0·94) | -0·15 (1·21) | 0·82 (0·68; 1·00)** |
| **Triglycerides (mmol/L)** | 552 | 312/65 | 0·09 (1·01) | -0·33 (0·91) | 0·66 (0·53; 0·83)** |
| **Total cholesterol (mmol/L)** | 623 | 250/56 | 0·12 (0·93) | -0·48 (1·11) | 0·54 (0·44; 0·67)** |
| **Malondialdehyde (µmol/L)** | 245 | 536/148 | -0·12 (0·64) | '-0·38 (1·30) | 0·78 (0·59; 1·04)* |
| **Nitrotyrosine (nM)** | 243 | 538/148 | -27·66 (121·01) | -83·08 (221·99) | 0·72 (0·53; 0·98)* |
| **Kynurenin (µmol/L)** | 243 | 537/149 | 0·09 (0·80) | 0·12 (0·78) | 1·04 (0·73; 1·47)* |
| **Kynurenin-acid (nM)** | 243 | 537/149 | -0·97 (16·19) | 4·66 (29·37) | 1·29 (0·94; 1·77)* |
| **ALT (U/L)** | 620 | 252/57 | -0·07 (0·37) | 0·26 (2·07) | 1·97 (1·13; 3·41)** |
| **AST (U/L)** | 625 | 249/55 | -0·07 (0·76) | 0·26 (1·60) | 1·45 (1·02; 2·06)** |
| **γ-GT (U/L)** | 622 | 251/56 | 0·05 (0·64) | -0·20 (1·80) | 0·81 (0·68; 0·98)** |
| **Albumin (g/L)** | 554 | 312/63 | 0·09 (0·93) | -0·32 (-3·81) | |
| **Creatinine (µm/L)** | 509 | 345/75 | 0·07 (0·20) | 0·10 (0·34) | 1·14 (0·96; 1·36)* |
| **Potassium (mmol/L)** | 665 | 224/40 | -0·07 (1·00) | 0·25 (0·95) | 1·4 (1·12; 1·64)** |
| **LDH (U/L)** | 598 | 272/59 | -0·05 (0·60) | 0·20 (1·83) | 1·33 (0·99; 1·77)** |
| **Uric acid (mmol/L)** | 623 | 250/56 | 0·07 (0·92) | -0·27 (1·23) | 0·71 (0·58; 0·86)** |

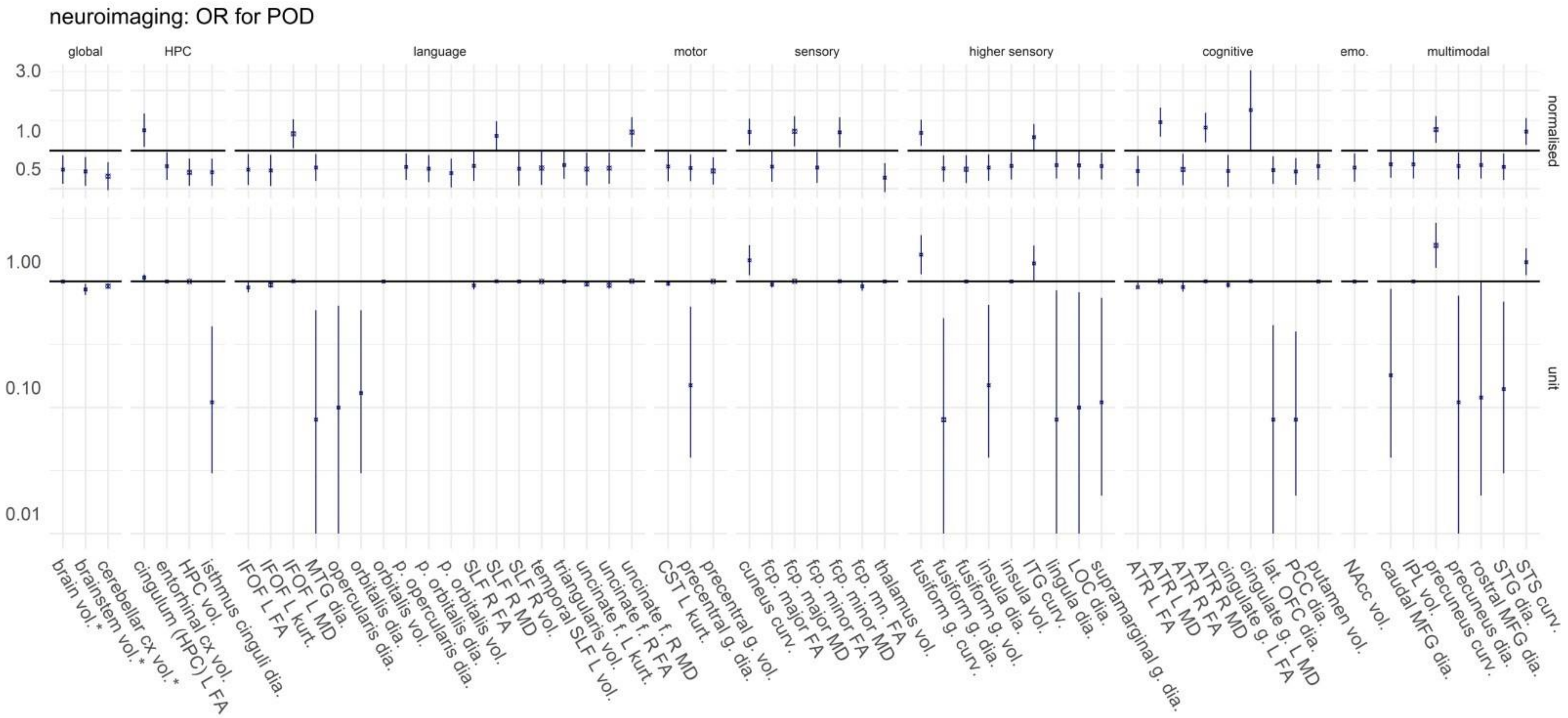


**EFIGURE 8: SUMMARY OF NEUROIMAGING VARIABLES FOR DIFFERENT BRAIN REGIONS.**

Odds ratios (OR) with corresponding 95% confidence interval are given for normalized variables (top) and for one unit change (bottom). In the upper chart, the odds ratio corresponds to change of one sample-specific standard deviation. In the lower chart, the odds ratios are generally given for a change of 1mm³ for volume, 1cm for diameter, 1/dm for mean curvature, 1% for fractional anisotropy, 1µm²/s for mean diffusivity and a 0·01 increase in kurtosis (unit-free). * For these volumes, odds ratios are given per 1cm³ change. Abbreviations: ATR: anterior thalamic radiation, CST: corticospinal tract, curv: mean curvature, cx: cortex, dia.: diameter (cortical thickness), f.: fascicle, FA: fractiona anisotropy, fcp: forceps, g.: gyrus, HPC: hippocampus, FG: inferior frontal gyrus, IFL: inferior parietal lobule, IFOF: inferior fronto-occipital fascicle, inf.: inferior, L: left hemisphere, lat.: lateral, LOC: lateral occipital cortex, long.: longitudinal, MD: mean diffusivity, MFG: middle frontal gyrus, MTG: middle temporal gyrus, ncl.: nucleus, NAcc: nucleus accumbens, OFC: orbitofrontal cortex, PCC: posterior cingulate cortex, ped.: peduncle, R: right hemisphere, SLF: superior longitudinal fascicle, STG: superior temporal gyrus, STS: superior temporal sulcus, sup.: superior, tr.: tract, vol.: volume

#### eTable 15: Neuroimaging in 745 patients without and 184 patients with POD.

No additional adjustments for global brain volume, center or scanner were made.

* Exclusion MRI data due to artifacts was left to the discretion of the individual researcher, resulting in different sample sizes for each neuroimaging modality. Ant.: anterior, BFCS: basal forebrain cholinergic system, FA: fractional anisotropy, fasc.: fasciculus, inf.: inferior, L: left, MD: mean diffusivity, NBM: Nucleus basalis magnocellularis (of Meynert), R: right., sup.: superior. Data are given number of included patients (N), missing data for patients without/with POD (nNPOD/nPOD), mean ± standard deviation or the absolute (relative) number of patients with a predictive characteristic among patients without/with POD. Odds ratios (OR) with 95% confidence intervals (CI). Standardization refers to the process of transforming a variable by centering around the mean and setting the standard deviation to 1. Thus, the odds ratio corresponds to a parameter change of one standard deviation based on the investigated sample. Standardized OR are only given for variables with 95% CI not crossing unity. Unless stated otherwise, values are reported in % for fractional anisotropy, µm²/s for mean diffusivity, $10^{-2}$ for kurtosis, cm for thickness, $dm^{-1}$ for mean curvature and mm³ for volume.

| | N | Missing $n_{NPOD}/n_{POD}$ | No POD | POD | OR (95% CI) | Standardized OR |
|---|---|---|---|---|---|---|
| **Brain volume (cm³)** | 478 | 342/109 | 1014±112 | 977±108 | *0·997 (0·995; 0·999)* | *0·71 (0·55; 0·92)* |
| **BFCS volume** | 478 | 342/109 | 2227±226 | 2173±219 | 0·999 (0·998; 1·000) | |
| **NBM volume** | 478 | 342/109 | 1759±180 | 1715±166 | 0·999 (0·997; 1·000) | |
| **Cerebral white matter volume (cm³)** | 492 | 331/106 | 438±60 | 428±55 | 0·997 (0·993; 1·001) | |
| **Cerebral cortex volume (cm³)** | 492 | 331/106 | 411±42 | 400±42 | 0·993 (0·988; 0·999) | |
| **Lateral Ventricle volume (cm³)** | 492 | 331/106 | 18·0±8·2 | 20·1±11·2 | 1·025 (0·999; 1·051) | |
| **Inferiolateral ventricle volume (cm³)** | 492 | 331/106 | 0·75±0·44 | 0·82±0·42 | 1·405 (0·843; 2·431) | |
| **Third ventricle volume (cm³)** | 492 | 331/106 | 1·95±0·68 | 2·08±0·59 | 1·307 (0·922; 1·854) | |
| **Fourth ventricle volume (cm³)** | 492 | 331/106 | 1·99±0·56 | 2·08±0·63 | 1·299 (0·863; 1·954) | |
| **Cerebellar white matter volume (cm³)** | 492 | 331/106 | 13·0±2·3 | 12·5±2·6 | 0·910 (0·815; 1·018) | |
| **Cerebellar cortex volume (cm³)** | 492 | 331/106 | 49·7±5·2 | 47·4±5·5 | *0·916 (0·872; 0·961)* | *0·63 (0·49; 0·81)* |
| **Thalamus volume** | 492 | 331/106 | 6347±738 | 6001±743 | *0·9993 (0·9990; 0·9997)* | *0·61 (0·47; 0·80)* |
| **Caudate nucleus volume** | 492 | 331/106 | 3361±546 | 3286±591 | 0·9997 (0·9993; 1·0002) | |
| **Putamen volume** | 492 | 331/106 | 4094±613 | 3929±538 | *0·9995 (0·9991; 0·9999)* | *0·75 (0·58; 0·97)* |
| **Pallidum volume** | 492 | 331/106 | 1763±292 | 1715±291 | 0·9994 (0·9986; 1·0003) | |
| **Hippocampus volume** | 492 | 331/106 | 3671±422 | 3504±431 | *0·9991 (0·9985; 0·9996)* | *0·67 (0·53; 0·89)* |
| **Amygdala volume** | 492 | 331/106 | 1369±248 | 1324±236 | 0·999 (0·998; 1·000) | |
| **Nucleus accumbens volume** | 492 | 331/106 | 397±101 | 367±107 | *0·997 (0·994; 0·999)* | *0·73 (0·57; 0·95)* |

| | | N | Missing $n_{NPOD}/n_{POD}$ | No POD | POD | OR (95% CI) | Standard-ized OR |
|---|---|---|---|---|---|---|---|
| **Brain stem volume (cm³)** | | 492 | 331/106 | 20·0±2·6 | 19·1±2·6 | *0·863 (0·780; 0·955)* | *0·68 (0·53; 0·89)* |
| **CA1** | | 309 | 483/137 | 2288±302 | 2263±313 | 1·000 (0·999; 1·001) | |
| **CA2** | | 309 | 483/137 | 35·10±9·02 | 35·65±10·42 | 1·007 (0·973; 1·042) | |
| **Dentate gyrus** | | 309 | 483/137 | 1427±203 | 1427±219 | 1·000 (0·998; 1·001) | |
| **Subiculum** | | 309 | 483/137 | 637±85 | 638±97 | 1·000 (0·996; 1·004) | |
| **High res. entorhinal cortex** | | 309 | 483/137 | 800±129 | 794±144 | 1·000 (0·997; 1·002) | |
| **Rostral anterior cin-gulate cortex** | Thickness | 492 | 331/106 | 2·782±0·207 | 2·742±0·177 | 0·37 (0·11; 1·25) | |
| | Mean cur-vature | 492 | 331/106 | 13·66±1·10 | 13·68±0·83 | 1·02 (0·81; 1·28) | |
| | Volume | 492 | 331/106 | 1971±387 | 1934±361 | 0·9997 (0·9991; 1·0004) | |
| **Caudal anterior cin-gulate cortex** | Thickness | 492 | 331/106 | 2·551±0·216 | 2·512±0·219 | 0·43 (0·14; 1·33) | |
| | Mean cur-vature | 492 | 331/106 | 13·17±1·15 | 13·22±1·24 | 1·04 (0·84; 1·29) | |
| | Volume | 492 | 331/106 | 1634±305 | 1620±296 | 0·9998 (0·9990; 1·0006) | |
| **Posterior cingulate cortex** | Thickness | 492 | 331/106 | 2·411±0·146 | 2·352±0·172 | *0·08 (0·02; 0·40)* | *0·68 (0·54; 0·87)* |
| | Mean cur-vature | 492 | 331/106 | 13·91±0·88 | 13·75±1·03 | 0·83 (0·64; 1·08) | |
| | Volume | 492 | 331/106 | 2790±400 | 2684±654 | 0·9994 (0·9989; 1·0000) | |
| **Isthmus cinguli** | Thickness | 492 | 331/106 | 2·263±0·173 | 2·194±0·196 | *0·11 (0·03; 0·44)* | *0·67 (0·53; 0·86)* |
| | Mean cur-vature | 492 | 331/106 | 12·73±0·80 | 12·67±0·90 | 0·92 (0·69; 1·23) | |
| | Volume | 492 | 331/106 | 2242±336 | 2175±466 | 0·9995 (0·9987; 1·0001) | |
| **Superior parietal gy-rus** | Thickness | 492 | 331/106 | 2·065±0·137 | 2·035±0·130 | 0·20 (0·03; 1·21) | |
| | Mean cur-vature | 492 | 331/106 | 11·61±0·82 | 11·77±0·77 | 1·27 (0·95; 1·71) | |
| | Volume | 492 | 331/106 | 11012±1416 | 10818±1409 | 0·9999 (0·9997; 1·0001) | |
| **Inferior parietal gy-rus** | Thickness | 492 | 331/106 | 2·310±0·129 | 2·282±0·125 | 0·18 (0·03; 1·18) | |
| | Mean cur-vature | 492 | 331/106 | 12·50±0·78 | 12·61±0·68 | 1·21 (0·88; 1·65) | |
| | Volume | 492 | 331/106 | 11318±1633 | 10923±1491 | *0·9998 (0·9997; 1·0000)* | *0·78 (0·60; 1·00)* |

| | | N | Missing $n_{NPOD}/n_{POD}$ | No POD | POD | OR (95%CI) | Standardized OR |
|---|---|---|---|---|---|---|---|
| **Insula** | Thickness | 492 | 331/106 | 2·811±0·164 | 2·759±0·172 | *0·15 (0·04; 0·65)* | *0·73 (0·58; 0·93)* |
| | Mean curvature | 492 | 331/106 | 12·24±0·87 | 12·21±0·85 | 0·96 (0·72; 1·27) | |
| | Volume | 492 | 331/106 | 6593±793 | 6373±796 | *0·9996 (0·9993; 1·0000)* | *0·76 (0·59; 0·97)* |
| **Supramarginal gyrus** | Thickness | 492 | 331/106 | 2·378±0·124 | 2·343±0·127 | *0·11 (0·02; 0·74)* | *0·76 (0·59; 0·97)* |
| | Mean curvature | 492 | 331/106 | 12·57±0·63 | 12·63±0·63 | 1·16 (0·79; 1·69) | |
| | Volume | 492 | 331/106 | 9192±1294 | 8825±1162 | 0·9998 (0·9996; 1·0000) | |
| **Precentral gyrus** | Thickness | 492 | 331/106 | 2·395±0·168 | 2·342±0·164 | *0·15 (0·04; 0·63)* | *0·73 (0·57; 0·93)* |
| | Mean curvature | 492 | 331/106 | 10·51±0·76 | 10·51±0·75 | 1·01 (0·73; 1·39) | |
| | Volume | 492 | 331/106 | 11840±1428 | 11308±1491 | *0·9997 (0·9996; 0·9999)* | *0·69 (0·54; 0·89)* |
| **Postcentral gyrus** | Thickness | 492 | 331/106 | 1·914±0·114 | 1·902±0·111 | 0·40 (0·05; 3·40) | |
| | Mean curvature | 492 | 331/106 | 10·58±0·69 | 10·70±0·69 | 1·22 (0·87; 1·72) | |
| | Volume | 492 | 331/106 | 8048±1042 | 7915±1048 | 0·9999 (0·9996; 1·0001) | |
| **Paracentral gyrus** | Thickness | 492 | 331/106 | 2·298±0·160 | 2·384±0·152 | 0·56 (0·12; 2·57) | |
| | Mean curvature | 492 | 331/106 | 10·20±1·00 | 10·38±1·10 | 1·18 (0·93; 1·48) | |
| | Volume | 492 | 331/106 | 3362±439 | 3341±448 | 0·9999 (0·9950; 1·0023) | |
| **Lateral occipital cortex** | Thickness | 492 | 331/106 | 2·095±0·121 | 2·064±0·105 | *0·10 (0·01; 0·82)* | *0·76 (0·59; 0·99)* |
| | Mean curvature | 492 | 331/106 | 13·79±0·77 | 13·93±0·74 | 1·26 (0·92; 1·72) | |
| | Volume | 492 | 331/106 | 10439±1473 | 10300±1430 | 0·9999 (0·9998; 1·0001) | |
| **Lingual gyrus** | Thickness | 492 | 331/106 | 1·924±0·101 | 1·898±0·104 | *0·08 (0·01; 0·85)* | *0·77 (0·60; 0·98)* |
| | Mean curvature | 492 | 331/106 | 13·93±0·75 | 14·05±0·73 | 1·24 (0·90; 1·72) | |
| | Volume | 492 | 331/106 | 5855±838 | 5751±797 | 0·9998 (0·9995; 1·0001) | |
| **Pericalcarine cortex** | Thickness | 492 | 331/106 | 1·593±0·103 | 1·570±0·103 | 0·11 (0·01; 1·24) | |
| | Mean curvature | 492 | 331/106 | 12·88±0·93 | 12·97±0·98 | 1·07 (0·85; 1·42) | |
| | Volume | 492 | 331/106 | 2031±389 | 2013±380 | 0·9999 (0·9992; 1·0005) | |

| | | N | Missing $n_{NPOD}/n_{POD}$ | No POD | POD | OR (95% CI) | Standard-ized OR |
|---|---|---|---|---|---|---|---|
| **Cuneus** | Thickness | 492 | 331/106 | 1·805±0·109 | 1·811±0·091 | 1·71 (0·17; 16·25) | |
| | Mean curvature | 492 | 331/106 | 13·89±0·86 | 14·19±0·93 | *1·47 (1·12; 1·94)* | *1·41 (1·10; 1·79)* |
| | Volume | 492 | 331/106 | 2761±423 | 2762±388 | 1·0000 (0·9994; 1·0006) | |
| **Precuneus** | Thickness | 492 | 331/106 | 2·247±0·122 | 2·212±±0·124 | *0·11 (0·01; 0·77)* | *0·76 (0·59; 0·97)* |
| | Mean curvature | 492 | 331/106 | 12·73±0·59 | 12·96±0·56 | *1·93 (1·28; 2·91)* | *1·47 (1·15; 1·88)* |
| | Volume | 492 | 331/106 | 8434±1067 | 8196±988 | 0·9998 (0·9995; 1·0000) | |
| **Superior frontal gyrus** | Thickness | 492 | 331/106 | 2·555±0·140 | 2·528±0·137 | *0·24 (0·04; 1·40)* | *0·77 (0·60; 0·98)* |
| | Mean curvature | 492 | 331/106 | 12·35±0·73 | 12·31±0·72 | 0·93 (0·66; 1·30) | |
| | Volume | 492 | 331/106 | 19144±2308 | 18613±2375 | 0·9999 (0·9998; 1·0000) | |
| **Rostral middle frontal gyrus** | Thickness | 492 | 331/106 | 2·236±0·118 | 2·207±0·112 | 0·12 (0·02; 0·98) | |
| | Mean curvature | 492 | 331/106 | 13·69±0·84 | 13·60±0·74 | 0·88 (0·65; 1·18) | |
| | Volume | 492 | 331/106 | 13232±1812 | 13013±1878 | 0·9999 (0·9998; 1·0001) | |
| **Caudal middle frontal gyrus** | Thickness | 492 | 331/106 | 2·429±0·151 | 2·389±0·161 | *0·18 (0·04; 0·87)* | *0·78 (0·61; 1·00)* |
| | Mean curvature | 492 | 331/106 | 11·32±0·86 | 22·29±0·77 | 0·97 (0·72; 1·29) | |
| | Volume | 492 | 331/106 | 5346±788 | 5159±877 | 0·9997 (0·9994; 1·0000) | |
| **Frontal pole** | Thickness | 492 | 331/106 | 2·627±0·229 | 2·576±0·231 | 0·37 (0·12; 1·07) | |
| | Mean curvature | 492 | 331/106 | 17·51±1·40 | 17·78±1·40 | 1·16 (0·97; 1·38) | |
| | Volume | 492 | 331/106 | 996±150 | 971±147 | 0·9989 (0·9971; 1·0005) | |
| **Medial orbitofrontal cortex** | Thickness | 492 | 331/106 | 2·426±0·166 | 2·388±0·174 | *0·23 (0·05; 1·04)* | *0·70 (0·55; 0·90)* |
| | Mean curvature | 492 | 331/106 | 13·83±0·72 | 13·80±0·76 | 0·92 (0·66; 1·30) | |
| | Volume | 492 | 331/106 | 4691±552 | 4566±559 | 0·9996 (0·9991; 1·0005) | |
| **Lateral orbitofrontal cortex** | Thickness | 492 | 331/106 | 2·618±0·140 | 2·569±0·148 | 0·08 (0·01; 0·45) | |
| | Mean curvature | 492 | 331/106 | 13·94±0·61 | 14·01±0·56 | 1·21 (0·81; 1·81) | |
| | Volume | 492 | 331/106 | 6573±756 | 6440±767 | 0·9998 (0·9994; 1·0001) | |

| | | N | Missing $n_{NPOD}/n_{POD}$ | No POD | POD | OR (95% CI) | Standard-ized OR |
|---|---|---|---|---|---|---|---|
| **Pars opercularis** | Thickness | 492 | 331/106 | 2·446±2·408 | 2·408±0·132 | *0·10 (0·01; 0·64)* | *0·74 (0·58; 0·95)* |
| | Mean curvature | 492 | 331/106 | 11·80±0·67 | 11·80±0·75 | 1·00 (0·70; 1·43) | |
| | Volume | 492 | 331/106 | 3712±537 | 3587±530 | 0·9995 (0·9990; 1·0000) | |
| **Pars orbitalis** | Thickness | 492 | 331/106 | 2·564±0·163 | 2·512±0·153 | *0·13 (0·03; 0·59)* | *0·72 (0·56; 0·92)* |
| | Mean curvature | 492 | 331/106 | 14·59±0·96 | 14·62±0·91 | 1·04 (0·80; 1·33) | |
| | Volume | 492 | 331/106 | 2240±288 | 2130±301 | *0·9986 (0·9977; 0·9995)* | *0·67 (0·51; 0·87)* |
| **Pars triangularis** | Thickness | 492 | 331/106 | 2·305±0·134 | 2·283±0·130 | 0·28 (0·04; 1·76) | |
| | Mean curvature | 492 | 331/106 | 12·59±0·80 | 12·59±0·71 | 1·00 (0·74; 1·37) | |
| | Volume | 492 | 331/106 | 3321±460 | 3204±498 | *0·9994 (0·9989; 1·0000)* | *0·77 (0·60; 0·99)* |
| **Superior temporal gyrus** | Thickness | 492 | 331/106 | 2·611±0·148 | 2·567±0·148 | 0·14 (0·03; 0·69) | |
| | Mean curvature | 492 | 331/106 | 10·88±0·66 | 10·85±0·60 | 0·93 (0·64; 1·35) | |
| | Volume | 492 | 331/106 | 10476±1304 | 10170±1284 | 0·9998 (0·9996; 1·0000) | |
| **Superior temporal sulcus** | Thickness | 492 | 331/106 | 2·424±0·135 | 2·395±0·145 | *0·22 (0·04; 1·27)* | *0·74 (0·58; 0·95)* |
| | Mean curvature | 492 | 331/106 | 10·62±0·99 | 10·96±0·92 | *1·42 (1·12; 1·83)* | *1·42 (1·11; 1·81)* |
| | Volume | 492 | 331/106 | 2000±308 | 1972±311 | 0·9997 (0·9989; 1·2689) | |
| **Middle temporal gyrus** | Thickness | 492 | 331/106 | 2·714±0·123 | 2·676±0·128 | *0·08 (0·01; 0·59)* | *0·74 (0·57; 0·94)* |
| | Mean curvature | 492 | 331/106 | 13·30±0·68 | 13·36±0·64 | 1·14 (0·80; 1·63) | |
| | Volume | 492 | 331/106 | 9789±1292 | 9567±1236 | 0·9999 (0·9997; 1·0001) | |
| **Inferior temporal gyrus** | Thickness | 492 | 331/106 | 2·651±0·123 | 2·623±0·128 | 0·16 (0·02; 1·11) | |
| | Mean curvature | 492 | 331/106 | 13·85±0·74 | 14·04±0·75 | *1·39 (1·01; 1·92)* | *1·28 (1·01; 1·63)* |
| | Volume | 492 | 331/106 | 9279±1286 | 8986±1286 | 0·9998 (0·9996; 1·0000) | |
| **Temporal pole** | Thickness | 492 | 331/106 | 3·616±0·242 | 3·565±0·252 | 0·43 (0·16; 1·15) | |
| | Mean curvature | 492 | 331/106 | 14·94±1·32 | 15·03±1·22 | 1·06 (0·88; 1·27) | |
| | Volume | 492 | 331/106 | 2121±330 | 2401±338 | 0·9998 (0·9991; 1·0005) | |

| | | N | Missing $n_{NPOD}/n_{POD}$ | No POD | POD | OR (95% CI) | Standard-ized OR |
|---|---|---|---|---|---|---|---|
| **Temporal trans-verse gyrus** | Thickness | 492 | 331/106 | 2·223±0·205 | 2·188±0·226 | 0·44 (0·14; 1·41) | |
| | Mean cur-vature | 492 | 331/106 | 10·31±1·14 | 10·37±1·03 | 1·05 (0·85; 1·30) | |
| | Volume | 492 | 331/106 | 860±150 | 833±154 | 0·9988 (0·9971; 1·0004) | |
| **Fusiform gyrus** | Thickness | 492 | 331/106 | 2·617±0·131 | 2·574±0·131 | *0·08 (0·01; 0·51)* | *0·72 (0·57; 0·92)* |
| | Mean cur-vature | 492 | 331/106 | 13·64±0·65 | 13·87±0·74 | *1·63 (1·14; 2·33)* | *0·71 (0·55; 0·92)* |
| | Volume | 492 | 331/106 | 8272±1110 | 7913±984 | *0·9997 (0·9995; 0·9999)* | *1·39 (1·09; 1·76)* |
| **Entorhinal cortex** | Thickness | 492 | 331/106 | 3·350±0·297 | 3·318±0·298 | 0·70 (0·31; 1·59) | |
| | Mean cur-vature | 492 | 331/106 | 12·52±1·41 | 12·51±1·28 | 0·99 (0·83; 1·18) | |
| | Volume | 492 | 331/106 | 2024±382 | 1920±363 | *0·9992 (0·9986; 0·9999)* | *0·75 (0·58; 0·97)* |
| **Parahippocampal gyrus** | Thickness | 492 | 331/106 | 2·665±0·253 | 2·612±0·251 | 0·45 (0·17; 1·17) | |
| | Mean cur-vature | 492 | 331/106 | 9·59±1·15 | 9·65±1·29 | 1·05 (0·85; 1·29) | |
| | Volume | 492 | 331/106 | 1877±259 | 1832±271 | 0·9993 (0·9984; 1·0003) | |
| **Ant. thalamic radia-tion L** | FA | 326 | 473/130 | 39·6±3·2 | 38·2±4·8 | *0·90 (0·83; 0·97)* | *0·69 (0·52; 0·91)* |
| | MD | 326 | 473/130 | 1108±150 | 1220±263 | *1·003 (1·002; 1·005)* | *1·68 (1·29; 2·20)* |
| | Kurtosis | 326 | 473/130 | 74·5±6·6 | 71·4±8·0 | 0·94 (0·90; 0·98) | |
| | Volume | 326 | 473/130 | 7454±1231 | 7691±1360 | 1·0001 (0·9999; 1·0004) | |
| **Ant. thalamic ration R** | FA | 326 | 473/130 | 35·6±3·2 | 34·4±4·1 | *0·90 (0·83; 0·98)* | *0·71 (0·53; 0·94)* |
| | MD | 326 | 473/130 | 1177±166 | 1264±229 | *1·002 (1·001; 1·004)* | *1·53 (1·17; 2·00)* |
| | Kurtosis | 326 | 473/130 | 65·0±7·2 | 63·6±7·0 | 0·97 (0·93; 1·01) | |
| | Volume | 326 | 473/130 | 6876±1186 | 7060±1192 | 1·0001 (0·9999; 1·0004) | |
| **Corticospinal tract L** | FA | 326 | 473/130 | 58·2±3·1 | 58·7±3·5 | 1·06 (0·96; 1·16) | |
| | MD | 326 | 473/130 | 914±50 | 920±60 | 1·002 (0·996; 1·007) | |
| | Kurtosis | 326 | 473/130 | 98·0±6·9 | 95·8±7·1 | *0·96 (0·92; 0·98)* | *0·75 (0·57; 0·98)* |
| | Volume | 326 | 473/130 | 4376±571 | 4319±511 | 0·9998 (0·9993; 1·0003) | |

| | | N | Missing $n_{NPOD}/n_{POD}$ | No POD | POD | OR (95% CI) | Standard-ized OR |
|---|---|---|---|---|---|---|---|
| **Corticospinal tract R** | FA | 326 | 473/130 | 59·6±3·2 | 60·1±3·9 | 1·05 (0·96; 1·14) | |
| | MD | 326 | 473/130 | 895±47 | 899±56 | 1·001 (0·995; 1·007) | |
| | Kurtosis | 326 | 473/130 | 95·00±7·4 | 93·5±7·6 | 0·97 (0·94; 1·01) | |
| | Volume | 326 | 473/130 | 3890±495 | 3817±409 | 0·9997 (0·9990; 1·0003) | |
| **Cingulate gyrus L** | FA | 326 | 473/130 | 46·8±5·5 | 44·7±6·4 | *0·94 (0·89; 0·99)* | *1·45 (1·07; 1·97)* |
| | MD | 326 | 473/130 | 925±54 | 987±329 | *1·005 (1·001; 1·010)* | *2·10 (1·02; 4·33)* |
| | Kurtosis | 326 | 473/130 | 78·5±10·3 | 75·9±10·7 | 0·98 (0·95; 1·00) | |
| | Volume | 326 | 473/130 | 1522±215 | 1506±214 | 0·9996 (0·9982; 1·0010) | |
| **Cingulate gyrus R** | FA | 326 | 473/130 | 43·1±6·4 | 41·3±7·2 | 0·96 (0·92; 1·01) | |
| | MD | 326 | 473/130 | 891±68 | 954±351 | 1·003 (1·000; 1·007) | |
| | Kurtosis | 326 | 473/130 | 70·6±11·8 | 70·7±12·2 | 1·00 (0·98; 0·917) | |
| | Volume | 326 | 473/130 | 764±119 | 758±214 | 0·9995 (0·9970; 1·0020) | |
| **Hippocampal cingu-lum L** | FA | 326 | 473/130 | 42·9±5·2 | 44·7±5·5 | *1·08 (1·01; 1·14)* | *0·69 (0·52; 0·93)* |
| | MD | 326 | 473/130 | 904±118 | 897±134 | 0·999 (0·997; 1·002) | |
| | Kurtosis | 326 | 473/130 | 57·4±11·3 | 57·4±12·9 | 1·00 (0·98; 1·03) | |
| | Volume | 326 | 473/130 | 444±74 | 442±58 | 0·9997 (0·9955; 1·0037) | |
| **Hippocampal cingu-lum R** | FA | 326 | 473/130 | 47·8±5·7 | 47·2±5·7 | 0·98 (0·93; 1·03) | |
| | MD | 326 | 473/130 | 854±90 | 886±153 | 1·002 (1·000; 1·005) | |
| | Kurtosis | 326 | 473/130 | 55·5±11·0 | 58·5±10·7 | 1·02 (1·00; 1·05) | |
| | Volume | 326 | 473/130 | 637±93 | 631±77 | 0·9994 (0·9961; 1·0026) | |
| **Forceps major** | FA | 326 | 473/130 | 55·9±4·7 | 54·3±6·2 | 0·94 (0·89; 1·00) | |
| | MD | 326 | 473/130 | 1093±105 | 1150±223 | 1·003 (1·001; 1·005) | |
| | Kurtosis | 326 | 473/130 | 75·4±7·7 | 74·9±8·9 | 0·99 (0·97; 1·03) | |
| | Volume | 326 | 473/130 | 6029±898 | 6157±934 | 1·0002 (0·9998; 1·0005) | |

| | | N | Missing $n_{NPOD}/n_{POD}$ | No POD | POD | OR (95% CI) | Standard-ized OR |
|---|---|---|---|---|---|---|---|
| **Forceps minor** | FA | 326 | 473/130 | 40·3±3·3 | 39·2±4·3 | *0·91 (0·84; 0·99)* | *0·74 (0·56; 0·97)* |
| | MD | 326 | 473/130 | 1001±65 | 1033±121 | *1·004 (1·001; 1·008)* | *1·40 (1·06; 1·84)* |
| | Kurtosis | 326 | 473/130 | 74·2±5·8 | 73·0±7·2 | 0·97 (0·92; 1·02) | |
| | Volume | 326 | 473/130 | 15401±2066 | 15308±2120 | 1·0000 (0·9998; 1·0001) | |
| **Inf. fronto-occipital fasc. L** | FA | 326 | 473/130 | 44·5±2·9 | 43·4±4·1 | *0·90 (0·82; 0·98)* | *0·71 (0·53; 0·94)* |
| | MD | 326 | 473/130 | 979±53 | 1008±131 | *1·004 (1·001; 1·008)* | *1·36 (1·04; 1·78)* |
| | Kurtosis | 326 | 473/130 | 83·7±5·2 | 81·5±7·1 | *0·94 (0·89; 0·99)* | *0·70 (0·52; 0·93)* |
| | Volume | 326 | 473/130 | 5788±730 | 5790±782 | 1·0000 (0·9996; 1·0004) | |
| **Inf. fronto-occipital fasc. R** | FA | 326 | 473/130 | 44·8±3·1 | 44·2±3·2 | 0·94 (0·85; 1·03) | |
| | MD | 326 | 473/130 | 967±53 | 980±63 | 1·004 (1·000; 1·10) | |
| | Kurtosis | 326 | 473/130 | 82·9±5·4 | 82·3±6·2 | 0·98 (0·93; 1·03) | |
| | Volume | 326 | 473/130 | 6435±824 | 6376±832 | 0·9999 (0·9995; 1·0003) | |
| **Inf. longitudinal fasc. L** | FA | 326 | 473/130 | 44·8±3·1 | 39·5±2·9 | 0·96 (0·86; 1·07) | |
| | MD | 326 | 473/130 | 964±49 | 970±61 | 1·002 (0·996; 1·008) | |
| | Kurtosis | 326 | 473/130 | 81·5±5·4 | 80·5±6·2 | 0·97 (0·92; 1·02) | |
| | Volume | 326 | 473/130 | 6082±861 | 5995±795 | 0·9999 (0·9995; 1·0003) | |
| **Inf. longitudinal fasc. R** | FA | 326 | 473/130 | 44·1±2·8 | 44·6±2·9 | 1·06 (0·96; 1·18) | |
| | MD | 326 | 473/130 | 949±49 | 949±60 | 1·000 (0·994; 1·006) | |
| | Kurtosis | 326 | 473/130 | 86·2±6·1 | 85·9±47·1 | 0·99 (0·95; 1·04) | |
| | Volume | 326 | 473/130 | 3786±512 | 3744±554 | 0·9998 (0·9993; 1·0004) | |
| **Sup. longitudinal fasc. L** | FA | 326 | 473/130 | 38·6±2·7 | 37·8±3·4 | 0·91 (0·83; 1·01) | |
| | MD | 326 | 473/130 | 1042±80 | 1062±82 | 1·003 (1·00; 1·01) | |
| | Kurtosis | 326 | 473/130 | 89·9±6·3 | 88·5±7·1 | 0·97 (0·93; 1·01) | |
| | Volume | 326 | 473/130 | 9183±1104 | 8919±1096 | 0·9998 (0·9995; 1·0000) | |

| | | N | Missing $n_{NPOD}/n_{POD}$ | No POD | POD | OR (95% CI) | Standard-ized OR |
|---|---|---|---|---|---|---|---|
| **Sup. longitudinal fasc. R** | FA | 326 | 473/130 | 41·9±3·4 | 40·8±4·8 | *0·93 (0·86; 1·00)* | *0·76 (0·58; 1·00)* |
| | MD | 326 | 473/130 | 956±65 | 979±100 | *1·004 (1·000; 1·008)* | *1·31 (1·00; 1·71)* |
| | Kurtosis | 326 | 473/130 | 93·1±6·9 | 91·1±9·0 | 0·97 (0·93; 1·00) | |
| | Volume | 326 | 473/130 | 7153±900 | 6869±860 | *0·9996 (0·9993; 1·0000)* | *0·72 (0·53; 0·98)* |
| **Uncinate fasc. L** | FA | 326 | 473/130 | 45·0±3·8 | 44·0±3·4 | 0·93 (0·86; 1·01) | |
| | MD | 326 | 473/130 | 992±70 | 1004±96 | 1·002 (0·998; 1·006) | |
| | Kurtosis | 326 | 473/130 | 73·0±6·7 | 70·8±7·5 | *0·95 (0·91; 0·99)* | *0·72 (0·53; 0·96)* |
| | Volume | 326 | 473/130 | 1274±164 | 1260±179 | 0·9995 (0·9977; 1·0012) | |
| **Uncinate fasc. R** | FA | 326 | 473/130 | 47·9±4·7 | 46·4±3·5 | 0·93 (0·87; 0·99) | |
| | MD | 326 | 473/130 | 938±81 | 970±84 | *1·004 (1·001; 1·008)* | *1·40 (1·07; 1·84)* |
| | Kurtosis | 326 | 473/130 | 65·8±8·9 | 65·4±8·1 | 0·99 (0·96; 1·03) | |
| | Volume | 326 | 473/130 | 562±84 | 564±77 | 1·0003 (0·9967; 1·0038) | |
| **Sup. longitudinal fasc. (temporal) L** | FA | 326 | 473/130 | 49·4±3·5 | 48·6±4·7 | 0·95 (0·88; 1·02) | |
| | MD | 326 | 473/130 | 919±59 | 637±84 | 1·004 (1·000; 1·008) | |
| | Kurtosis | 326 | 473/130 | 98·5±6·9 | 97·0±8·8 | 0·97 (0·93; 1·01) | |
| | Volume | 326 | 473/130 | 2769±382 | 2655±342 | *0·9992 (0·9983; 1·0000)* | *0·73 (0·53; 0·99)* |
| **Sup. longitudinal fasc. (temporal) R** | FA | 326 | 473/130 | 51·3±4·4 | 50·0±6·6 | 0·95 (0·90; 1·01) | |
| | MD | 326 | 473/130 | 887±62 | 907±110 | 1·003 (1·000; 1·007) | |
| | Kurtosis | 326 | 473/130 | 100·6±7·8 | 98·6±10·6 | 0·97 (0·94; 1·01) | |
| | Volume | 326 | 473/130 | 1500±225 | 1444±209 | 0·9989 (0·9975; 1·0002) | |

**eTable 16: Mean area under the curve (AUC), and calibration (Brier score) with 95% confidence intervals (CI) for all models.**

* These models were calculated for the subsample of patients with RNA data for the purpose to compare model perfomances in this subgroup of patients.

| Dataset combination for model building | AUC | | Brier Score | |
|---|---|---|---|---|
| | Mean | 95%CI | Mean | 95%CI |
| **Models using exclusively preoperative data for prediction in the whole BioCog cohort** | | | | |
| **Clinical** | 0·76 | (0·69; 0·81) | 0·14 | (0·13; 0·16) |
| **Clinical + Blood** | 0.73 | (0.68; 0.79) | 0.15 | (0.14; 0.15) |
| **Blood** | 0·61 | (0·54; 0·68) | 0·18 | (0·16; 0·20) |
| **Imaging** | 0·58 | (0·54; 0·62) | 0·23 | (0·22; 0·24) |
| **Model using preoperative data and precipitating factors in the whole BioCog cohort** | | | | |
| **Clinical + Precipitants + Blood periop.** | 0·83 | (0·79; 0·86) | 0·12 | (0·12; 0·13) |
| **Clinical + Precipitants** | 0·80 | (0·77, 0·84) | 0·13 | (0·12, 0·14) |
| **Clinical + Blood periop.** | 0·79 | (0·75, 0·82) | 0·13 | (0·13, 0·14) |
| **Clinical + Pain** | 0·76 | (0·72, 0·80) | 0·14 | (0·13, 0·15) |
| **Blood periop.** | 0·74 | (0·68; 0·77) | 0·18 | (0·16; 0·20) |
| **Precipitants** | 0·71 | (0·68; 0·75) | 0·14 | (0·14; 0·15) |
| **Pain** | 0·58 | (0·56; 0·6) | 0·17 | (0·16; 0·18) |
| **Models using exclusively preoperative data for prediction in the BioCog RNA subsample** | | | | |
| **Clinical (for comparison*)** | 0·69 | (0·65; 0·74) | 0·16 | (0·16; 0·18) |
| **RNA + Clinical** | 0·66 | (0·61; 0·71) | 0·17 | (0·16; 0·18) |
| **RNA** | 0·66 | (0·62; 0·70) | 0·17 | (0·16; 0·17) |
| **µRNA** | 0·47 | (0·40; 0·54) | 0·20 | (0·19; 0·22) |
| **Model using preoperative data and precipitating factors in the whole BioCog subsample** | | | | |
| **Clinical + Precipitants + Blood periop· (for comparison*)** | 0·78 | (0·73; 0·83) | 0·15 | (0·14; 0·16) |
| **Clinical + Precipitants + Blood periop. + RNA + RNA periop.** | 0·77 | (0·72; 0·83) | 0·15 | (0·14; 0·16) |
| **Clinical + Precipitants + RNA periop.** | 0·74 | (0·71; 0·78) | 0·16 | (0·16; 0·17) |
| **RNA + RNA periop.** | 0·77 | (0·72; 0·82) | 0·15 | (0·14; 0·16) |
| **RNA periop.** | 0·73 | (0·69; 0·78) | 0·16 | (0·15; 0·17) |
| **µRNA periop.** | 0·60 | (0·54; 0·66) | 0·20 | (0·19; 0·21) |

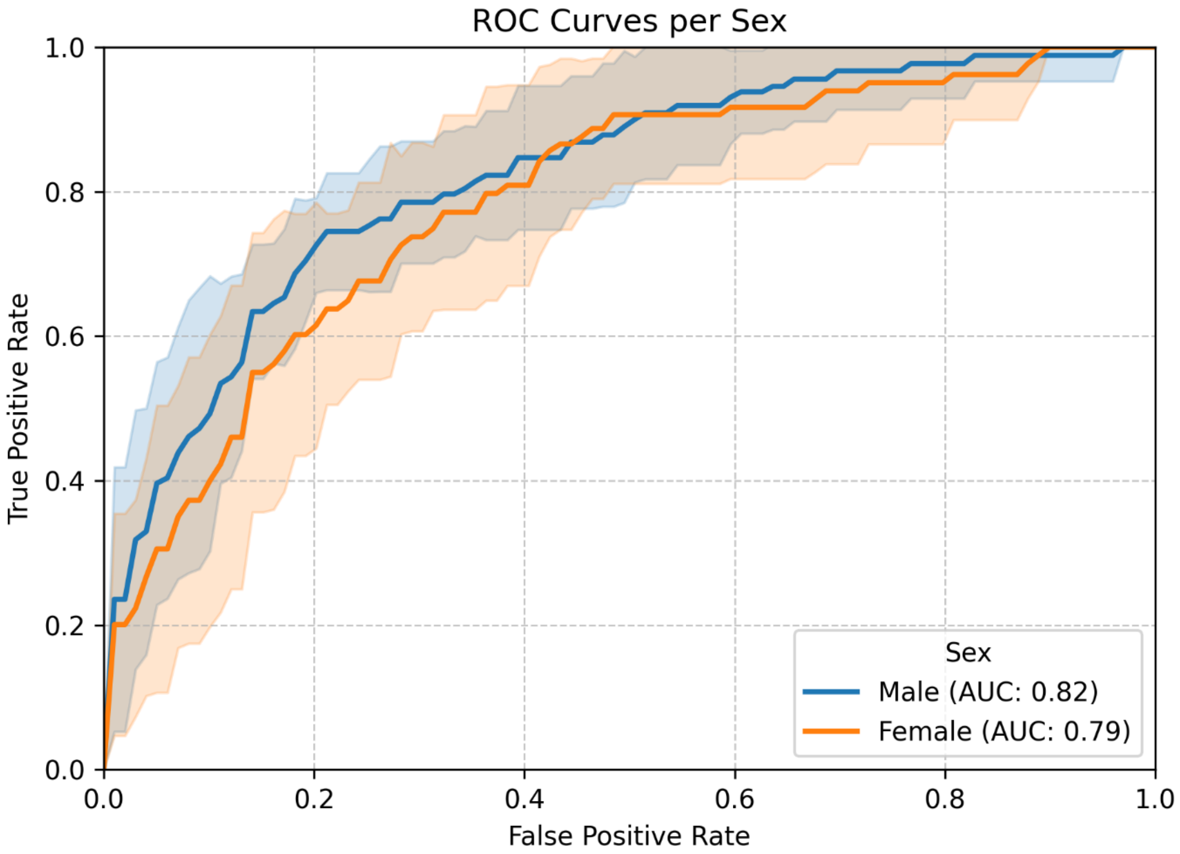


**eFigure 9: Receiver-Operating Curves for the best-performing model ("Clinical + Precipitants + Blood Periop.") stratified by sex.**

The model is equally suitable for both female and male patients.

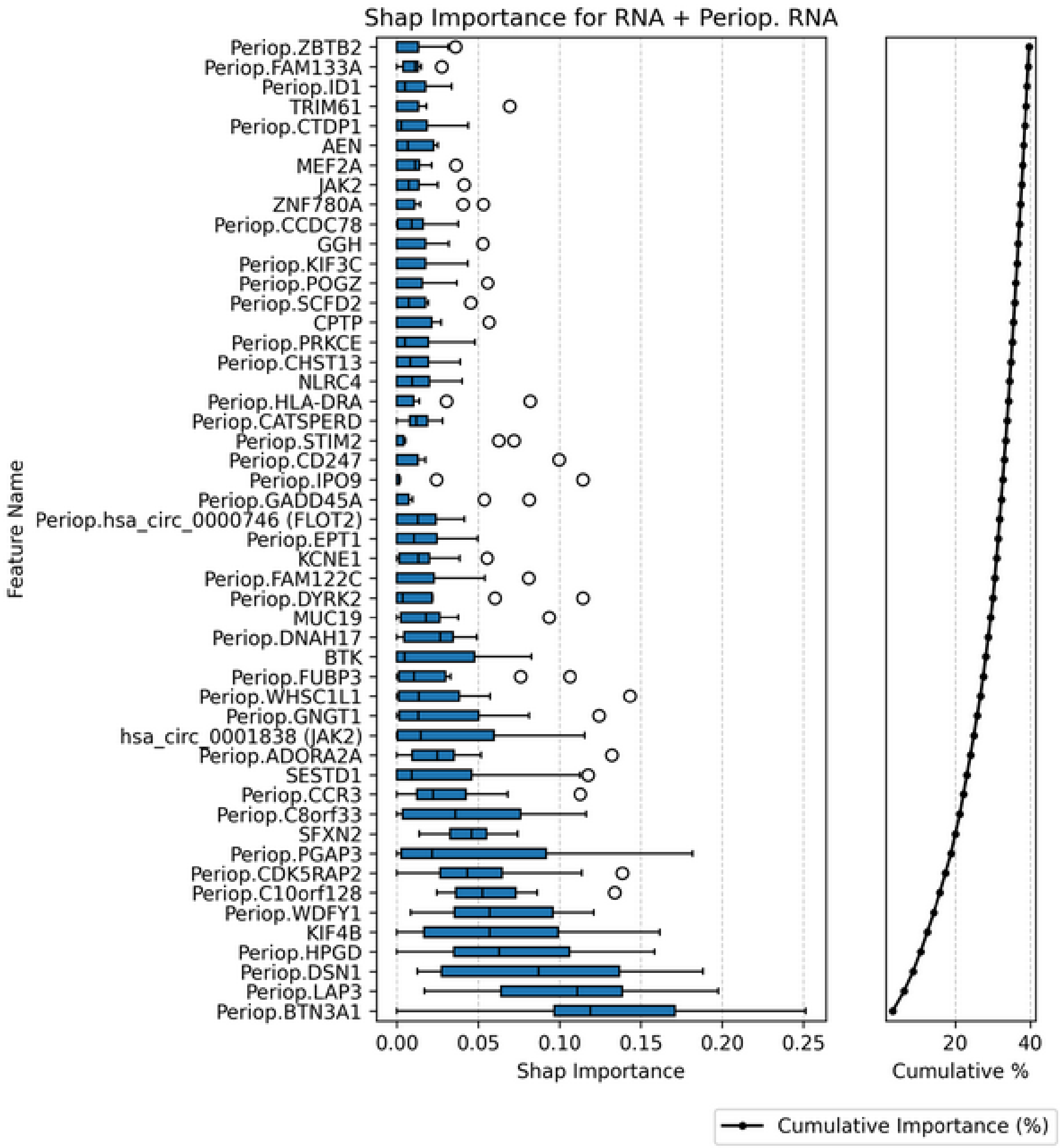


**EFIGURE 10: FEATURE IMPORTANCE IN THE SECOND-BEST PREDICTIVE MODEL ON TRANSCRIPTOMIC DATA**

#### eChapter 2.3: Associations between transcript abundance and mortality

**eTable 17: Associations between mRNA and mortality until 90 days after surgery.**

Cox regression of seven transcripts adjusted for age, sex and Charlson's Comorbidity Index has been performed to analyze association between relevant transcripts from the gradient-boosted trees model of POD and postoperative mortality. P-values have not been adjusted for multiple tests. 503 patients,who provided preoperative mRNA data and of whom 22 died until the 90th postoperative day, were included in the analysis of KIF4B and JAK2. Of 374 patients providing both pre- and postoperative mRNA data for analysis of HPGD, BTN3A1, LAP3 and DSN1, 17 died until the 90th postoperative day.

| Transcript | Regression coefficient (95%CI) |
|---|---|
| **Perioperative HPGD (15-hydroxyprostaglandin dehydrogenase)** | -0·70 (-0·99; -0·41) |
| **Perioperative BTN3A1 (butyrophilin 3A1)** | 2·58 (1·68; 3·49 |
| **Perioperative LAP3 (leucine aminopeptidase 3)** | 1·36 (0·58; 2·13) |
| **Perioperative DSN1 (DSN1 component of MIS12 kinetochore complex)** | -0·09 (-1·12; 0·95) |
| **Preoperative KIF4B (kinesin family member 4B)** | 0·38 (-0·86; 1·62) |
| **Preoperative JAK2 (janus kinase 2)** | 1·54 (0·58; 2·51) |
| **Preoperative circular JAK2 (janus kinase 2 circular mRNA)** | 1·37 (0·63; 2·10) |

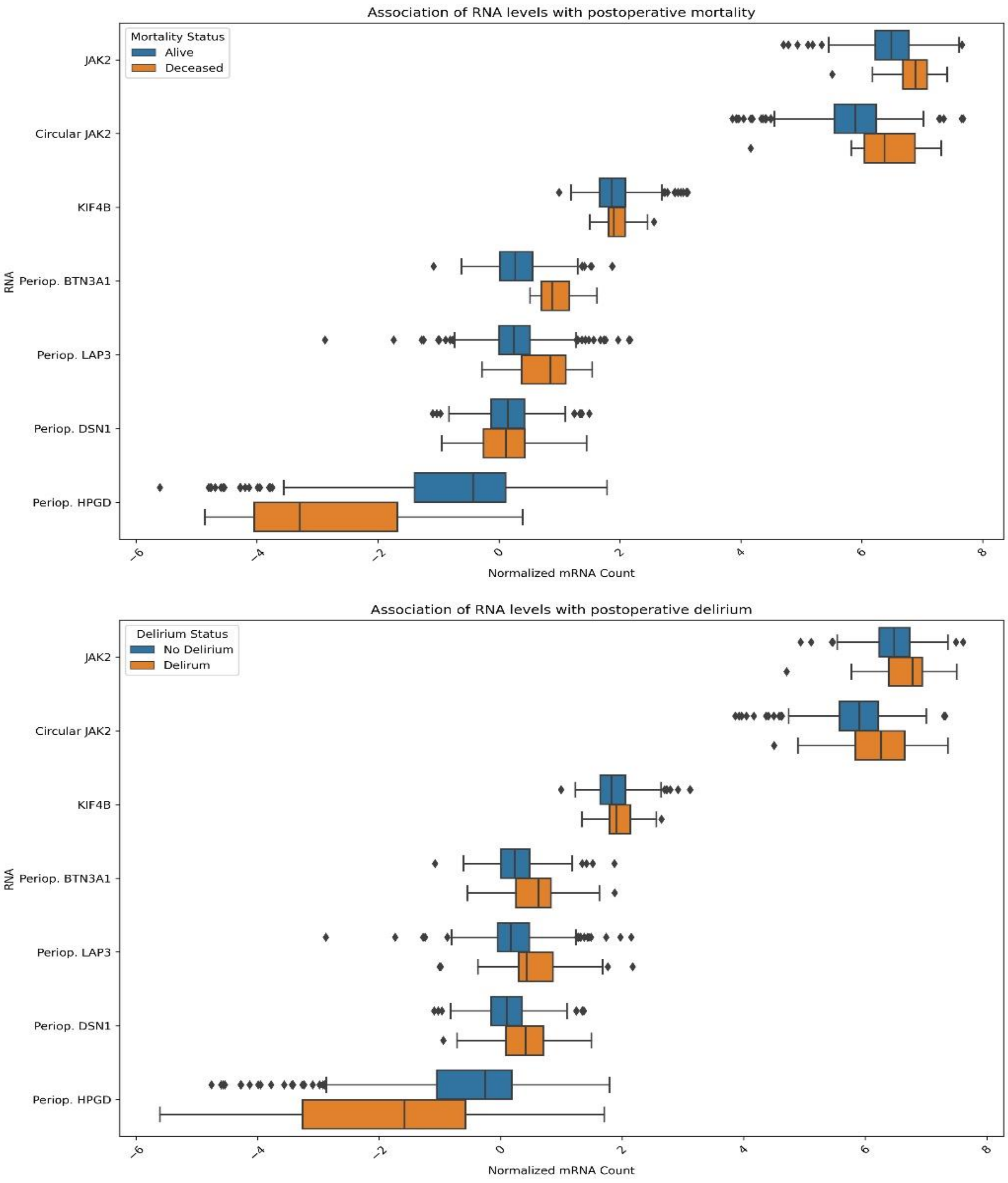


**EFIGURE 11: PREDICTIVE TRANSCRIPTS STRATIFIED BY 90D MORTALITY (TOP) AND POSTOPERATIVE DELIRIUM.**

Abbreviations: Periop.: perioperative, referring to perioperative change in mRNA abundance

### eChapter 3: References


1. Fried LP, Tangen CM, Walston J, Newman AB, Hirsch C, Gottdiener J, et al. Frailty in older adults: evidence for a phenotype. J Gerontol A Biol Sci Med Sci. 2001;56(3):M146-56.
2. Ensrud KE, Ewing SK, Taylor BC, Fink HA, Cawthon PM, Stone KL, et al. Comparison of 2 frailty indexes for prediction of falls, disability, fractures, and death in older women. Arch Intern Med. 2008;168(4):382-9.
3. Ludwig K, Graf von der Schulenburg JM, Greiner W. German Value Set for the EQ-5D-5L. Pharmacoeconomics. 2018;36(6):663-74.
4. Versteegh MM, Vermeulen KM, Evers SMAA, de Wit GA, Prenger R, Stolk EA. Dutch Tariff for the Five-Level Version of EQ-5D. Value Health. 2016;19(4):343-52.
5. Herdman M, Gudex C, Lloyd A, Janssen M, Kind P, Parkin D, et al. Development and preliminary testing of the new five-level version of EQ-5D (EQ-5D-5L). Qual Life Res. 2011;20(10):1727-36.
6. Stekhoven DJ, Buhlmann P. MissForest--non-parametric missing value imputation for mixed-type data. Bioinformatics. 2012;28(1):112-8.
7. Feinkohl I, Borchers F, Burkhardt S, Krampe H, Kraft A, Speidel S, et al. Stability of neuropsychological test performance in older adults serving as normative controls for a study on postoperative cognitive dysfunction. BMC Res Notes. 2020;13(1):55.
8. Kauffmann A, Gentleman R, Huber W. arrayQualityMetrics--a bioconductor package for quality assessment of microarray data. Bioinformatics. 2009;25(3):415-6.
9. Desikan RS, Segonne F, Fischl B, Quinn BT, Dickerson BC, Blacker D, et al. An automated labeling system for subdividing the human cerebral cortex on MRI scans into gyral based regions of interest. Neuroimage. 2006;31(3):968-80.
10. Yushkevich PA, Pluta JB, Wang H, Xie L, Ding SL, Gertje EC, et al. Automated volumetry and regional thickness analysis of hippocampal subfields and medial temporal cortical structures in mild cognitive impairment. Hum Brain Mapp. 2015;36(1):258-87.
11. Yushkevich PA, Wang H, Pluta J, Das SR, Craige C, Avants BB, et al. Nearly automatic segmentation of hippocampal subfields in in vivo focal T2-weighted MRI. Neuroimage. 2010;53(4):1208-24.
12. Ashburner J. A fast diffeomorphic image registration algorithm. Neuroimage. 2007;38(1):95-113.
13. Zaborszky L, Hoemke L, Mohlberg H, Schleicher A, Amunts K, Zilles K. Stereotaxic probabilistic maps of the magnocellular cell groups in human basal forebrain. Neuroimage. 2008;42(3):1127-41.
14. Lammers F, Borchers F, Feinkohl I, Hendrikse J, Kant IMJ, Kozma P, et al. Basal forebrain cholinergic system volume is associated with general cognitive ability in the elderly. Neuropsychologia. 2018;119:145-56.
15. Veraart J, Fieremans E, Novikov DS. Diffusion MRI noise mapping using random matrix theory. Magn Reson Med. 2016;76(5):1582-93.
16. Kellner E, Dhital B, Kiselev VG, Reisert M. Gibbs-ringing artifact removal based on local subvoxel-shifts. Magn Reson Med. 2016;76(5):1574-81.
17. Neto Henriques R. Advanced Methods for Diffusion MRI Data Analysis and their Application to the Healthy Ageing Brain 2018.
18. Andersson JLR, Sotiropoulos SN. An integrated approach to correction for off-resonance effects and subject movement in diffusion MR imaging. Neuroimage. 2016;125:1063-78.
19. Jeurissen B, Tournier JD, Dhollander T, Connelly A, Sijbers J. Multi-tissue constrained spherical deconvolution for improved analysis of multi-shell diffusion MRI data. Neuroimage. 2014;103:411-26.
20. Smith SM, Jenkinson M, Woolrich MW, Beckmann CF, Behrens TE, Johansen-Berg H, et al. Advances in functional and structural MR image analysis and implementation as FSL. Neuroimage. 2004;23 Suppl 1:S208-19.
21. Ronneberger O., Fischer P., T. B. U-Net: Convolutional Networks for Biomedical Image Segmentation. In: Navab N, Hornegger J, Wells WM, Frangi AF, editors. Medical Image Computing and Computer-Assisted Intervention – MICCAI 2015: Springer International Publishing; 2015. p. 234-41.

22. Wasserthal J, Neher P, Maier-Hein KH. TractSeg - Fast and accurate white matter tract segmentation. Neuroimage. 2018;183:239-53.
23. Cook NR. Use and misuse of the receiver operating characteristic curve in risk prediction. Circulation. 2007;115(7):928-35.
